# Single-molecule cfDNA sequencing establishes clinical utility for ecDNA monitoring and multimodal liquid biopsy analysis

**DOI:** 10.64898/2026.04.08.26350410

**Authors:** Carolin M. Sauer, Nicholas Tovey, Anetta Ptasinska, Debbie Hughes, Joanne Stockton, Sonia Zumalave, Alistair G. Rust, Claire Lynn, Virginia Livellara, Francois Sevrin, Courtney Himsworth, Francesc Muyas, Marilena Nicolaidou, Georgia Parry, Eunice Paisana, Rita Cascão, Saira Waqar Ahmed, Shireena A. Yasin, Laura Rey Portela, Pooja Balasubramanian, G.A. Amos Burke, Aditi Vedi, Claudia C. Faria, Lynley V. Marshall, Thomas S. Jacques, Michael Hubank, Darren Hargrave, Sally George, Paola Angelini, John Anderson, Louis Chesler, Andrew D. Beggs, Isidro Cortés-Ciriano

## Abstract

Cell-free DNA (cfDNA) profiling enables minimally invasive cancer detection and monitoring. We present SIMMA, a low-input single-molecule sequencing approach that enables multimodal whole-genome and high-depth targeted sequencing of the same cfDNA sample for both tumour-agnostic and tumour-informed liquid biopsy analysis. Across 792 plasma and cerebrospinal fluid cfDNA samples from 277 paediatric patients with diverse brain and extracranial tumours, SIMMA enabled tumour diagnosis, detection of driver mutations, and reconstruction of extrachromosomal DNA (ecDNA) months before clinical relapse. Using conformal prediction trained on genome-wide fragmentomics, genomic and epigenomic data, SIMMA predicts disease burden as a continuous variable and provides well-calibrated uncertainty estimates for each sample, achieving a limit of detection of ∼100 ppm from low-pass whole-genome sequencing data. In summary, SIMMA establishes the clinical utility of multimodal cfDNA profiling with uncertainty quantification for individual patients and unlocks the potential of ecDNA as a liquid biopsy biomarker for disease detection and monitoring across diverse aggressive malignancies.

## Introduction

Molecular profiling of tumour samples has become a cornerstone of precision cancer medicine. However, obtaining tumour material through tissue biopsy or surgical resection is not always clinically feasible. Moreover, tissue biopsies are not amenable to serial sampling for longitudinal disease monitoring and fail to capture the genomic heterogeneity within and across tumour sites^1,2^. To circumvent these challenges, analysis of circulating tumour DNA (ctDNA) through molecular profiling of cell-free DNA (cfDNA) in plasma or other body fluids^3,4^ has emerged as a minimally invasive alternative to tissue-based tumour sampling, also enabling cancer detection and minimal residual disease (MRD) monitoring^5–8^.

Existing liquid biopsy assays fall into two major paradigms. Tumour-informed approaches focus on detecting patient-specific somatic mutations previously identified through sequencing of tumour samples. This strategy can achieve limits of detection (LOD) in the parts per million (ppm) range, which has proven particularly powerful for ultra-sensitive MRD monitoring^9–13^. However, the need to access and analyse tumour tissue samples limits scalability, deployment and reporting turnaround times^11^. Moreover, tumour-informed approaches are not suitable for cancer type classification or screening. By contrast, tumour-agnostic methods rely on the detection of shifts in genome-wide patterns in cfDNA associated with the presence of ctDNA, such as somatic copy number aberrations (SCNAs)^14^, epigenomic alterations^15–19^ or changes in cfDNA fragmentation^20,21^. While more suitable for multi-cancer detection, tumour-agnostic methods have lower sensitivity. Other tumour-agnostic methods rely on the detection of cancer-associated mutations. However, cfDNA fragments harbouring somatic mutations arising from non-neoplastic clonal expansions can confound ctDNA detection, thereby limiting specificity^22^. Therefore, multimodal liquid biopsy methods combining the strengths of both approaches are needed to deliver accurate disease diagnosis and monitoring, as well as to inform therapeutic intervention across molecularly diverse cancer types.

Integrating genomic and epigenomic information has the potential to improve the sensitivity and specificity of cfDNA analysis^16,18^. However, existing multimodal cfDNA profiling approaches require multiple short-read sequencing assays, which may not be feasible due to limited cfDNA yields from plasma samples. Moreover, short-read sequencing fails to resolve repetitive regions, read out long DNA fragments or reconstruct the diversity of somatic structural variants (SVs) underpinning malignant transformation^2,23^ and drug resistance^24–26^. By contrast, single-molecule sequencing technologies permit simultaneous detection of genomic aberrations and DNA modifications in individual native DNA molecules^27–30^, including cfDNA^29^. Yet the potential of single-molecule sequencing for liquid biopsy analysis remains largely unrealised due to the lack of experimental protocols compatible with low-input cfDNA and computational methods for multimodal data analysis and integration.

To overcome these limitations, we developed SIMMA (**S**ingle-molecule **I**ntegrative **M**ulti**M**odal **A**nalysis of cfDNA), a novel liquid biopsy assay for low-input single-molecule sequencing of cfDNA that combines whole-genome sequencing (WGS) and high-depth targeted sequencing of the same cfDNA sample with multimodal data integration. Specifically, SIMMA utilises multimodal machine learning with well-calibrated uncertainty quantification to predict tumour fraction (TF) values from low-pass cfDNA WGS data in a tumour-agnostic manner with a LOD of ∼100 ppm. Analysis of 792 cfDNA samples from 277 patients with solid paediatric cancers shows that SIMMA can inform tumour detection, monitoring and classification, detect driver mutations and reconstruct complex genomic rearrangements in cfDNA, including ecDNA, months before clinical detection.

## Results

### Scalable multimodal single-molecule sequencing of low-input cfDNA

To enable multimodal liquid biopsy analysis using low input volumes of cfDNA we developed SIMMA (**Fig. 1a; Methods**). Briefly, cfDNA is end-repaired and cleaned using beads. One aliquot of end-repaired DNA is prepared for nanopore WGS by ligating native barcodes to enable multiplexing. Next, 16 barcoded cfDNA samples are pooled, sequencing adaptors ligated, and the multiplexed samples analysed via nanopore long-read WGS. To maximise sequencing output, we optimized the efficiency of ligation and elution times, thereby enabling the analysis of low input cfDNA samples (**Methods**). Benchmarking analysis using a 30-fold dilution series of NA12878 DNA in the 5-150ng range revealed that SIMMA yields a two-fold increase in sequencing output compared to commercial protocols (**Fig. 1b; Supplementary Fig. 1a-f**). Importantly, the sequencing output generated for each sample is proportional to the amount of input DNA (*R* = 0.97, *P*<0.0001, Pearson correlation), even for highly imbalanced multiplexed sequencing runs (**Fig. 1c**).

**Figure 1.**
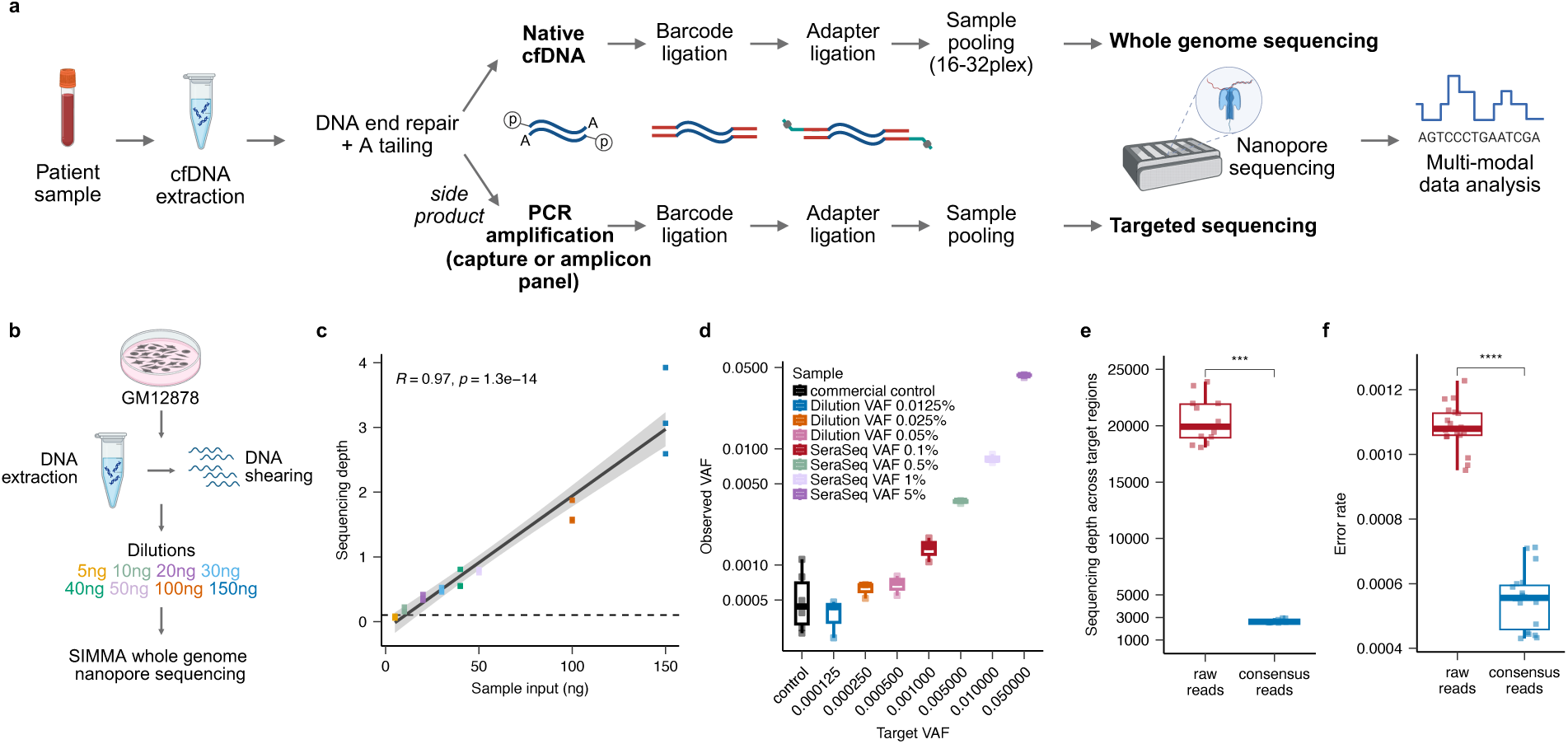
Overview and performance of SIMMA. **(a)** Overview of the SIMMA workflow for whole-genome sequencing of native DNA and targeted sequencing of amplified DNA from low input cfDNA samples. **(b)** Overview of the experimental design implemented to evaluate the efficiency of SIMMA for multiplexed whole-genome sequencing. **(c)** Pearson’s correlation between the total DNA input (ng) vs sequencing data output. Each dot corresponds to the mean across three replicated. The dashed horizontal line indicates a sequencing depth of 0.1x, which was obtained with 5ng of input DNA. **(d)** Boxplots reporting the VAF value of mutations in contrived samples estimated after UMI-based consensus read calling. Each dot corresponds to a mutation. The boxplots show the VAF values detected across six commercial cancer-free control cfDNA samples and a dilution series generated using the SeraSeq® ctDNA Complete™ Reference Material. **(e)** Total sequencing depth achieved using SIMMA before and after UMI-based consensus read calling. **(f)** Sequencing error rate estimated based on 375 homozygous sites before and after UMI-based consensus read calling. cfDNA: cell-free DNA; ng: nanograms; UMI: unique molecular identifier; VAF: variant allele frequency.

Optionally, the remaining end-repaired DNA can be used for targeted sequencing (**Methods**). To assess sensitivity, we analysed commercially available contrived samples containing SNVs and SCNAs in cancer genes using a custom capture panel with unique molecular identifiers (UMIs) spanning 76 cancer genes (**Supplementary Fig. 1g; Supplementary Table 1; Methods**). The variant allele frequency (VAF) values for mutations estimated using SIMMA were concordant with the reference values (**Fig. 1d; Methods**). After UMI error correction and consensus read calling using a custom approach (**Methods**), we estimated an error rate of ∼5×10^−4^ and obtained an effective mean on-target sequencing depth of 2,282x (range 1,230-2,956x; **Fig. 1e-f**). This performance was consistent even when multiplexing three samples per flow cell with as little as 1ng of input DNA.

Together, these results show that SIMMA enables scalable multimodal whole-genome and targeted sequencing from the same cfDNA sample with as little as 5ng of total input DNA.

### Validation of the clinical utility of SIMMA in paediatric cancer at a national scale

To evaluate the clinical utility of SIMMA, we analysed 792 cfDNA samples from 277 patients with paediatric solid tumours and 81 plasma samples from cancer-free controls (**Fig. 2a-b; Supplementary Fig. 2; Supplementary Table 2-3**). These included 260 plasma samples collected from 218 patients with clinical relapse or treatment-refractory disease enrolled in the Stratified Medicine Paediatrics (SMPaeds) programme between 2019 and 2022 across 20 paediatric oncology centres in the UK^31^. In addition, we analysed 474 plasma and 58 cerebrospinal fluid (CSF) cfDNA samples collected serially from 59 patients with paediatric solid tumours (median follow-up time: 23.8 months; range 1.6-52.5 months). The most frequent diagnoses in our combined cohort were sarcoma (*n*=84), neuroblastoma (*n*=71), glioma and astrocytoma (*n*=22), Wilms tumour (*n*=19) and medulloblastoma (*n*=15; **Fig. 2c and Supplementary Fig. 2**). A total of 19 patients had known cancer predisposition syndromes, including Li-Fraumeni (*n*=4), Neurofibromatosis type 1 (*n*=3), *DICER1* syndrome (*n*=2), Gorlin syndrome (*n*=1) and Beckwith-Wiedemann syndrome (*n*=1; **Supplementary Fig. 2**). Similar to the dilution experiment shown in **Fig. 1b-c**, the amount of input DNA strongly correlated with sequencing depth, which averaged to 0.5x across all samples (range 0.001-14.6x; **Supplementary Fig. 3a-b**). Of note, this yield was limited by the reduced availability of remaining cfDNA material after delivering short-read cfDNA analyses by SMPaeds^31^ rather than by sequencing capacity.

**Figure 2.**
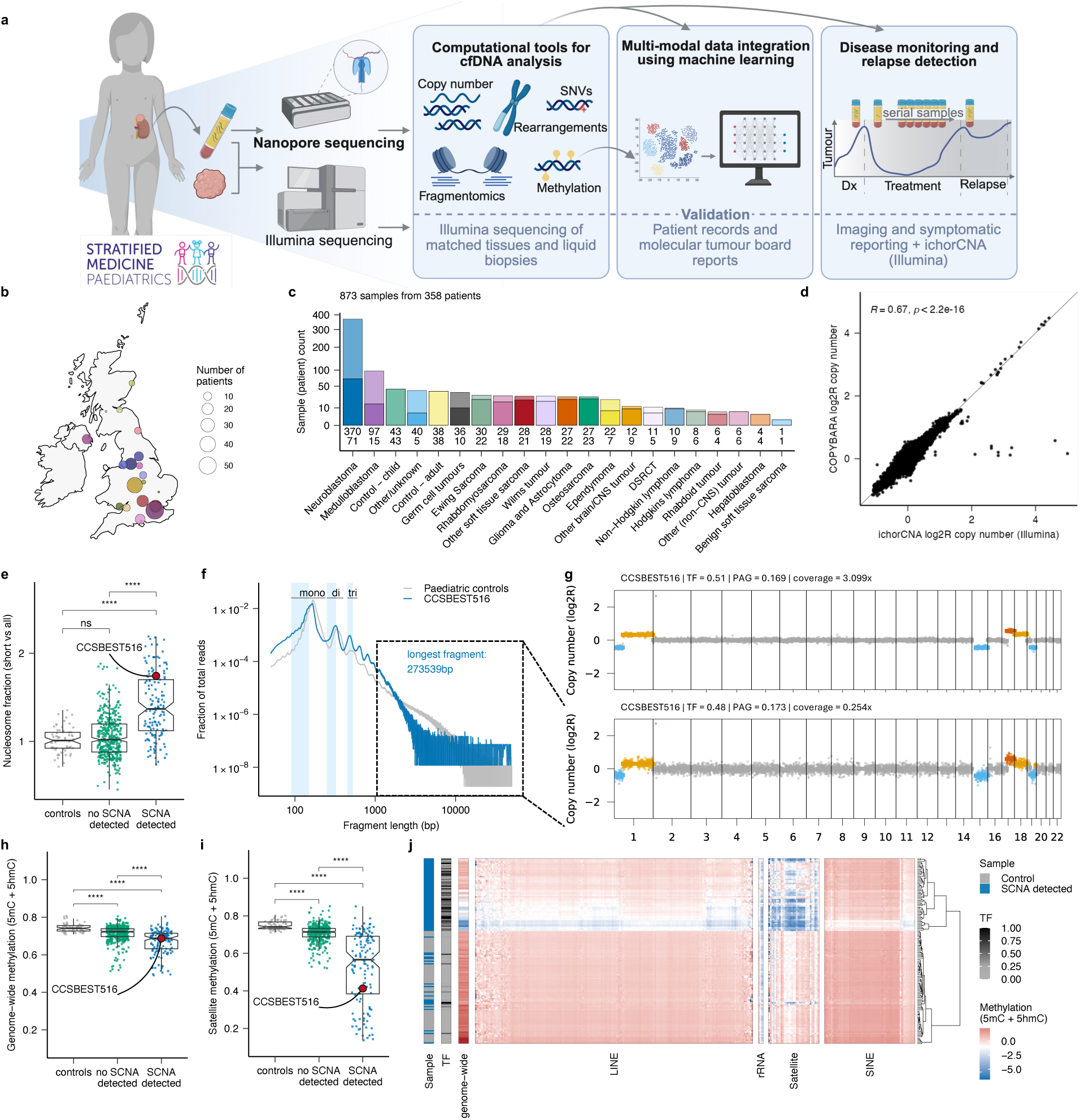
Multimodal cfDNA data analysis with SIMMA. **(a)** Schematic representation of the validation of the clinical utility of SIMMA across paediatric solid tumours. **(b)** Distribution of paediatric cancer hospitals across the UK involved in the SMPaeds study. The size of the circles is proportional to the number of patients enrolled in SMPaeds at each site. **(c)** Bar plot showing the number of patients (bold colour) and samples (pale colour) included in this study across cancer types. Total number of patients (bottom) and samples (top) are shown below each bar. **(d)** Correlation between the normalized read counts estimated for 500kbp bins across the genome using aliquot-matched matched Illumina and nanopore cfDNA WGS data (n=198; *R*=Pearson’s correlation coefficient). **(e)** Fraction of sequencing reads shorter than the expected modal length for mono-, di- and tri- nucleosomal fragments for cancer-free cfDNA samples (black), cfDNA samples from cancer patients in which no somatic SCNAs were detected (green), and samples in which SCNAs were detected using COPYBARA (blue). **(f)** Fragment size distribution computed using cfDNA WGS data from cfDNA sample CCSBEST516 (blue) and paediatric cancer-free controls (grey). The size ranges corresponding to fragments shorter than the expected modal length for mono-, di- and tri-nucleosomal fragments in cfDNA from cancer-free controls are highlighted by blue boxes. **(g)** COPYBARA copy number profile for CCSBEST516 before (top) and after (bottom) COPYBARA *in silico* size selection of reads >1000bp in length. **(h)** Fraction of methylated CpG sites (5mC + 5hmC) across the genome in control samples, and samples with or without SCNAs detected using COPYBARA. **(i)** Fraction of methylated CpG sites (5mC + 5hmC) across satellite regions in control samples, and samples with or without SCNA detected using COPYBARA. (**j**) Heatmap showing scaled 5mC + 5hmC methylation levels across repetitive regions. Rows correspond to cfDNA samples sorted based on unsupervised clustering. Only cfDNA samples from cancer-free individuals and cancer patients in which COPYBARA detected ctDNA signal based on SCNA analysis are shown. bp: base pairs; DSRCT: desmoplastic small round cell tumour; PAG: percentage abnormal genome; SCNA: somatic copy number aberration; TF: tumour fraction.

SMPaeds previously established the utility of low-pass WGS and targeted Illumina sequencing of plasma cfDNA for patients with relapsed or refractory brain and solid tumours^31,32^. The availability of clinical reports and matched liquid biopsy results makes this cohort ideal to assess the clinical utility of SIMMA. Thus, we sought to compare SIMMA results with clinical reports of tumour sequencing and cfDNA analyses, including 193 cfDNA samples for which we had matched Illumina and SIMMA results. To detect SCNAs in SIMMA data we developed COPYBARA. In brief, COPYBARA detects large and focal SCNAs, infers tumour ploidy and estimates the TF in cfDNA (**Methods**; **Supplementary Fig. 4**). The SCNAs detected using COPYBARA were highly concordant with matched tumour and cfDNA Illumina sequencing data despite a one-order-of-magnitude difference in coverage between the two datasets (**Fig. 2d**; **Supplementary Figs. 3c, 5-9**). Targeted sequencing of a cancer gene panel identified somatic point mutations in driver genes, including *ALK, TP53, MYC, CCND1* and *ATRX* (**Supplementary Fig. 10-11**). Driver mutations were detected in matched CSF and plasma cfDNA samples from patients with paediatric brain tumours (**Supplementary Fig. 10b**). Mutation frequencies of pathogenic coding SNVs detected in cfDNA averaged to 0.012 (range: 0.0014-0.026; **Supplementary Fig. 10c**), consistent with previous reports^33^.

Fragment size distributions across cancer samples showed the expected pattern and shift towards shorter cfDNA fragments. This effect was observed for mono-, di- and tri-nucleosomal cfDNA fragments, including in samples with undetectable ctDNA signal based on SCNA analysis (**Fig. 2e; Extended Data Fig. 1a-b**). *In silico* fragment size analysis revealed increased sensitivity for ctDNA detection when jointly analysing fragments shorter than the expected modal length for mono-, di- and tri-nucleosomal cfDNA fragments (**Extended Data Fig. 1c-g; Methods**). As opposed to short-read sequencing, SIMMA also enabled detection and sequencing of long native DNA fragments. On average, 2.1% of cfDNA sequencing reads were longer than 1,000bp (range 0.016-39.2%; **Fig. 2f**). The top 1% of fragments (*n*=41,453) had a median length of 30,494bp. Genome-wide copy number profiles estimated using all cfDNA reads or only those longer than 1,000bp were highly concordant across multiple cancer types, indicating that plasma cfDNA from patients with cancer contains a subset of long tumour-derived fragments (**Fig. 2f-g; Extended Data Fig. 1c-g**).

Analysis of genome-wide 5-methylcytosine (5mC) and 5-hydroxymethylcytosine (5hmC) DNA methylation patterns revealed a significantly higher degree of genome-wide hypomethylation in cfDNA (**Fig. 2h**), consistent with the loss of DNA methylation commonly observed in human tumours^34,35^. Next, we investigated the degree of hypomethylation across repeat classes by aligning sequencing reads to the gapless human genome assembly T2T-CHM13^36^. We detected broad hypomethylation across repeat classes, with the strongest effect observed for satellite repeats (**Fig. 2i-j and Extended Data Fig. 2a-b**). Notably, we observed significant differences in genome-wide methylation in samples with and without detectable SCNAs. This indicates that genome-wide methylation changes in repetitive regions, such as centromeres, which are currently understudied due to the limitations of short-read sequencing, enable sensitive ctDNA detection. Additionally, unbiased clustering analysis of genome-wide 5mC and 5hmC patterns grouped cfDNA samples by diagnosis, showing that SIMMA enables tumour type classification (**Extended Data Fig. 2c**).

### Detection and reconstruction of ecDNA molecules in cfDNA

Many of the most aggressive paediatric and adult solid tumours are driven by complex genomic aberrations, including chromothripsis and oncogene amplification in ecDNA, which often arise early during tumour evolution^2,37–39^. Despite the potential of ecDNA detection to inform prognosis^38,40^, clinical trial design and therapeutic development^41^, our understanding of the genomic and epigenomic properties of ecDNA in cfDNA remains limited, thus hindering our ability to detect ecDNA in liquid biopsies^42^.

Due to the high number of copies of ecDNA molecules in cancer cells^38^ and the potential of long reads to reconstruct complex DNA rearrangements^43^, including ecDNA^2,44,45^, we first aimed to detect ecDNA-associated breakpoints in cfDNA using SAVANA^43^ (**Methods**). We identified ecDNA molecules and associated rearrangements in 18 samples from 10 patients. These included circular and complex structures generated by chromothripsis events involving multiple genomic loci and >15 breakpoints, some of which were also detected using CoRAL^44^ (**Fig. 3a-b, Extended Data Fig. 3 and Supplementary Figs. 12-14**). The sequencing depth at ecDNA regions was several hundred-fold higher than that of chromosomal DNA (**Fig. 3a**). For example, we detected >800 copies of a complex ecDNA molecule carrying a *MYCN* amplification in a neuroblastoma cfDNA sample with an average genome-wide sequencing depth of just 0.6x (**Fig. 3b**), indicating that the high copy number of ecDNA enables its detection even at low sequencing depth.

**Figure 3.**
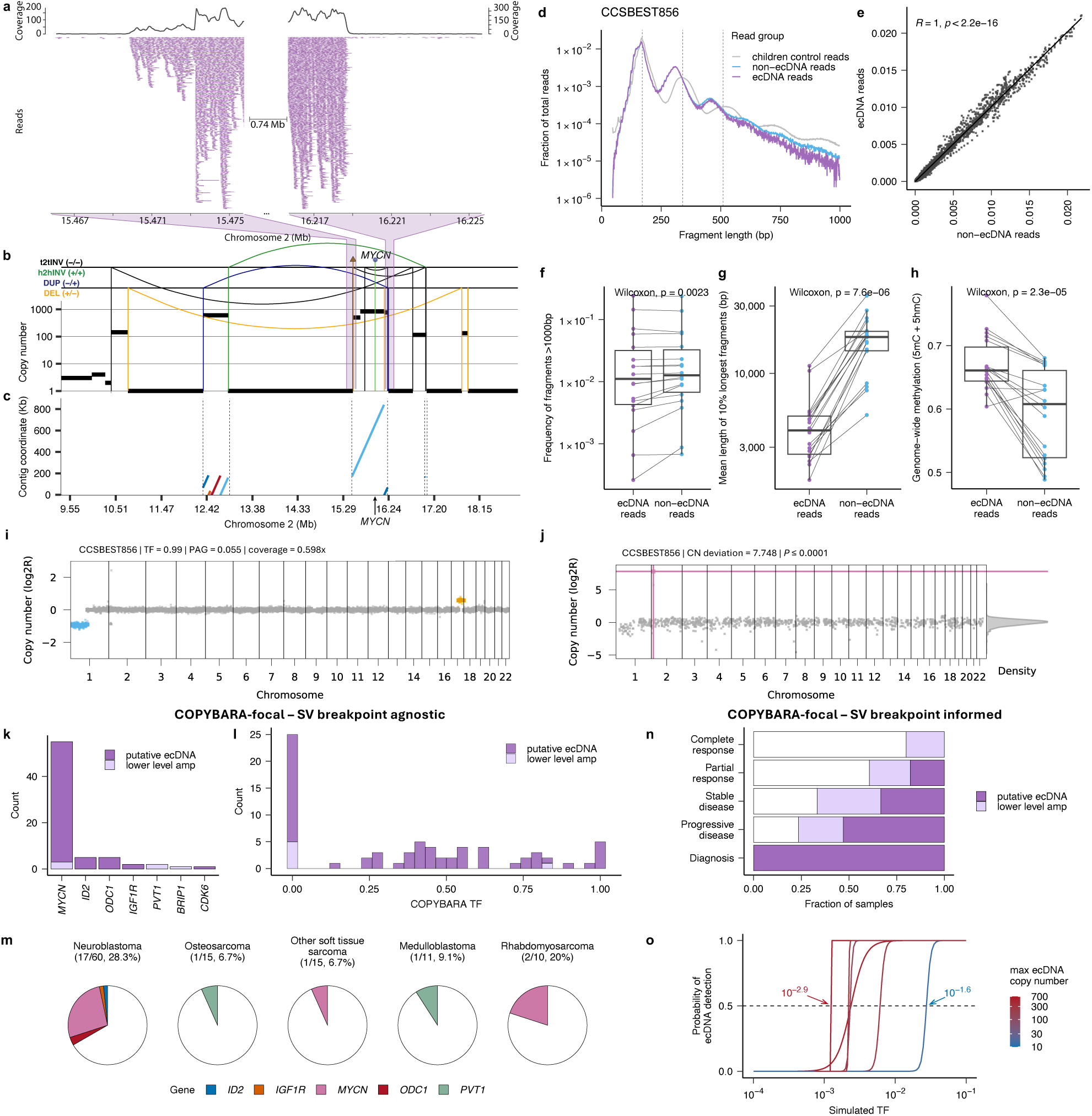
Detection and reconstruction of ecDNA in cfDNA. **(a)** Aligned cfDNA reads from sample CCSBEST856 mapping to a highly amplified region on chromosome 2 containing *MYCN*. **(b)** Genomic rearrangement profile detected in cfDNA sample CCSBEST856 using SAVANA. The areas highlighted in purple represent the coverage boundaries at rearrangement breakpoints depicted in the coverage track in (**a**). Absolute copy number data is represented by the black horizontal lines. SVs are represented by vertical lines. **(c)** Assembly of the ecDNA structure generated by the rearrangements shown in (**b**) using nanopore cfDNA reads. The plot shows the alignment of the assembled contigs to chromosome 2 of the human reference genome. **(d)** Fragment size profile of cfDNA reads mapping to ecDNA (purple) and non-ecDNA (blue) regions for sample CCSBEST856. The fragmentation profile computed using cfDNA WGS data from 24 paediatric control samples is shown in grey for comparison. **(e)** Correlation of fragmentation size profiles mapping to ecDNA and non-ecDNA regions for all ecDNA-positive samples (n=18). **(f)** Frequency of cfDNA fragments >1,000bp in length mapping to ecDNA and non-ecDNA regions from ecDNA-positive samples. **(g)** Mean length of the 10% longest cfDNA fragments >1,000bp mapping to ecDNA and non-ecDNA regions from ecDNA-positive samples. **(h)** Fraction of 5mC and 5hmC at CpG dinucleotides across reads mapping to ecDNA and non-ecDNA regions from ecDNA-positive samples. **(i)** Genome-wide copy number profile for sample CCSBEST856 generated using COPYBARA. Losses and gains are shown in light blue and yellow, respectively. **(j)** Detection of *MYCN* ecDNA (pink dot) using COPYBARA-focal in sample CCSBEST856. **(k)** Bar plot showing the number of all paediatric cancer cfDNA samples (n=792) with ecDNA amplifying oncogenes detected using COPYBARA-focal. **(l)** Bar plot showing the number of samples with ecDNA amplifying oncogenes detected using COPYBARA-focal across tumour fraction (TF) values. **(m)** Fraction of patients with oncogenes amplified in ecDNA detected in cfDNA using COPYBARA-focal across paediatric malignancies with data available for at least 10 patients. **(n)** Bar plot showing the fraction of samples from ecDNA-positive patients in which ecDNA were detected by disease status. For this analysis, SV breakpoint coordinates mapping to the boundaries of genomic regions amplified in ecDNA were used to inform ecDNA detection using COPYBARA-focal. **(o)** LOD for ecDNA detection in cfDNA using COPYBARA-focal. SV breakpoint coordinates mapping to the boundaries of genomic regions amplified in ecDNA were used to inform ecDNA detection across simulated dilutions with TF values ranging from 10^−6^ to 0.9 generated for six ecDNA-positive cfDNA samples. Logistic regression models (lines) were fitted to binary detection outcomes across dilution series for each sample. The LOD for each sample was estimated as the TF corresponding to 50% detection probability (indicated by dashed line). The colour of each line corresponds to the ecDNA copy number detected in each of the six cfDNA-positive samples analysed. Box plots in **f-h** show the median, first and third quartiles (boxes) and the whiskers encompass observations within 1.5× the interquartile range from the first and third quartiles. Amp.: amplification; ecDNA: extrachromosomal DNA; Mb: metabases; Kb: kilobases; bp: base pairs; DEL: deletion-like rearrangement; DUP: duplication-like rearrangement; h2hINV: head-to-head inversion; t2tINV, tail-to-tail inversion; SCNA: somatic copy number aberration; SV: structural variant; TF: tumour fraction; PAG: percentage abnormal genome; bp: base pairs.

Next, we sought to reconstruct ecDNA molecules through *de novo* genome assembly of cfDNA reads mapping to ecDNA. For example, we generated an assembly comprising 6 contigs for an ecDNA molecule of 1.3Mb in size (contig size range: 27,737 - 843,316bp; N50: 843,316bp; **Fig. 3c**). Sequencing reads mapping to ecDNA molecules followed the characteristic fragment size distribution expected for cfDNA derived from chromosomal tumour DNA across all ecDNA-positive samples (**Fig. 3d-e; Extended Data Fig. 3; Supplementary Figs. 12-14**). The fraction of cfDNA fragments longer than 1,000bp mapping to ecDNA and non-ecDNA regions was comparable (**Fig. 3f**). However, the longest cfDNA fragments detected mapped to non-ecDNA regions (**Fig. 3g**). Similarly, we consistently detected significantly higher levels of 5mC and 5hmC in ecDNA reads (**Fig. 3h**). Collectively, these results indicate that ecDNA molecules are fragmented in cfDNA, and that chromosomal DNA rather than ecDNA is the main source of the longest DNA fragments and hypomethylation detected in cfDNA from cancer patients.

Motivated by these results, we developed COPYBARA-focal, an algorithm to detect ecDNA in cfDNA samples even with a low TF or sequencing depth. In brief, COPYBARA-focal determines the number of ecDNA copies in either a tumour-agnostic or tumour-informed manner by assessing differences in sequencing depth at regions of interest, e.g. oncogenes, over a sample-specific background (**Fig. 3i-j; Methods**). Using COPYBARA-focal in a tumour-agnostic manner, we detected putative ecDNA molecules in 8.2% (65/792) of all cfDNA samples from patients with cancer (**Fig. 3k**). Notably, we detected ecDNA in cfDNA samples spanning a dynamic TF range, including samples with no detectable ctDNA signal based on genome-wide SCNA analysis (**Fig. 3l; Extended Data Fig. 4a**). *MYCN* amplification was the most frequent event, consistent with the enrichment of neuroblastomas in our cohort and the high frequency of oncogene amplification in ecDNA in this disease^46^ (**Fig. 3m**).

Next, we harnessed ecDNA-associated breakpoint coordinates to inform ecDNA detection in serial samples. This analysis detected ecDNA in samples collected across all RECIST 1.1 (Response Evaluation Criteria in Solid Tumors) categories (**Fig. 3n**). Furthermore, to assess the LOD of COPYBARA-focal for ecDNA detection, we generated serial dilutions by mixing sequencing reads from cancer-free controls and 6 ecDNA-positive cfDNA samples with a high TF (**Methods**). This analysis estimated an LOD of ∼10^−3^, varying depending on ecDNA copy number (**Fig. 3o).** Notably, COPYBARA-focal did not detect ecDNA in cancer-free controls (**Extended Data Fig. 4b-c**). This indicates both high sensitivity and specificity even for samples with a genome-wide sequencing depth below 0.5x.

Together, these results show that SIMMA and COPYBARA-focal permit the detection and reconstruction of ecDNA molecules and the associated rearrangements from cfDNA with high specificity and sensitivity in both tumour-informed and tumour-agnostic settings.

### Personalized tumour burden prediction with uncertainty quantification

Previous work has explored the use of machine learning to increase sensitivity for ctDNA detection^47–49^. However, the trustworthiness and clinical application of AI models remain contingent on the ability to provide well-calibrated uncertainty estimates for individual predictions rather than global metrics of model performance that may not translate to prospective samples^50^. Here, we leverage conformal prediction (CP)^51,52^ to integrate the multi-omic features detected in cfDNA using SIMMA and generate well-calibrated confidence intervals for individual TF predictions (**Fig. 4a**). Specifically, CP is a non-parametric mathematical framework compatible with any AI model to quantify the uncertainty of individual predictions at a user-defined confidence level. CP ensures that the confidence regions will always be calibrated provided that data points from the training and external sets are exchangeable, which is a common requirement in predictive modelling^51,52^. For example, at a confidence level of 80%, at least 80% of the predicted confidence intervals will encompass the true value, thus making it possible to control the false positive rate. As a result, the practical utility of CP models is evaluated by their ability to deliver confidence intervals narrow enough to be useful in practice.

**Figure 4.**
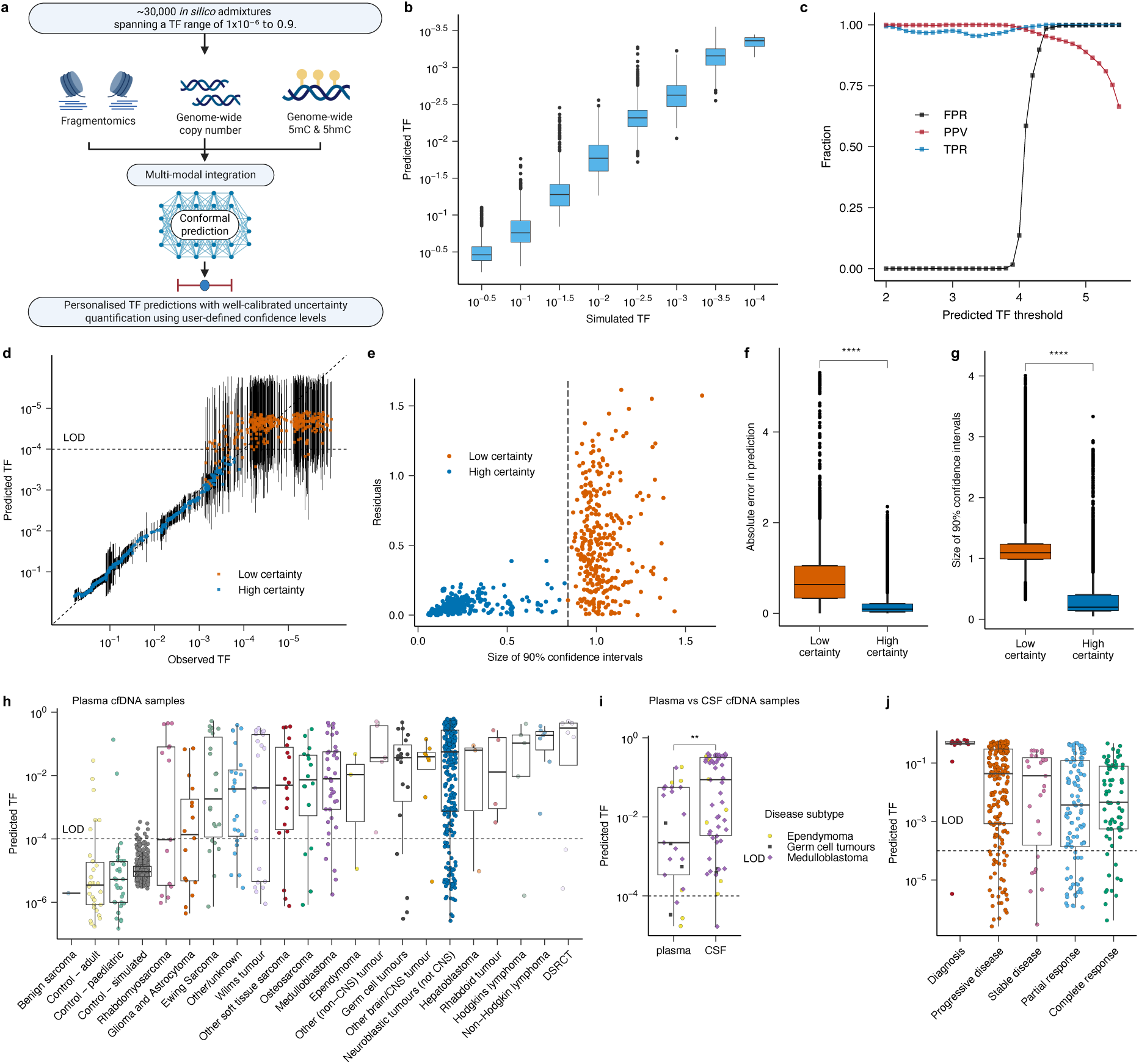
Multimodal prediction of TF with uncertainty quantification for individual patients using multimodal Conformal Prediction. (**a**) Schematic representation of the Conformal Prediction (CP) workflow implemented in this study for the prediction of the TF for individual cfDNA samples. (**b**) Correlation between simulated and predicted TF values across LOPO models for simulations with a TF value above the LOD of 10^−4^. (**c**) Assessment of the false positive rate (FPR), positive predictive value (PPV) and true positive rate (TPR) for leave-one-patient-out CP models. (**d**) Correlation between simulated and predicted TF values using the CP model trained on all *in silico* admixtures excluding those from patient G0064. The predicted 90% confidence intervals for each dilution generated for patient G0064 are shown. Predictions with low and high certainty based on the size of the predicted 90% confidence intervals are shown in orange and blue, respectively. (**e**) Size of the 90% confidence intervals against the absolute error in prediction (residuals) computed using the LOPO model for patient G0064. Each dot corresponds to one *in silico* admixture. The vertical line corresponds to the threshold beyond which errors in prediction show high variance, indicating that predictions are unreliable. Distribution of the absolute error in prediction (**f**) and the size of the predicted 90% confidence intervals (**g**) across all LOPO models. Datapoints in **f-g** are stratified based on the certainty of each prediction as shown in panel (**e**). (**h**) Predicted TF values using multimodal CP for plasma cfDNA samples from cancer-free controls and paediatric cancer patients stratified based on tumour type for all samples with a sequencing depth >0.1x. (**i**) Predicted TF values using multimodal CP for plasma and CSF cfDNA samples from patients with paediatric brain tumours. (**j**) Predicted TF values for plasma cfDNA samples with sequencing depths >0.1x stratified based on RECIST 1.1 categories for all paediatric cancer patients analysed in this study with RECIST 1.1 data available (n=425). Significance was assessed using the Wilcoxon’s test (\*\*\**P* < 0.0001; \*\*\*\**P* < 0.00001). Box plots in **c** and **f-j** show the median, first and third quartiles (boxes) and the whiskers encompass observations within 1.5× the interquartile range from the first and third quartiles. CNS: central nervous system; CSF: cerebrospinal fluid; DSRCT: desmoplastic small round cell tumour; LOD: limit of detection; TF: tumour fraction.

A critical barrier to the application of AI methods in liquid biopsy analysis is the difficulty of accurately measuring true TF values for enough cfDNA samples to enable model training. To address this challenge, we utilised 43 samples with TF values >0.30 estimated using COPYBARA to simulate TF values down to 10^−6^ by mixing sequencing reads from cancer-free controls with each of the selected cfDNA samples to a sequencing depth of 0.1-6x (**Fig. 4a; Methods; Supplementary Table 4**). Each *in silico* admixture was encoded using multi-omic covariates based on SCNAs, fragmentomics and methylation data (**Methods**). Finally, we performed leave-one-patient-out (LOPO) experiments by training Random Forest (RF) regression models on all admixtures except for those from one patient, which were held out to simulate the scenario in which a model would be applied to cfDNA samples from a patient not included in the training set.

Overall, LOPO models achieved high predictive power, with mean R^2^ and RMSE values of 0.95 and 0.51, respectively (**Fig. 4b**), and predicted well-calibrated confidence intervals consistently spanning less than one order of magnitude for reliable predictions even when imposing a 90% confidence level (**Supplementary Fig. 15a**). To quantify the LOD of our models, we computed the false positive rate (FPR) and the true predictive value (PPV) by testing whether the lower bound of the 90% predicted confidence interval for each prediction was higher than an increasingly lower threshold (**Methods**). This analysis revealed an LOD of ∼100 ppm, and a PPV of 99.97% (**Fig. 4c**). Consistent with these results, the predicted and the true TF values showed a strong positive correlation up to the LOD, beyond which the model predictions plateaued, indicating that ctDNA signal in admixtures with TF values below the LOD are undetectable by the model (**Fig. 4d**). Importantly, the absolute error in prediction and the size of the confidence intervals significantly increased for samples below the LOD (**Fig. 4e-g**) and decreased as a function of sequencing depth (**Supplementary Fig. 15b-c**). Therefore, the size of the predicted confidence interval provides a versatile approach to identify predictions beyond the LOD as unreliable.

We next applied the LOPO models to patients with paediatric cancers and controls not considered for model training. TF predictions were overall below the LOD for both adult and paediatric controls and for a benign soft-tissue sarcoma (**Fig. 4h**). We detected ctDNA signal in plasma cfDNA for all cancer types, including brain tumours (**Fig. 4h**), and in cfDNA CSF samples from ependymoma, intracranial germ cell tumour and medulloblastoma patients (**Fig. 4i; Extended Data Fig. 5; Supplementary Fig. 16**). For example, in medulloblastoma patient **R0136**, the TF in CSF was high, revealing a gain of chromosome 7 and i(17q), which were detectable even at genome-wide sequencing depths <0.1x (**Extended Data Fig. 5**). Similarly, in ependymoma patient **R0124**, ctDNA was detectable in both plasma and CSF cfDNA **(Supplementary Fig. 16).**

Together, these results indicate that multimodal data integration using CP permits sensitive and specific detection of ctDNA in both plasma and CSF across molecularly and anatomically diverse paediatric extracranial and brain tumours, while also providing well-calibrated uncertainty estimates for individual predictions.

### Longitudinal disease monitoring through multimodal AI and ecDNA detection

Early detection of relapse and metastatic spread remains an unmet clinical need, especially for paediatric patients with aggressive solid cancers^53^. To evaluate the clinical utility of SIMMA for disease monitoring and early relapse detection, we analysed 532 plasma and CSF cfDNA samples linked with clinical and imaging data from 59 patients with paediatric cancers (median cfDNA samples per patient: 5; **Supplementary Fig. 2**). We predicted a TF value higher than the LOD for 92% of samples collected at diagnosis (12/13), and 83% (162/196) and 76% (88/116) of samples collected at timepoints corresponding to progressive disease (PD) and partial response (PR) according to RECIST 1.1 criteria, respectively (**Fig. 4j**). Moreover, we detected ctDNA signal in 85% of cfDNA samples collected at timepoints when patients were assessed as having a complete response (CR; 62/73) (**Fig. 4j**).

To further investigate the potential of SIMMA for MRD monitoring and early detection of relapse, we first analysed longitudinal cfDNA samples from high-risk neuroblastoma patients and used COPYBARA-focal to monitor the number of ecDNA copies in cfDNA longitudinally (**Methods**). Overall, the copy number of ecDNA molecules amplifying *MYCN* correlated with imaging and clinical data throughout treatment, and enabled detection of disease progression before clinical relapse. For example, in patient **R0118,** diagnosed with metastatic neuroblastoma, the largely cystic and necrotic left adrenal mass reduced in size during the first two months of standard firstline chemotherapy treatment, indicating a partial response. At this timepoint, cfDNA analysis detected both ctDNA signal using multimodal CP as well as an ecDNA amplifying *MYCN* against the backdrop of a diploid genome (Timepoint 1 in **Fig. 5a-b; Supplementary Fig. 12c**). Upon disease progression with disseminated skeletal metastasis after completion of initial Rapid COJEC (cisplatin [C], vincristine [O], carboplatin [J], etoposide [E], and cyclophosphamide [C]) induction chemotherapy, the ecDNA copy number increased to several hundred copies and remained high throughout additional rounds of chemotherapy given prior to and until the patient’s death (**Fig. 5**). Structural analysis of the identified ecDNA with SAVANA and *de novo* genome assembly confirmed the circular nature of the ecDNA molecule, which was 773,976bp in size (assembled into 4 contigs of 719, 51,891, 229,985 and 491,381bp; Timepoint 2 in **Fig. 5**). Notably, the largest contig encompassed the four breakpoints generating the two structural variants underpinning the circularization of the ecDNA. Strikingly, we were able to detect the same ecDNA molecule and associated breakpoints in all cfDNA samples from this patient during disease progression (**Supplementary Fig. 17**), indicating that the ecDNA molecule remained stable throughout treatment, a phenomenon that was also observed in neuroblastoma patient **R0171** (**Supplementary Fig. 18**).

**Figure 5.**
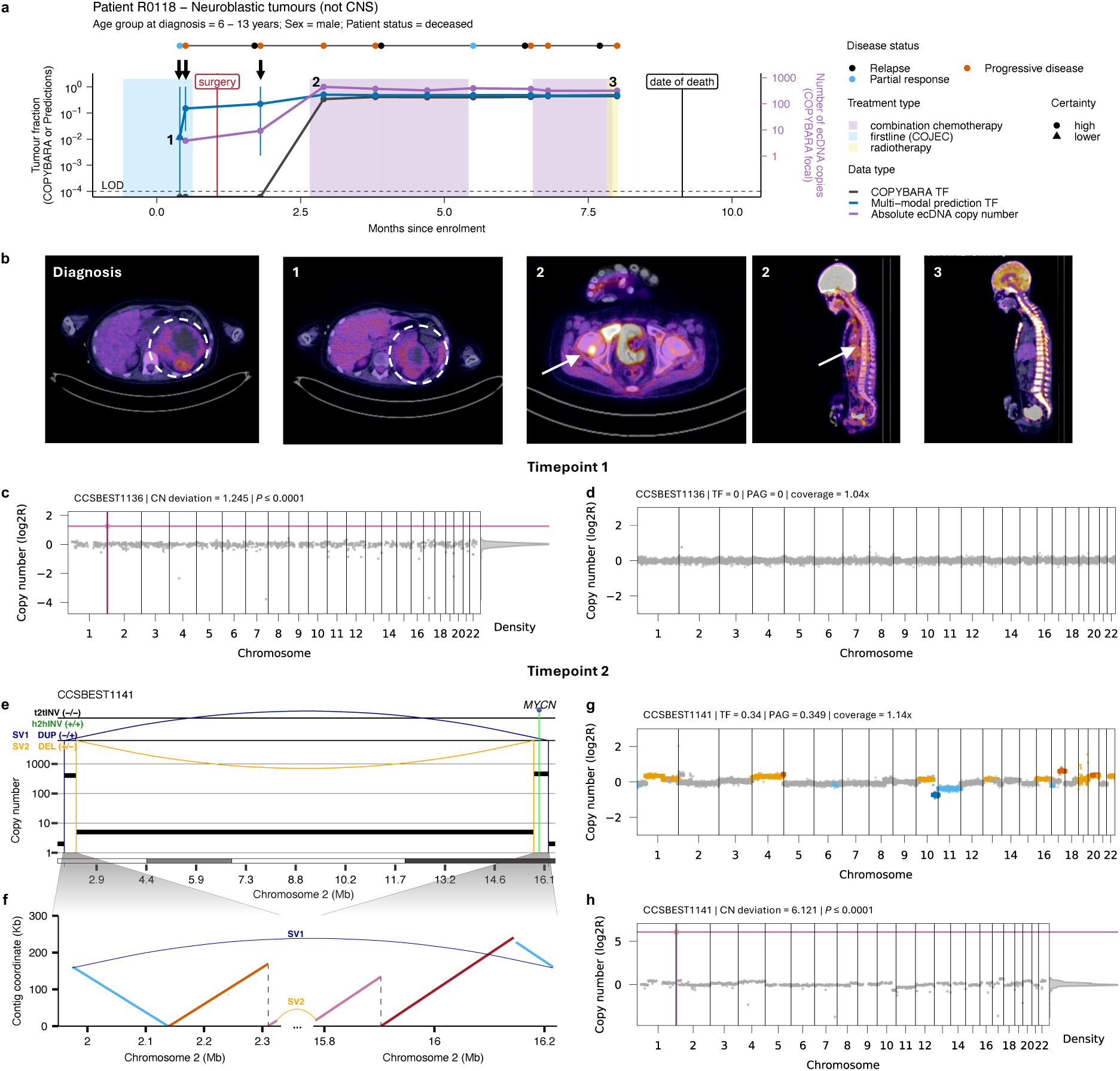
Longitudinal tracking of tumour burden using ecDNA and multimodal CP for neuroblastoma Patient R0118. **(a)** Longitudinal disease trajectory for a male neuroblastoma patient (R0118) showing the TF predictions over time using either COPYBARA (genome-wide copy number analysis; dark grey) or predictions computed using multimodal CP (blue). 90% confidence intervals for TF predictions are shown in blue. Predictions for which the 90% confidence interval goes beyond the LOD (as indicated by the dashed line) are considered of low certainty and are depicted as triangles. The purple line shows the absolute ecDNA copy number detected using COPYBARA-focal informed by ecDNA-associated SV breakpoints. Treatment is indicated by coloured boxes. Disease status by RECIST 1.1 criteria are indicated for each cfDNA sample by coloured points. **(b)** Clinical images from Positron Emission Tomography (PET) with 2-deoxy-2-[18F]fluoro-D-glucose (FDG) scans for the timepoints marked with numbers. **(c)** Detection of the ecDNA containing *MYCN* (shown in pink) using COPYBARA-focal at timepoint 1. **(d)** Genome-wide copy number profile estimated using COPYBARA, which predicted a TF of 0 at timepoint 1. **(e)** Genomic rearrangement profile showing SCNAs and SVs detected using SAVANA showing the ecDNA amplifying *MYCN*. Absolute copy number data are represented by the black horizontal lines. SVs are represented by vertical lines. DEL: deletion-like rearrangement; DUP: duplication-like rearrangement; h2hINV: head-to-head inversion; t2tINV, tail-to-tail inversion. **(f)** Assembly of the ecDNA molecule using cfDNA nanopore sequencing reads. Each contig represented by a different colour. The ecDNA-associated SVs shown in **(e)** are shown by arched lines. **(g)** Genome-wide copy number profile generated using COPYBARA. Gains and amplifications are shown in yellow and orange, respectively. Losses and deletions are shown in light blue and dark blue, respectively. **(h)** Detection of the ecDNA containing *MYCN* (pink dot) at Timepoint 2 using COPYBARA-focal. CN deviation: copy number deviation; ecDNA: extrachromosomal DNA; LOD: limit of detection; PAG: percentage abnormal genome; SCNA: somatic copy number aberration; SV: structural variant; TF: tumour fraction.

In another patient, **G0050,** a complex ecDNA molecule generated by chromothripsis involving 18 SVs and amplifying *MYCN* was detected at the time of disease presentation (Timepoint 1 in **Fig. 6a-c; Supplementary Fig. 12a**). After receiving firstline chemotherapy with COJEC and radiotherapy, a complete radiological response (CR) was observed and ecDNA was not detectable (Timepoint 3 in **Fig. 6a-b**). However, at two later CR timepoints, we detected the same ecDNA molecule 4 months before clinical relapse, as well as positive ctDNA signal using the CP models (indicated by arrows in **Fig. 6a**). Notably, ecDNA molecules were detectable in samples with a sequencing depth below 0.1x, indicating the high sensitivity of ecDNA analysis for ctDNA detection. Similarly, in neuroblastoma patient **R0059**, we detected *MYCN* amplification in a simple ecDNA molecule 3 months before relapse (**Extended Data Figs. 6**). The ecDNA copy number fluctuated during treatment, correlating with clinical imaging data, TF predictions generated using CP and genome-wide SCNAs, which were detectable at timepoints with a high TF (**Extended Data Fig. 6**). By contrast, in patient **R0075**, *MYCN* amplification in ecDNA was the only detectable SCNA throughout chemotherapy treatment after a third relapse with bone metastasis, which was consistent with progressive disease (**Extended Data Fig. 7; Supplementary Fig. 12b**).

**Figure 6.**
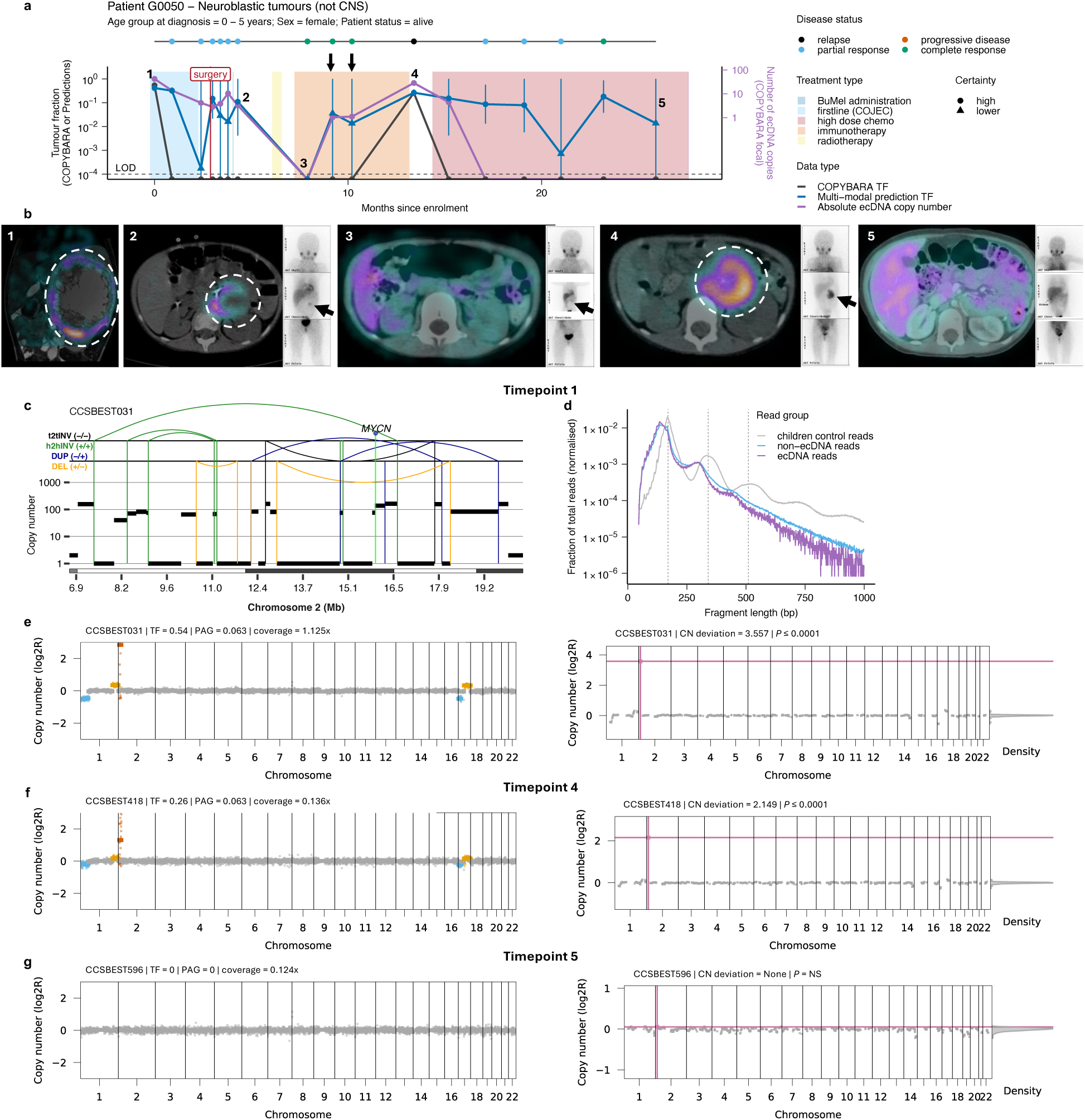
Longitudinal tracking of tumour burden using ecDNA and multimodal CP for neuroblastoma Patient G0050. **(a)** Longitudinal disease trajectory for a female neuroblastoma patient (G0050) showing the TF predictions over time using either COPYBARA (genome-wide copy number analysis; dark grey) or predictions computed using multimodal CP (blue). 90% confidence intervals for TF predictions are shown in blue. Predictions for which the 90% confidence interval goes beyond the LOD (as indicated by the dashed line) are considered of low certainty and are depicted as triangles. The purple line shows the absolute ecDNA copy number detected using COPYBARA-focal informed by ecDNA-associated SV breakpoints. Treatment is indicated by coloured boxes. Disease status by RECIST 1.1 criteria are indicated for each cfDNA sample by coloured points. **(b)** Clinical images from metaiodobenzylguanidine (MIBG) scans taken at the timepoints marked with numbers. **(c)** Genomic rearrangement profile showing SCNAs and SVs detected using SAVANA showing the ecDNA amplifying *MYCN*. Absolute copy number data are represented by the black horizontal lines. SVs are represented by vertical lines. DEL: deletion-like rearrangement; DUP: duplication-like rearrangement; h2hINV: head-to-head inversion; t2tINV, tail-to-tail inversion. **(d)** Fragment size profile of cfDNA reads mapping to ecDNA (purple) and non-ecDNA (blue) regions for sample CCSBEST031. The fragmentation profile of cancer-free paediatric cfDNA samples is shown in grey for comparison. **(e-g)** Genome-wide COPYBARA copy number profile (left) and detection of the ecDNA amplifying *MYCN* (pink dot) using COPYBARA-focal (right) for timepoints 1, 5 and 7. Gains and amplifications are shown in yellow and orange, respectively. Losses and deletions are shown in light blue and dark blue, respectively. CN deviation: copy number deviation; ecDNA: extrachromosomal DNA; LOD: limit of detection; PAG: percentage abnormal genome; SCNA: somatic copy number aberration; SV: structural variant; TF: tumour fraction.

The ecDNA copy number detected in cfDNA was also informative of favourable treatment response. For example, in patient **R0087**, diagnosed with metastatic neuroblastoma, we detected a complex ecDNA harbouring *MYCN* in two samples both collected at the time of diagnosis (**Supplementary Fig. 19**). However, ecDNA was not detectable in any of the 7 subsequent longitudinal samples collected during a complete clinical response spanning >15 months post-surgery (**Extended Data Fig. 8**). Similarly, in patient **R0135** we detected and assembled a complex ecDNA molecule amplified to a total *MYCN* copy number of >500 at the time of diagnosis associated with characteristic chromosome 1p loss and amplification of 17q (**Fig. 3; Supplementary Fig. 13b**). The ecDNA copy number decreased in a continuous manner during firstline COJEC induction chemotherapy and remained undetectable in the 8 subsequent cfDNA samples collected over 7 months post-surgery, radiotherapy and immunotherapy treatment (**Extended Data Fig. 9**). However, ctDNA was detectable by the multimodal models at PD timepoints, indicating that ecDNA detection and multimodal data integration provide complementary information that enhances ctDNA detection.

SIMMA also enabled monitoring of disease evolution for tumours not driven by ecDNA. For example, in a medulloblastoma patient with bone metastases, **R0119,** multimodal CP predicted a TF of 0.87 in plasma cfDNA. Partial responses during treatment were reflected in cfDNA TF values and correlated with metabolic treatment responses (**Extended Data Fig. 10**), indicating that SIMMA permits detection of extracranial metastatic disease in plasma from patients with paediatric brain tumours.

Together, these results show that both TF predictions based on multimodal data and the reliable detection of simple and complex ecDNA molecules in cfDNA enables monitoring of treatment response, analysis of disease evolution and the early detection of relapse in patients with cancer.

## Discussion

SIMMA is a novel single-molecule framework for the scalable analysis of cfDNA through both multimodal genome-wide and targeted sequencing designed to be compatible with low-input liquid biopsy samples in both tumour-agnostic and tumour-informed settings. Importantly, SIMMA enables multimodal cfDNA analysis from a single assay, which may be particularly advantageous in settings where cfDNA input or infrastructure are insufficient to support multiple dedicated assays and to expedite the turnaround time of results. While in this study we have relied on nanopore sequencing, SIMMA is compatible with other approaches for multimodal DNA profiling, with implications for cancer and other diseases^27–29^.

Analysis of a large collection of plasma and CSF cfDNA samples from patients with paediatric cancers with molecularly diverse solid tumour types supports the clinical utility of SIMMA for tumour type classification, relapse detection, disease monitoring and the identification of cancer drivers. As opposed to short-read panel and low-pass WGS of cfDNA^31^, the multimodal data integration enabled by SIMMA permits ctDNA detection in plasma from paediatric brain tumour patients and improves sensitivity for tumour-agnostic relapse detection. Therefore, SIMMA broadens the cancer types for which liquid biopsy analysis can deliver clinically useful results.

Unbiased sequencing of short and long cfDNA fragments allowed us to analyse repetitive genomic regions recalcitrant to short-read cfDNA profiling. Our data show that hypomethylation at satellite repeats provides a strong and specific signal to identify ctDNA, and that long DNA fragments detected in plasma are of cancer origin, thus revealing novel cancer signals in cfDNA to be exploited for ctDNA detection.

Assessment of the trustworthiness of model predictions for individual patients remains a critical barrier to the adoption of AI in clinical settings^50^. Here, we show that multimodal data integration using CP provides well-calibrated confidence intervals for individual predictions in tumour-agnostic settings. Of particular clinical relevance, the size of the confidence intervals permits the identification of unreliable predictions and the quantification of the LOD of models in a data-driven manner. Critically, the sequencing depth we achieved for clinical samples was constrained by the amount of cfDNA remaining after short-read analyses generated by the SMPaeds programme^31^, thereby limiting the attainable LOD. Similarly, the number of patients and tumour types included in the training set was also limited, which constrains the models ability to generalise, especially for features with strong cancer-type specificity, such as methylation. However, as larger quantities of cfDNA can typically be collected from both paediatric and adult patients with cancer, we anticipate improved sensitivity for models trained on cohorts with greater cfDNA input and more diverse tumour types. Because CP can be implemented with any AI algorithm, including deep learning architectures^54^, the framework we present here could be further extended to incorporate other data modalities detected in plasma, e.g., proteomics^55^, or clinical data sets, including imaging or electronic health records, thereby facilitating the application of multimodal AI data integration with uncertainty quantification for cancer detection and monitoring.

Complex genomic rearrangements, such as ecDNA and chromothripsis, are pervasive across the most aggressive paediatric and adult cancers, and are mechanistically linked with malignant transformation^2,23,56^ and drug resistance^38–40^. Our results show that tumour progression and chemotherapy resistance in *MYCN-*amplified neuroblastoma^57^ correlated with ecDNA copy number in cfDNA. We harnessed ecDNA-specific somatic rearrangements detected in cfDNA to inform ecDNA detection for minimal residual disease monitoring, which showed that ecDNA can be detected months before clinical relapse. Although the prevalence of ecDNA in non-neoplastic tissues remains unknown, we did not detect ecDNA in cancer-free controls or in housekeeping genes in cfDNA from patients with cancer, indicating that ecDNA provides high specificity for ctDNA detection. Beyond ecDNA, SIMMA may enable MRD monitoring through the detection of rearrangements without requiring matched tumour whole-genome sequencing data, which are rarely available in most cancer hospitals worldwide. Collectively, our results unlock the potential to harness ecDNA and other complex rearrangements detected in cfDNA as liquid biopsy biomarkers for clinical trials, cancer detection and MRD monitoring in both tumour-agnostic and tumour-informed settings, with broad implications for paediatric and adult malignancies.

## Methods

### Human subjects and patient recruitment

Patients with paediatric cancers were recruited to the Stratified Medicine Paediatrics (SMPaeds) study, as previously described^31^ (IRAS: 246557/264925, doi: 10.1186/ISRCTN21731605). SMPaeds is a UK precision medicine program, which since 2019 has recruited patients with paediatric-type tumours at the time of relapse or treatment-refractory disease from across 20 paediatric oncology centres across England, Scotland, Wales and Northern Ireland. In addition, patients with paediatric cancers from whom longitudinal blood samples were collected were recruited to the local Liquid Biopsy Trial (IRAS: 281408) at the Great Ormond Street and Royal Marsden hospitals (London, UK). Plasma samples from cancer-free individuals were collected in the Neurosurgery Department at Hospital de Santa Maria, Unidade Local de Saúde de Santa Maria (ULSSM) and stored at Biobanco of Gulbenkian Institute for Molecular Medicine, Lisbon Academic Medical Center (Lisbon, Portugal). Ethical approval was obtained from the Ethics Committee of ULSSM (Ref. No.: 367/18). Written informed consent from patient or legal guardian (in the case of minors, with age-appropriate assent where possible) was obtained from all patients recruited to these studies before study participation in accordance with the European and National Ethical Regulation (law 12/2005). Patients did not receive financial compensation for donating samples. In addition, we analysed 35 commercially available control samples from Cambridge Bioscience from 19-58 year old patients (median age: 29 years; Cat No.: PLSSKF2ED, PLSSKF2LH, and PLSSKF2SH; **Supplementary Table 5**).

### Sample collection and DNA extraction

Whole blood samples were collected into cfDNA blood collection tubes (Streck), and plasma was separated from blood within seven days of venepuncture by the Royal Marsden Hospital Laboratory team using double centrifugation. Specifically, whole blood was spun at 1600 g for 10 min. The resulting plasma fraction was collected and spun again at 1600 g for another 10 mins, subsequently aliquoted into cryovials, and stored at −80°C until further use. cfDNA was extracted from plasma samples using the QIAamp DSP Circulating Nucleic Acid kit (Cat. No.: 61504, Qiagen) and eluted in 40µl of molecular grade nuclease-free water into Eppendorf DNA LoBind® Tubes (Eppendorf).

### Single-molecule integrative multimodal analysis (SIMMA) of cfDNA

cfDNA samples were sequenced on the ONT PromethION nanopore sequencing platform unless otherwise stated. Library preparation was performed as follows and as illustrated in **Fig. 1a**:

#### DNA repair and end prep

Following cfDNA extraction, DNA was end-repaired by mixing 0.875µl of NEBNext FFPE DNA Repair Buffer, 0.875µl Ultra II End-Prep Reaction buffer (NEB, Cat. No.: E7546 component E7647AVIAL), 0.75µl Ultra II End-Prep Enzyme Mix (NEB, Cat. No.:E7546 component E7646AVIAL) and 0.5µl NEBNext FFPE DNA Repair Mix (NEB, Cat. No.: M6630) with 12µl of extracted cfDNA. Following gentle pipette mixing and spinning down, the reaction was incubated on a thermal cycler at 20°C for 12 mins, followed by a further incubation at 65°C for 12 mins. Next, a 1:1.2 AMPure XP (Beckman Coulter, Inc., Cat. No.:A63881) bead clean-up was performed with two 80% EtOH washes, and the end-prepped cfDNA was eluted into 40µl of molecular grade nuclease-free water to ensure sufficient sample volume for simultaneous WGS and additional targeted sequencing. For low input samples with <10ng of starting cfDNA, the elution volume was lowered to 25µl.

#### Native barcode ligation

Following DNA end-repair, 22.5µl of end-repaired DNA was mixed with 2.5µl of a unique barcode (NB01-96 from the Native Barcoding Kit 96 V14 SQK-NBD114.96) and 25µl of Blunt/TA Ligase Master Mix (NEB, Cat. No.: M0367) and incubated for 30 mins at room temperature. 5µl of EDTA (0.5M) was added to terminate the ligation reaction. Importantly, the remainder of end-repaired DNA (∼17.5µl) was set aside (and if required stored at −80°C) for targeted sequencing as described below. Barcoded samples were pooled together. Multiplexing of 16 cfDNA samples was performed unless otherwise stated. Subsequently, a 1:1 AMPure XP (Beckman Coulter, Inc., Cat. No.: A63881) bead clean up with two 80% EtOH washes was performed and multiplexed and cleaned samples were eluted into 35µl of molecular grade nuclease-free water.

#### Adapter ligation

Next, 35µl of barcoded and pooled DNA were mixed with 5µl of Native Adapter, 10µl of NEBNext Quick Ligation Reaction Buffer (5X; NEB, Cat. No.: E6056 component E6058AVIAL), and 5µl of Quick T4 DNA Ligase (NEB, Cat. No.: E6056 component E6057AVIAL) and incubated for 30-40 mins at room temperature. Subsequently, adapter-ligated DNA was cleaned up with AMPure XP (Beckman Coulter, Inc., Cat. No.: A63881) beads (1:1.2), resuspended to wash twice with Short Fragment Buffer (SFB), and eluted into 25µl of Elution Buffer.

#### Targeted sequencing

Optionally, the remaining end-repaired DNA (∼12-17.5µl) set aside during whole genome sequencing library preparation was used for targeted sequencing using e.g. PCR-based panels, such as the Oncomine™ Pan-Cancer Cell-Free panel, or capture-based panels, such as the xGen^TM^ Hyb Capture Panel (Integrated DNA Technologies, IDT). While multiple panels were tested and successfully run, for the main analysis of clinical samples in this study, we developed a custom IDT xGen^TM^ Hyb Capture Panel targeting 76 genes commonly mutated in paediatric solid tumours by extending a previously published gene panel^31,32^. The genes included in the panel were: *ACVR1, AKT1, ALK, AMER1, ARID1A, ATAD1, ATM, ATRX, BCOR, BRAF, CCND1, CCND2, CCNE1, CDK4, CDK6, CDKN2A, CDKN2B, CPM, CREBBP, CTNNB1, CYP27B1, EGFR, ERBB2, EWSR1, EZH2, FBXW7, FGFR1, FGFR2, FGFR3, FGFR4, GLI2, H3F3A, HIST1H3A, HIST1H3B, HIST1H3C, HIST2H3C, HRAS, IDH1, IDH2, IGF1R, KIT, KRAS, LIN28B, MAP2K1, MDM2, MDM4, MET, METTL1, MMP11, MYC, MYCL, MYCN, MYOD1, NF1, NRAS, NSD3, PDGFRA, PIK3CA, PIK3R1, PPM1D, PTCH1, PTEN, PTPN11, RB1, SLC35E3, SMARCA4, SMARCB1, SMO, STAG2, SUFU, TERT, TP53, TSC1, TSC2, TSFM* and *WT1*. The remaining end-repaired DNA from the WGS workflow was prepared for capture hybridisation using the xGen^TM^ cfDNA & FFPE DNA Library Prep v2 MC kit, following the manufacturer’s instructions. Briefly, adapters containing 32 optimised, fixed UMI sequences (8bp in length) were ligated to the end-repaired DNA, which was subsequently amplified using 10-12 PCR cycles depending on input material. For example, 12 cycles of PCR were used for samples with an input DNA of 1ng, whereas only 10 cycles were used for samples yielding >10ng of DNA. During this step, sample index barcodes were also introduced. Following PCR cleanup, libraries were quality-checked and quantified, and 500ng of total amplified DNA were taken forward to the hybridisation step of our xGen^TM^ custom panel. A total of three samples were pooled together (500ng each; total 1.5µg for each capture reaction), and hybridisation and capture were performed using the xGen^TM^ Hyb and Wash Reagents v3 Kit following manufacturer’s instructions unless otherwise stated. Importantly, hybridisation was carried out for 2h before capturing target DNA, washing and PCR amplification. The post-capture PCR reaction was run using a total of 12 PCR cycles. Subsequently, DNA libraries were cleaned up, quantified and prepared for nanopore sequencing by ligating nanopore adaptors to the captured DNA using the Ligation sequencing DNA V14 kit (SQK-LSK114, ONT).

#### Multiplexed single-molecule sequencing

Barcoded samples for whole-genome sequencing were sequenced across two R10.4.1 PromethION flow cells by multiplexing 8-16 samples per library, and each library was split between the two flow cells. Samples prepared for panel sequencing were also sequenced on one PromethION flow cell, with three samples per capture and flow cell. In both settings, sequencing libraries were loaded at 50fmol and sequenced for 72hrs with the minimum read length set to 20bp. After loading, a flow cell would actively sequence undisturbed for the duration of its run. If a flow cell was underperforming after the first 24hrs of sequencing, a supplementary wash step was performed to improve pore occupancy and refresh running reagents using the Flow Cell Wash kit (EXP-WSH004, ONT).

#### Base-calling and processing of single-molecule sequencing data

Demultiplexing, base-calling and 5mC and 5hmC detection for the WGS nanopore data were performed using the super-accurate model of Guppy v6.5.7 (dna_r10.4.1_e8.2_400bps_modbases_5hmc_5mc) with a qscore filter of 10. Additionally, adaptors and barcodes were trimmed during base-calling. The resulting trimmed and demultiplexed sequencing reads were aligned to GRCh38 or T2T-CHM13v2.0 using minimap2 (v2.24)^58^ with parameters “-ax map-ont --MD”. Sequencing success and quality were assessed by estimating the total number of reads and sequencing depth per barcode/sample using mosdepth v0.3.3^59^, and by investigating read fragment sizes. Base-calling for IDT panel sequencing data was performed using Dorado v7.4.14 with the high-accuracy model (https://github.com/nanoporetech/dorado/).

#### Analysis of the efficiency of multiplexing for WGS

To test and validate SIMMA for WGS of cfDNA, DNA (NA12878) was extracted from the GM12878 lymphoblastoid cell line and sheared to 178bp using the LE220-plus Focused-Ultrasonicator (Covaris) to approximately mimic cfDNA. The extracted and sheared DNA was diluted to 5ng, 10ng, 20ng, 30ng, 40ng, 50ng, 100ng and 150ng in total volumes of 12µl each, and subjected to three different library preparation protocols: i) SIMMA, ii) the ONT Native Barcoding Library preparation using the Native Barcoding Kit 96 V14 protocol (August 2023), and iii) the ONT Native Barcoding Library preparation protocol for cfDNA (v4, November 2023). Samples of 5-150ng input DNA were pooled, 8-plexed and sequenced in triplicate on three independent flow cells for each library preparation protocol. Raw sequencing data were base-called using Guppy v6.5.7. Sequencing reads were aligned to GRCh38 using minimap2 as described above, and assessed for genome-wide sequencing output using mosdepth v0.3.3^59^.

### Sensitivity and specificity analysis for mutation detection using contrived samples

For the development and validation of low-input cfDNA panel sequencing of end-repaired DNA, we utilised SeraSeq® ctDNA Complete^TM^ Reference Material with VAF values for the mutations of 5%, 1%, 0.5% and 0.1% (0710-0669, 0710-0671, 0710-0672, and 0710-0673, respectively, Seracare). SeraSeq samples were subjected to our SIMMA workflow to obtain the end-repaired DNA side product. Subsequently, 10ng of end-repaired DNA were used as input material for target panel sequencing using our custom IDT xGen^TM^ Hyb Capture Panel as outlined above. Each SeraSeq reference sample was analysed in triplicate. In addition, we generated a dilution series to obtain additional SeraSeq VAF samples at 0.05%, 0.025% and 0.01%. For this purpose, the SeraSeq® ctDNA Complete^TM^ Reference Material was diluted using sheared genomic background DNA (DNA24385) isolated from the GM24385 B-lymphocyte cell line. Diluted SeraSeq samples were processed using the same workflow used to prepare the original SeraSeq samples and were sequenced and analysed in triplicate.

### Downstream analysis of single-molecule cfDNA sequencing data

#### Estimation of sequencing depth

Sequencing depth of whole-genome sequencing and panel sequencing data was assessed using mosdepth v0.3.3^59^ with “MOSDEPTH_PRECISION=5”. For panel sequencing data, the amplicon target bed file was used to compute the sequencing depths for each target region.

#### Genome-wide analysis of DNA methylation

Methylation signal for 5mC and 5hmC modifications was extracted from BAM files using modkit v0.3.2. Read-level statistics across the whole BAM file were estimated using “modkit summary” with the “--only-mapped” parameter. bedMethyl tables with summary counts of modified and unmodified bases across reference CpG dinucleotides were generated using “modkit pileup” with the “--cpg” parameter providing the GRCh38 or T2T-CHM13v2.0 genome as reference. The resulting bedMethyl table was subsequently used to compute the fraction of unmodified vs modified (5mC and 5hmC) cytosines across 500Kbp non-overlapping bins. Additionally, per-read methylation modifications were extracted using “modkit extract” with the “--cpg” parameter. The output table was then used to compute the total number of modified (5mC or 5hmC) and unmodified CpG dinucleotides for each individual read, along with its total read length, 4-base start and end motif, and genomic alignment position.

#### Analysis of DNA methylation across repetitive regions in the human genome

To assess methylation signal across repetitive regions of the human genome, the “modkit stats” functionality of modkit v0.5.0 was used. Repetitive regions for LINEs, rRNA, satellites and SINEs were extracted from Repeatmasker tracks downloaded from the USCS Genome Browser^60^ for GRCh38 or T2T-CHM13v2.0. The mean fractions of 5mC and 5hmC modifications were subsequently computed across different repeat classes, families and subfamilies. To investigate methylation signal across satellites at centromeres and telomeres, sequencing reads aligned to the T2T-CHM13v2.0 genome were used.

#### Fragmentomics analysis

Fragment size distributions were estimated by directly computing the read length of individual reads for each sample using SAMtools as follows: “samtools view ${bam} | cut -f 10 | perl -ne ‘chomp;print length($_). “\n“’ | sort | uniq -c”.

For nucleosome fraction and fragment size selection analyses using COPYBARA, reads were stratified by fragment length as follows. Reads of 90–240bp were classified as mono-nucleosomal, and those between 90–150bp, considered shorter than the expected modal length of 167bp for mono-nucleosomal cfDNA fragments from cancer-free controls, were classified as short mono-nucleosomal. Similarly, reads of 250-400bp and 450-600bp were classified as di- and tri-nucleosomal, respectively, and those between 250-325bp and 450-525bp were classified as short di and tri-nucleosomal. Fragments with read lengths >1000 bp were classified as “ultralong.”

#### Detection of somatic SNVs in targeted cfDNA sequencing data

Following Dorado basecalling, fastq files were demultiplexed based on their i5 and i7 barcodes using cutadapt v5.1^61^ with the following parameters “--action=none -e 0.2” and all remaining adapters were trimmed from reads by detecting P5 and P7 adapter sequences using cutadapt v5.1 with “--action=trim -n 2 –revcomp -e 0.2”. Subsequently, demultiplexed and trimmed fastq files were converted to unaligned BAM files using picard FastqToSam and for each read UMIs were extracted and added to the BAM files using pysam v0.20.0 and python v3.9.7 to the read tags RX ZA and ZB. The resulting UMI-tagged BAM files were then aligned to GRCh38 using winnowmap v2.03, potential sequencing errors in UMIs were corrected using the fgbio v3.0.0 (https://github.com/fulcrumgenomics/fgbio) functionality CorrectUmis with “--max-mismatches=3 --min-distance=1”, and reads were grouped into read families based on their UMIs and genomic positions using the fgbio v3.0.0 functionality GroupReadsByUmi with “--strategy=adjacency --edits=0 --min-map-q=5”. Finally, consensus reads were called on read families with ≥2 reads using a family-size-dependent strategy to balance read retention and error suppression. Specifically, for families containing two reads, a conservative pairwise alignment approach was applied using the Biopython package v1.85^62^, in which reads were globally aligned and only bases that agreed in both position and nucleotide identity were retained in the consensus sequence, while all discrepant or indel-associated positions were masked as ambiguous (N; **Supplementary Figs. 10-11**). This ensured that uncertainty from minimal read support was explicitly represented without introducing false substitutions. For families containing ≥3 reads, multiple sequence alignment and consensus calling were performed using SPOA v4.1.5, which robustly infers consensus sequences from higher-depth read families^63^. Consensus sequences were subsequently aligned to GRCh38 using minimap2 (v2.24)^58^ with parameters “-ax map-ont --MD”.

### Detection of somatic copy number aberrations using COPYBARA

COPYBARA is a Python algorithm designed to detect large and focal somatic copy number aberrations (SCNAs) and infer tumour ploidy and tumour fraction using low-coverage cfDNA WGS data. As summarised in **Supplementary Fig. 4**, COPYBARA consists of the following steps:

#### Binning of the reference genome

First, COPYBARA bins the reference genome into equally sized, non-overlapping bins (by default 500Kbp in size) and computes the number of unknown bases, the proportion of bases overlapping with blacklisted regions if provided, as well as the GC content for each bin.

#### Read counting

Subsequently, COPYBARA counts the number of primary alignments with a minimum read mapping quality ≥ 5 for each bin. Read counting is performed across the same bins for a matched normal sample if available. Bins overlapping with blacklisted regions or with more than 75% of unknown bases are excluded. Additionally, bins containing no reads are not considered for further analysis. Next, COPYBARA assess and corrects GC bias using a LOESS-based approach and converts read counts to log2 ratios (log2R) by normalising read counts using a matched normal if provided. Alternatively, if no matched normal sample is available, the user can provide a panel of normals (PoN). This can be generated by using the built-in “copybara pon” functionality of COPYBARA, which aggregates read counts across multiple normal samples. In this study, all cfDNA samples from paediatric patients with cancer were analysed using a PoN generated using 16 control plasma samples collected from children without cancer. If neither a matched normal nor a PoN are available, read counts are self-normalised using the median read counts of the sample being analysed.

#### Smoothening and segmentation

Following GC correction and normalisation, log2 read counts are smoothened using a trimmed mean approach to reduce noise and are Anscombe square-root transformed to stabilise variance before segmentation. Copy number (CN) segments and breakpoints are then detected with circular binary segmentation (CBS)^64,65^ using 1,000 permutations. To minimise the risk of over-segmentation, adjacent segments from the CBS step are merged if their median log2 read count values are not above a user-defined quantile of all absolute median differences. By default, only samples with a sequencing depth <0.01x undergo segment merging with a quantile threshold of 0.2.

#### Absolute copy number fitting and TF estimation

To improve the accuracy and efficiency of absolute copy number fitting, COPYBARA automatically estimates a suitable range of tumour TF values for each sample by searching for changes in copy number signal and estimating the copy number deviation as a proxy for TF. Specifically, COPYBARA analyses the distribution of relative copy number values across chromosomes and chromosome boundaries to identify multimodal patterns indicative of subclonal or aneuploid populations. Chromosomes and chromosome splits are screened for multimodality using the bimodality coefficient and kernel density estimation. The distances between density peaks are then used to estimate the copy number deviation and to define the minimum and maximum TF bounds, with a user-defined buffer (default=0.05) to accommodate for uncertainty. Chromosomes with high noise or known technical artefacts are excluded from this analysis. Subsequently, COPYBARA performs absolute copy number fitting, as previously described^43,66^. In brief, COPYBARA applies a grid search encompassing the previously defined sample-specific TF search space ({0.1*k* ∣ *k* ∈ [MinTF_tumour_.. MaxTF_tumour_]} range) and a ploidy search space of {0.01*k* ∣ *k* ∈ [1.7 .. 3.7]} range. For each TF-ploidy candidate pair COPYBARA computes a fitting error function by estimating the root mean squared deviation (RMSD) or alternatively the mean absolute deviation (MAD) between observed and integer copy number states across the whole genome. Candidate fits are filtered to exclude those with biologically implausible results, such as large proportions (>0.1) of segments with zero or negative copy number, or insufficient proportion of segments (<0.5) close to integer values. Finally, the best-fitting TF and ploidy are selected and used to compute absolute copy numbers for each segment as follows:

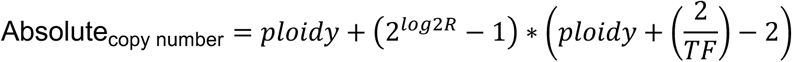

For cases where the TF was fitted to 0, the above formula simplifies to *Absolute_copy number_*= *ploidy* ∗ 2*^log2R^*.

After absolute copy number fitting, COPYBARA assigns each genomic segment to a copy number category based on its absolute copy number relative to the sample ploidy. To allow consistent categorisation across samples with different estimated ploidy levels, the categorisation is performed by first transforming absolute copy number values into ploidy-normalized copy number values using the following equation:

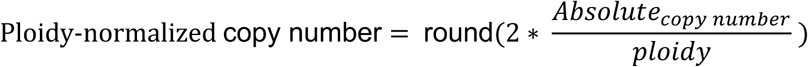

Segments are then classified into the following categories according to the estimated ploidy-normalized copy number values. Deletion: 0 or Absolute_copy number_ = 0; Loss: 1; Neutral: 2; Gain: 3; Amplification: ≥ 4.The percentage of abnormal genome (PAG) is then calculated as the fraction of the genome, weighted by segment length, that is not classified as neutral:

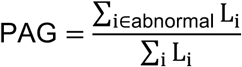

where *L_i_* is the length of the segment *i*, and “abnormal” includes all segments categorized as deletion, loss, gain, or amplification.

### Copy number analysis with size selection using COPYBARA

In addition to copy number analysis including all sequencing reads, COPYBARA provides the functionality to perform *in silico* size selection prior to copy number analysis and fitting by defining “--min_read_size” and “--max_read_size”. While all samples were primarily analysed without *in silico* size selection, additional analyses with size selection were performed on samples with sequencing depths ≥1x to only consider i) short mono-nucleosomal reads of 90-150bp in length, ii) short mono-, di- and tri-nucleosomal reads of 90-150bp, 250-325bp and 450-500bp in length, respectively, and iii) ultra-long fragments with read lengths >1000bp.

### Detection of ecDNA in cfDNA using COPYBARA-focal

The COPYBARA-focal module of COPYBARA enables the detection of focal amplifications and ecDNA in user-defined regions of interest (RoIs). For each RoI, COPYBARA-focal randomly samples 1,000 background regions of equivalent length to the RoI across the genome, excluding the RoI itself and 500Kbp downstream and upstream of the RoI. Background regions overlapping with blacklisted regions or containing >75% of unknown bases are excluded. Subsequently, COPYBARA-focal performs read counting and normalisation as described above for each RoI and all background regions. To assess whether the resulting log2R copy number observed in a given RoI significantly differs from the background copy number distribution (i.e., copy number deviation), a one-sample Student’s *t*-test is applied. The test statistic is calculated as:

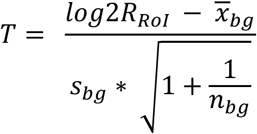

where 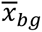 and 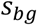 denote the mean and unbiased standard deviation of the background copy number values, respectively. The degrees of freedom were set to 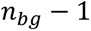, where 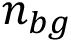 denotes the number of background regions included in the analysis. *P* values were obtained from the corresponding *t*-distribution under a greater-than (enrichment) alternative hypothesis. For RoIs passing the significance criterion, the magnitude of the CN deviation was estimated as described above. Additionally, if fitted TF and ploidy values from a genome-wide COPYBARA run are provided, the absolute copy number of the RoI is inferred as described above. Regions with an estimated absolute copy number ≥8 were annotated as putative ecDNA for further investigation. For cohort-wide analyses of ecDNA detection, P values were adjusted for multiple testing using the Benjamini–Hochberg false discovery rate (FDR) method. Only putative ecDNA results with FDR-adjusted *P* values < 0.5 were considered for downstream analyses.

### Reconstruction and assembly of ecDNA molecules using cfDNA WGS data

To detect SV breakpoints and copy number amplifications associated with ecDNA molecules in cfDNA, we used SAVANA v1.3.7^43^ in tumour-only mode. Following SV calling across the genome, complementary SCNA analysis was performed by priming the purity and ploidy search spaces for absolute copy number fitting using the TF and ploidy values obtained from COPYBARA, setting the bin size to 10Kbp and enforcing SV breakpoints to be incorporated into copy number segmentation. Subsequently, SV breakpoints initially discarded due to low tumour read support but supporting changes in copy number were rescued using “savana classify –cna_rescue”. Finally, ecDNA rearrangements and copy number profiles were visualised using the ReConPlot package^67^. In addition, we used CoRAL^44^ v1 and AmpliconArchitect^68^ v1.2.2 to detect ecDNA in cfDNA data. Before running AmpliconArchitect, seed regions were identified using COPYBARA with a bin size of 500Kbp and SAMtools (v1.19.2) with parameters “-Q 20”. The identified ecDNA regions were manually inspected for unexpectedly high read depth and the presence of split reads with soft-clipped bases at suspected breakpoints indicative of structural variants at the boundaries of ecDNA molecules. To assemble ecDNA molecules, we used Hifiasm v0.25.0-r726^69^. We used grid search to identify the optimal parameter values. Specifically, we considered the following parameter values: k-mer length {5, 10, 20} and --rl-cut {70, 100, 150, 200, 250, 300, 350}. All other parameters were set to default values and Hifiasm was run using the “-ont” option.

### ecDNA analysis in serial cfDNA samples

For patients with serial cfDNA samples and with evidence of ecDNA with well-defined genomic coordinates estimated based on soft-clipped reads detected by SAVANA, COPYBARA-focal was run across serial samples available to track ecDNA signal over time. To determine the limit of detection of COPYBARA-focal for ecDNA analysis, we generated in silico admixtures using sequencing reads from 22 paediatric cancer-free control samples and one ecDNA-positive cfDNA sample. Next, COPYBARA-focal was run on each dilution in a tumour-informed manner. To evaluate specificity, we applied the same analysis to 1,000 admixtures generated using cancer-free controls.

### Data encodings for multimodal integration using Conformal Prediction

To train CP models, we encoded each cfDNA sample by concatenating the following sets of covariates: (1) Genome-wide log2R values estimated using COPYBARA for non-overlapping 500Kbp bins; (2) fraction of 5mC and 5hmC at CpG sites across the genome; (3) fraction of 5mC and 5hmC at CpG dinucleotides across the genome using 500Kbp bins; (4) fraction of 5mC and 5hmC at CpG dinucleotides across all LINE, SINE, rRNA and satellite repeats; (4) the model fragment lengths for mono-, di- and tri-nucleosomal fragments; (5) fraction of fragments shorter than the expected modal length for mono-, di- and tri-nucleosomal fragments in cfDNA from cancer-free individuals; and (6) fraction of methylated 5mC and 5hmC at CpG dinucleotides across fragment sizes ranging from 60–1000 bp, calculated in 10 bp increments for each of the 16 two-base end motifs.

### Personalized tumour fraction prediction with uncertainty quantification

SIMMA relies on Conformal Prediction and Random Forest (RF) regression to predict the TF of a given cfDNA sample with uncertainty quantification using as covariates diverse feature sets derived from cfDNA nanopore WGS data. To establish a training set, we generated *in silico* tumour admixtures with TF values in the 10^−1^-10^−6^ range by combining the sequencing reads from paediatric cancer-free controls with sequencing reads sampled without replacement from one cfDNA sample from a set of 32 patients with cancer with TF values >0.46 as determined by COPYBARA. To predict the TF of a given cfDNA sample, we trained RF regression models using Scikit-Learn^70^ and default parameter values, except for the number of trees, which was set to 500. To generate predictions for each patient, we trained a leave-one-patient-out (LOPO) RF model using the *in silico* admixtures from all other patients to ensure that no data from the patient being analysed was used for model training. This LOPO strategy allows us to assess the predictive power that the models would have on new patients in a prospective clinical setting. To ensure that the performance of our models was not driven by spurious correlations, we performed Y-scrambling experiments by training RF models on randomized TF values.

We implemented CP to provide reliability estimates for each predicted TF value in the form of confidence intervals. In brief, the training data was divided into a proper training set (70% of instances) and a calibration set (30%). We trained a RF model on the proper training set using default parameter values, which was then applied to the calibration set. We next computed the nonconformity score, *⍺*, for each instance in the calibration set as follows:

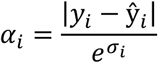

where *yi* is the simulated TF for the *i*th instance in the calibration set, and ŷ*i* and 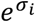 are the mean and standard deviation of the predicted TF for the *i*th instance across base learners (i.e., trees in the RF model). Dividing by 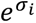 results in narrower confidence intervals for instances for which the predictions across base learners are more correlated, indicative of higher certainty. The list of non-conformity scores, α, for the calibration set is then sorted in increasing order. Next, the percentile corresponding to the confidence level desired is computed, .g., α_90_ for a confidence level of 90%, which guarantees that at least 90% of the predicted confidence intervals will contain the true value. Confidence intervals for new instances are computed as follows:

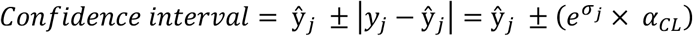

Where *y_j_* corresponds to e.g., the *j*th instance in the test set, and ŷ*_j_* and 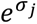 are the mean and standard deviation of the predicted TF for the *j*th instance in the test set by the RF model trained on the proper training set.

To estimate the LOD of CP models, we performed changepoint detection analysis^71^ to identify the size of the confidence interval corresponding to an increase in the size of the predicted confidence intervals as shown in Fig. 4e.

### Analysis of matched Illumina cfDNA sequencing data

Data from SMPaeds samples for which matching Illumina sequencing was available was reanalysed for comparison and validation purposes. To this end, bam files (previously aligned to the hg19 reference genome as described previously^31^) were converted to fastq files using SAMtools v1.19.2^72^ and aligned to GRCh38 using BWA-MEM2^73^ v2.2.1. Subsequently, ichorCNA^14^ v0.3.2 was used to estimate tumour fractions and detect SCNAs using 500Kbp bins. To ensure comparability between tools and analyses, a PoN was generated using the “createPanelOfNormals.R” functionality of ichorCNA, inputting the same 16 control plasma samples from cancer-free children that were used for the COPYBARA PoN described above.

### Statistical analysis and visualisation

All statistical analyses presented in this study were performed using R version 4.5.1. The level of significance was set at 0.05 unless otherwise stated. Multiple-testing correction was performed using the Benjamini–Hochberg procedure with an FDR threshold of 0.05. No statistical methods were used to predetermine sample sizes. Genomic rearrangement profiles were visualised using the R package ReConPlot^67^.

## Code availability

COPYBARA is available at https://github.com/cortes-ciriano-lab/copybara-cf.

## Data availability

Illumina cfDNA sequencing data for SMPaeds samples are available under controlled data access at the European Genome-Phenome Archive (EGA) under accession number: EGAS50000000549. The nanopore cfDNA sequencing data generated in this study will be available under controlled access at the EGA. The reference genome build GRCh38 was downloaded from https://hgdownload.soe.ucsc.edu/downloads.html. The genome assembly T2T-CHM13v2.0 was downloaded from https://www.ncbi.nlm.nih.gov/datasets/genome/GCF_009914755.1/.

## Author contributions

I.C.-C. designed, supervised, administered and obtained funding for the study. C.M.S., N.T., A.P., J.S., A.D.B., and I.C.-C. developed SIMMA. C.M.S. developed COPYBARA with the input of I.C.-C. C.M.S and I.C.-C. performed nanopore sequencing data analysis, multimodal analysis using conformal prediction, ecDNA detection analyses, and integration of genomics and clinical data sets, with support from S.Z. and F.M. N.T., A.P., J.S., A.D.B. performed nanopore sequencing experiments with input from C.M.S. and I.C.-C. D.H., C.H., M.N., E.P., R.C., C.C.F., L.V.M., F.S., L.R.P., P.B., T.S.J., M.H., D.H., S.G., P.A., and J.A. supervised sample and clinical data collection, sample shipments and cfDNA extractions. C.M.S., V.L., C.H., G.P., P.A. and S.G. collected and analysed clinical data. S.W.A., S.A.Y. and T.S.J. performed pathology review. G.A.A.B., A.V., L.V.M., T.S.J., M.H., D.H., S.G., P.A., J.A. and L.C. contributed samples, clinical data and clinical study coordination. A.G.R. and C.L. provided computational support. C.M.S. and I.C.-C. generated the figures, with input from N.T., A.P., J.S. and A.D.B. C.M.S. and I.C.-C. wrote the manuscript with input from all authors. All authors read and approved the final version of the manuscript.

## Acknowledgements

We thank the patients and their families for their participation in this study, which was primarily funded by Cancer Research UK (RCCFELCEA-May21\100001 awarded to I.C.-C.). C.M.S., S.Z., A.G.R. and I.C.-C. thank EMBL for funding. C.M.S. acknowledges support from the Marie Skłodowska-Curie Actions grant 101106070. A.D.B. received salary support from a Cancer Research UK Advanced Clinician Scientist Award (RCCACSANov16\23923) and MRC Senior Clinical Fellowship (MR/X006433/1). T.S.J. is grateful for funding from the Brain Tumour Charity (including the EVEREST Centre (GN-000707)), Olivia Hodson Cancer Fund, Cancer Research UK and the National Institute of Health Research. All research at Great Ormond Street Hospital NHS Foundation Trust and UCL Great Ormond Street Institute of Child Health is made possible by the NIHR Great Ormond Street Hospital Biomedical Research Centre. The views expressed are those of the author(s) and not necessarily those of the NHS, the NIHR or the Department of Health. We also acknowledge support from Cancer Research UK (CRUK; A24566), Children with Cancer UK (CWCUK; 17-235), Rosetrees Trust, and the Aoife’s Bubbles, Abbie’s Fund and Christopher’s Smile charities. We have received support from national networks, including the Experimental Cancer Medicine Center (ECMC) and the VIVO Biobank. We would like to thank Regan Barfoot, Leah Conner, Tracey Crowe and the wider Research Team at The Children’s and Young Persons Unit at The Royal Marsden Hospital, as well as the Royal Marsden Medical Research Centre. We would also like to thank Emily Baker and Rebekah Mantell for their dedication to discussing with the families of patients with paediatric cancers and for processing consent forms. We acknowledge funding from GIMM-CARE, funded by the European Union under grant agreement No. 101060102, co-funded by the Portuguese Government, the National Foundation for Science and Technology (FCT), ARICA, Jerónimo Martins, the Gulbenkian Institute for Molecular Medicine and Lisbon Academic Medical Centre. We thank the Biobanco of the Gulbenkian Institute for Molecular Medicine, Lisbon Academic Medical Center (Lisbon, Portugal) for their technical support. We thank the computational infrastructure provided by the EMBL-EBI and the Birmingham Environment for Academic Research (BEAR) at the University of Birmingham. We also thank all staff involved from the University of Birmingham Clinical Trials Unit and all external expert representatives on the study steering committee of SMPaeds. Some figures were generated using BioRender.com.

