## Supplementary Figures for "Single-molecule cfDNA sequencing establishes clinical utility for ecDNA monitoring and multimodal liquid biopsy analysis"

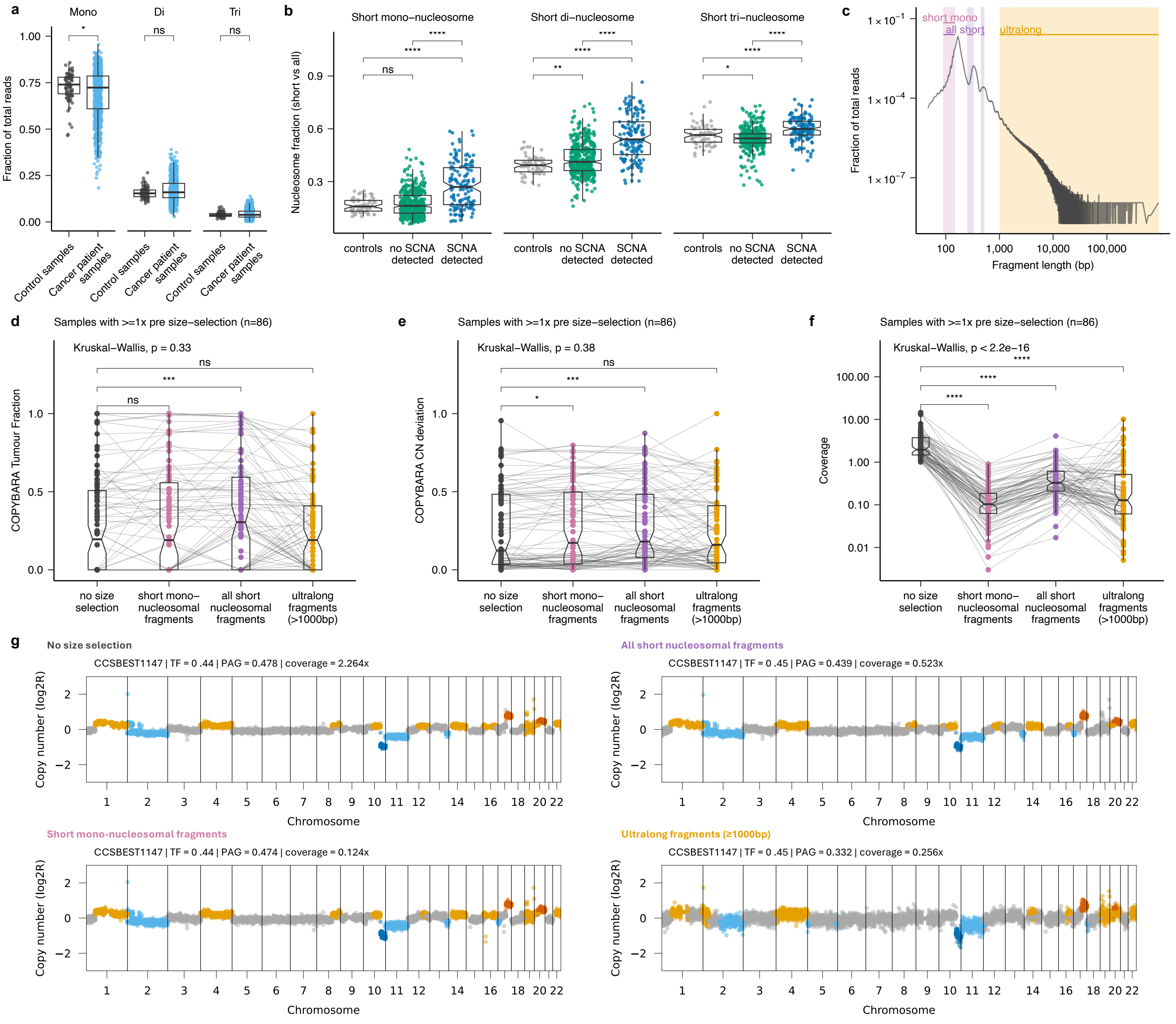


**Extended Data Figure 1. Fragmentomic feature analysis of cfDNA using SIMMA. (a)** Comparison of the fraction of reads detected as mono-, di-, or tri-nucleosomal fragments between cfDNA samples obtained from cancer-free control (grey) or paediatric cancer patients (light blue). **(b)** Fraction of short vs by all reads belonging to mono-, di- and tri-nucleosomal cfDNA fragments (**Methods**) from cfDNA samples obtained from cancer-free controls (grey), and paediatric cancer patients with SCNAs (blue), and without SCNAs (green) detected in cfDNA. **(c)** Example fragment size profile illustrating regions for COPYBARA size selection as follows: short mono-nucleosomal reads: 90-150bp short mono-, di- and tri-nucleosomal reads: 90-150bp, 250-325bp and 450-500bp, respectively, and ultralong fragments with read lengths >1000bp. Comparison of **(d)** COPYBARA TF, **(e)** COPYBARA CN deviation and **(f)** coverage obtained before (black) and after *in silico* size selection using COPYBARA for short mono-nucleosomal fragments (pink; 90-150bp), all short nucleosomal fragments (purple; short mono-, di- and tri-nucleosomal reads: 90-150bp, 250-325bp and 450-500bp) and ultralong fragments (>1000bp; orange). **(g)** Genome-wide COPYBARA copy number profile before and after COPYBARA *in silico* size selection for indicated fragment size for cfDNA sample CCSBEST1147. Gains and amplifications are shown in yellow and orange, respectively. Losses and deletions are shown in light blue and dark blue, respectively. Box plots in **a-b, d-f** show the median, first and third quartiles (boxes) and the whiskers encompass observations within 1.5× the interquartile range from the first and third quartiles.

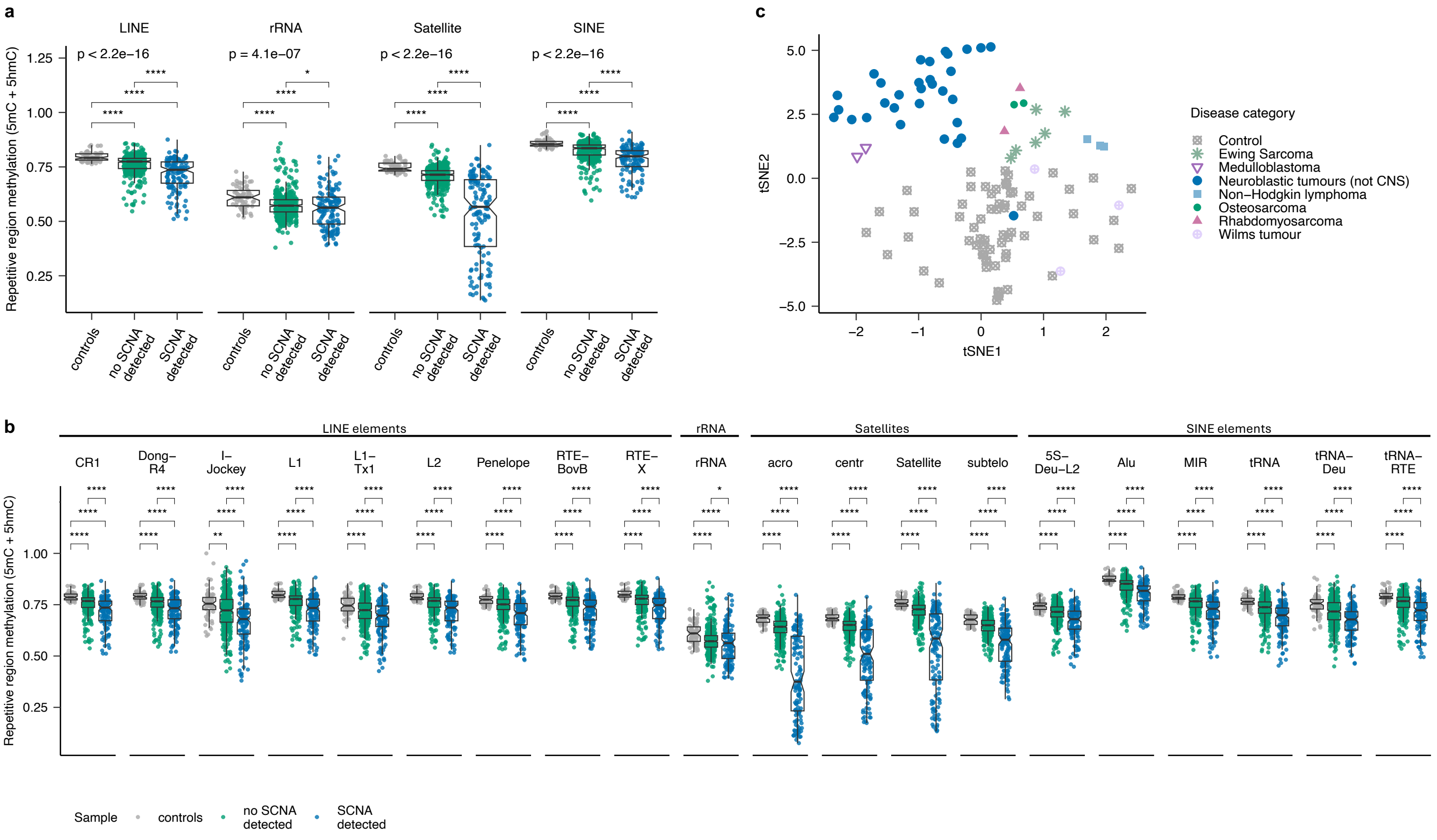


**Extended Data Figure 2. Genome-wide methylation analyses of cfDNA samples from paediatric cancer patients with solid tumours.**
(**a-b**) Fraction of 5mC and 5hmC at CpG dinucleotides across repetitive regions in plasma cfDNA samples from cancer-free controls (grey), paediatric cancer patients with SCNAs (blue), and without SCNAs (green) detected in cfDNA. (**c**) Clustering analysis of cfDNA genome-wide 5mC profiles of cancer-free controls and paediatric cancer patients. Box plots in **a-b** show the median, first and third quartiles (boxes) and the whiskers encompass observations within 1.5× the interquartile range from the first and third quartiles.

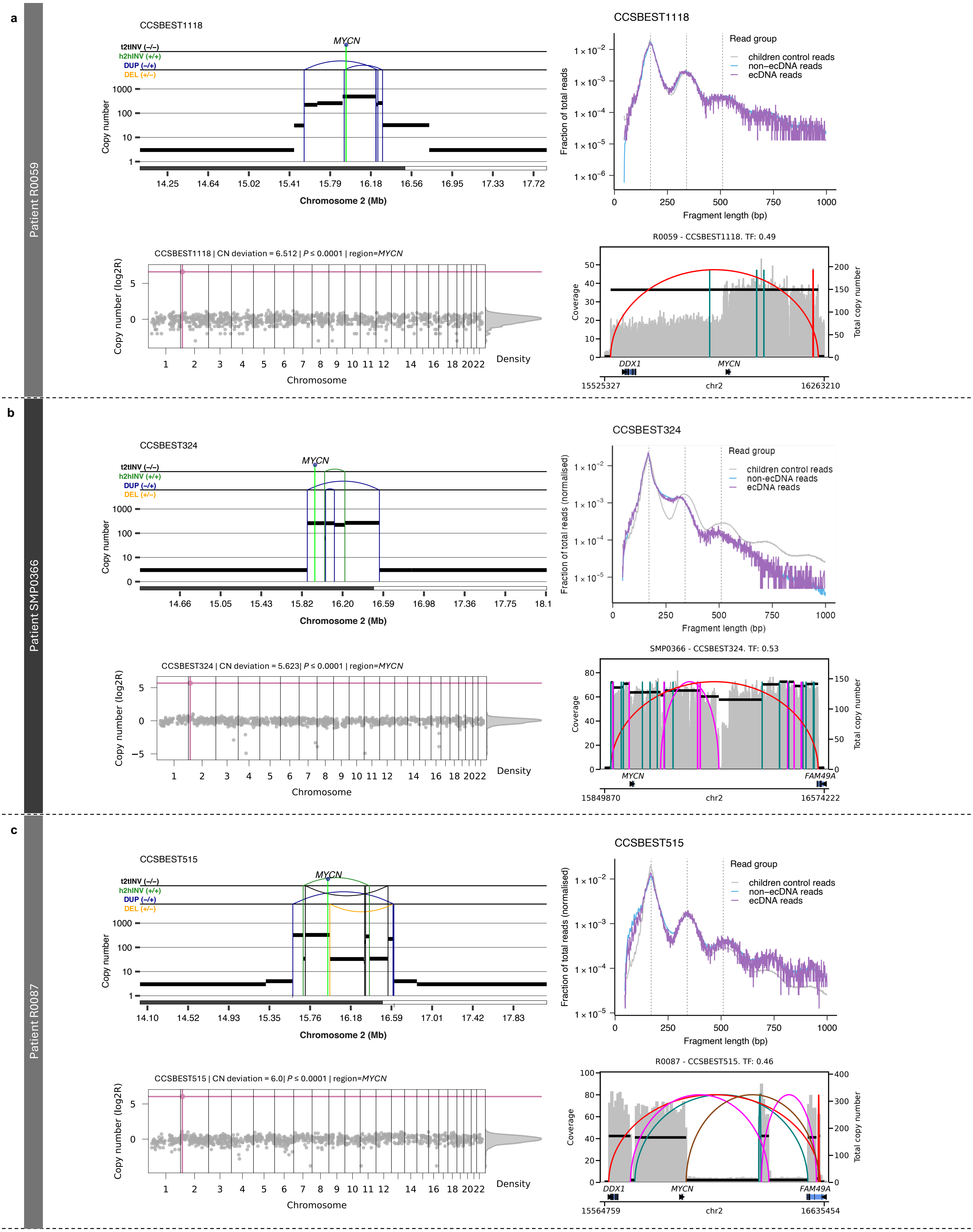


**Extended Data Figure 3. Detection and reconstruction of ecDNA in plasma cfDNA** from patients **(a)** R0059, **(b)** SMP0366 and **(c)** R0087.
For each patient, the **top left** panel depicts the genomic rearrangement profiles showing SCNAs and SVs detected using SAVANA showing the ecDNA amplifying *MYCN*. Absolute copy number data are represented by the black horizontal lines. SVs are represented by vertical lines. DEL: deletion-like rearrangement; DUP: duplication-like rearrangement; h2hINV: head-to-head inversion; t2tINV, tail-to-tail inversion. The **top right** panel shows the fragment size profile of cfDNA reads mapping to ecDNA (purple) and non-ecDNA (blue) regions. The fragmentation profile computed using cfDNA WGS data paediatric control samples is shown in grey for comparison. The **bottom left** panel shows the detection of ecDNA (pink dot) using COPYBARA-focal. Where available, the **bottom right** panel depicts the genomic copy number profile showing SCNAs and SVs detected using CoRAL.

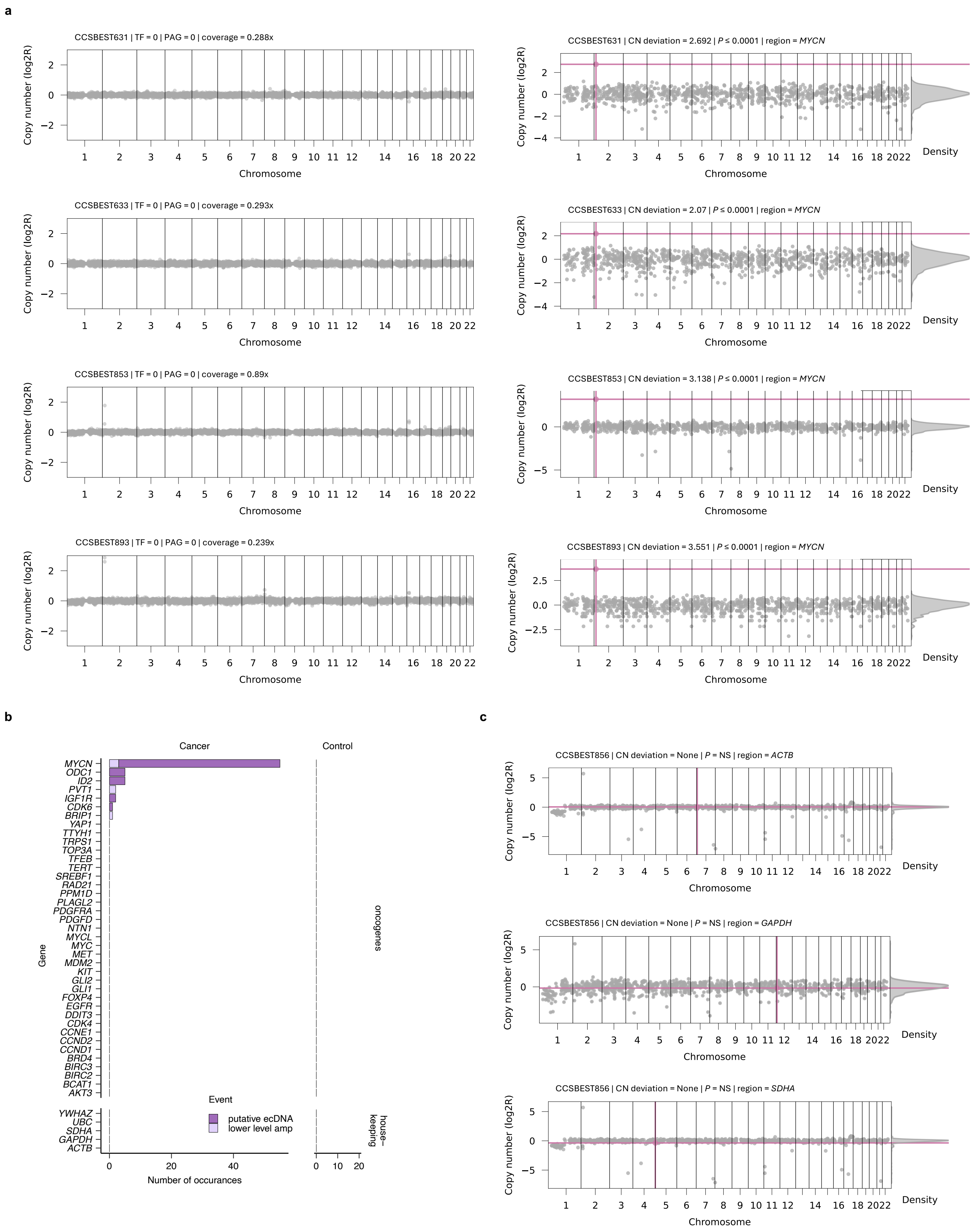


**Extended Data Figure 4.** (**a**) Examples of cfDNA samples with a no ctDNA signal detected based on SCNA analysis using COPYBARA in which COPYBARA-focal detected *MYCN* amplification in ecDNA. (**b**) Number of samples with ecDNA detected in oncogenes and housekeeping genes across cfDNA samples from paediatric cancer patients (n=65) and cancer-free controls (n=46) showing that COPYBARA-focal does not yield false positive ecDNA calls. (**c**) COPYBARA-focal results for three representative housekeeping genes for sample CCSBEST856 showing that no ecDNA was detected.

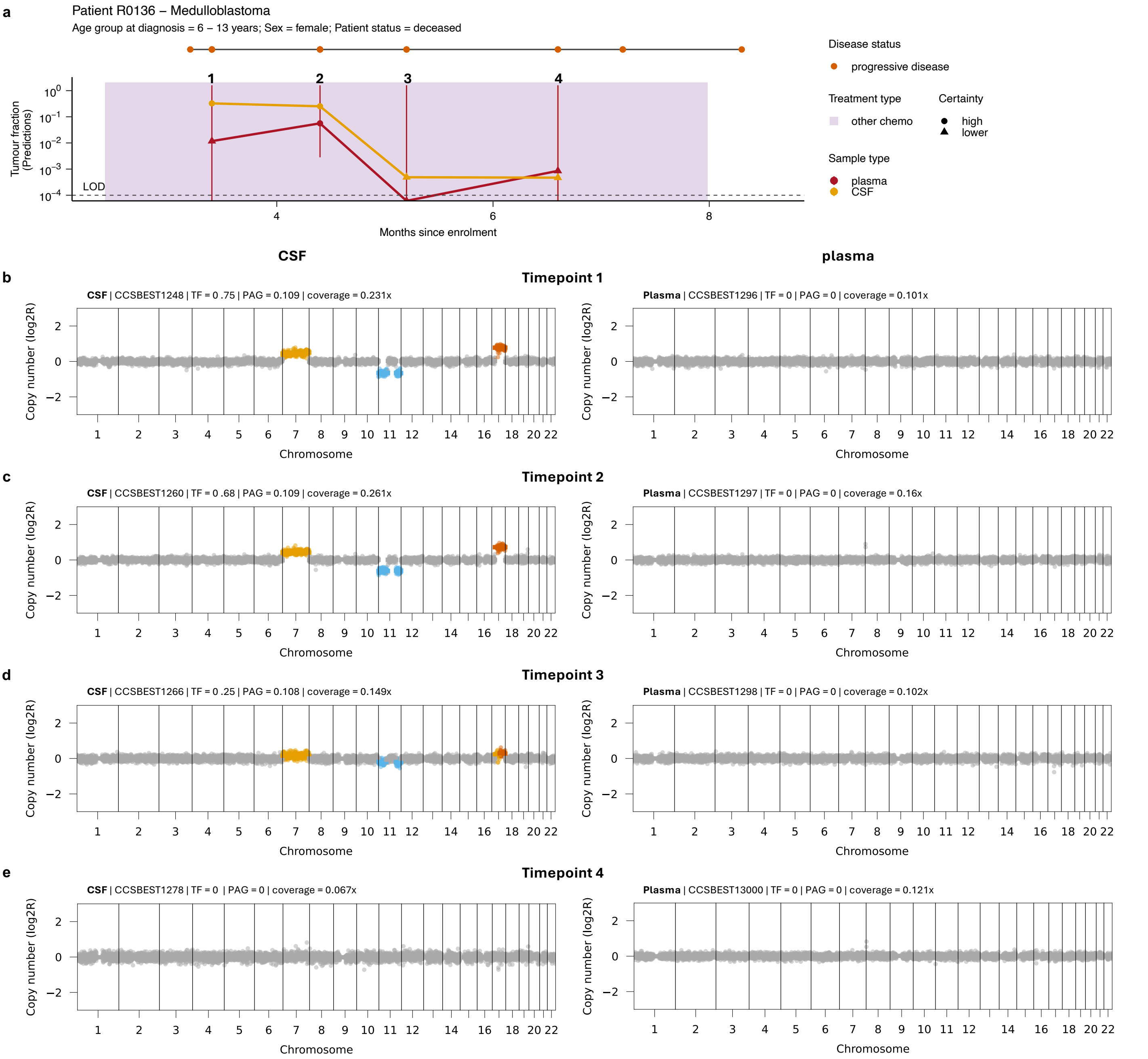


**Extended Data Figure 5. Longitudinal tracking of tumour burden using matched plasma and CSF cfDNA samples from medulloblastoma Patient R0136.**
**(a)** Longitudinal disease trajectory for a female medulloblastoma patient showing the TF predictions computed using multimodal CP obtained from time-matched plasma (red) or CSF (orange) samples. Predictions for which the 90% confidence interval goes beyond the LOD (as indicated by the dashed line) are considered of low certainty and are depicted as triangles. Treatment is indicated by coloured boxes. Disease status by RECIST 1.1 criteria are indicated for each cfDNA sample by coloured points. **(b-e)** Genome-wide COPYBARA copy number profile for matched CSF (left) and plasma (right) cfDNA samples for selected timepoints. Gains and amplifications are shown in yellow and orange, respectively. Losses and deletions are shown in light blue and dark blue, respectively.
CN deviation: copy number deviation; PAG: percentage abnormal genome; TF: tumour fraction.

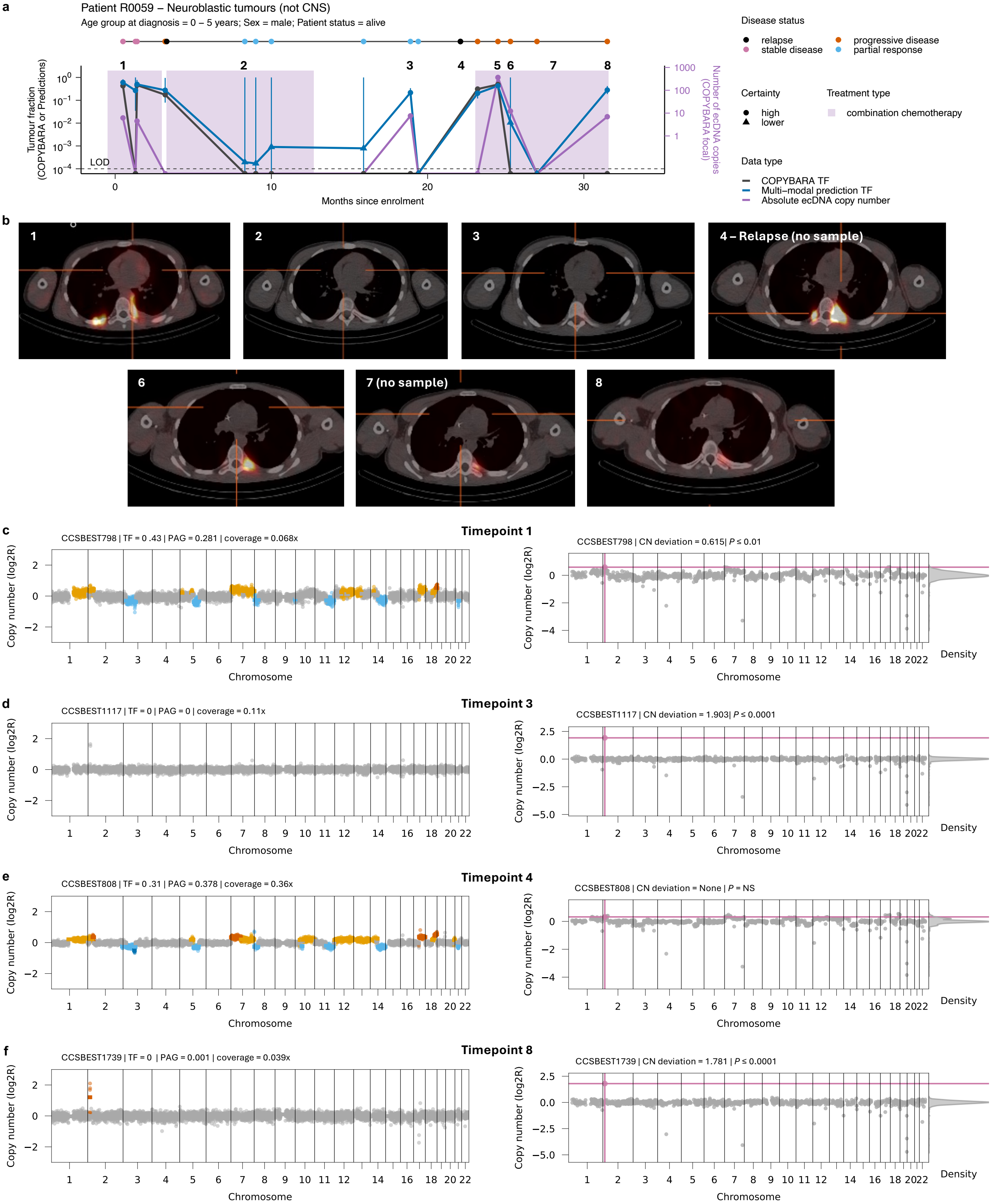


**Extended Data Figure 6. Longitudinal tracking of tumour burden using ecDNA and multimodal CP for neuroblastoma Patient R0059.**

**(a)** Longitudinal disease trajectory for a male neuroblastoma patient showing the TF predictions over time using either COPYBARA (genome-wide copy number analysis; dark grey) or predictions computed using multimodal CP (blue). 90% confidence intervals for TF predictions are shown in blue. Predictions for which the 90% confidence interval goes beyond the LOD (as indicated by the dashed line) are considered of low certainty and are depicted as triangles. The purple line shows the absolute ecDNA copy number detected using COPYBARA-focal informed by ecDNA-associated SV breakpoints. Treatment is indicated by coloured boxes. Disease status by RECIST 1.1 criteria are indicated for each cfDNA sample by coloured points. **(b)** Clinical images from metaiodobenzylguanidine (MIBG) scans taken at the timepoints marked with numbers. **(c-f)** Genome-wide COPYBARA copy number profile (left) and detection of the ecDNA (shown in pink) using COPYBARA-focal (right) for selected timepoints. Gains and amplifications are shown in yellow and orange, respectively. Losses and deletions are shown in light blue and dark blue, respectively.

CN deviation: copy number deviation; ecDNA: extrachromosomal DNA; LOD: limit of detection; PAG: percentage abnormal genome; SCNA: somatic copy number aberration; SV: structural variant; TF: tumour fraction.

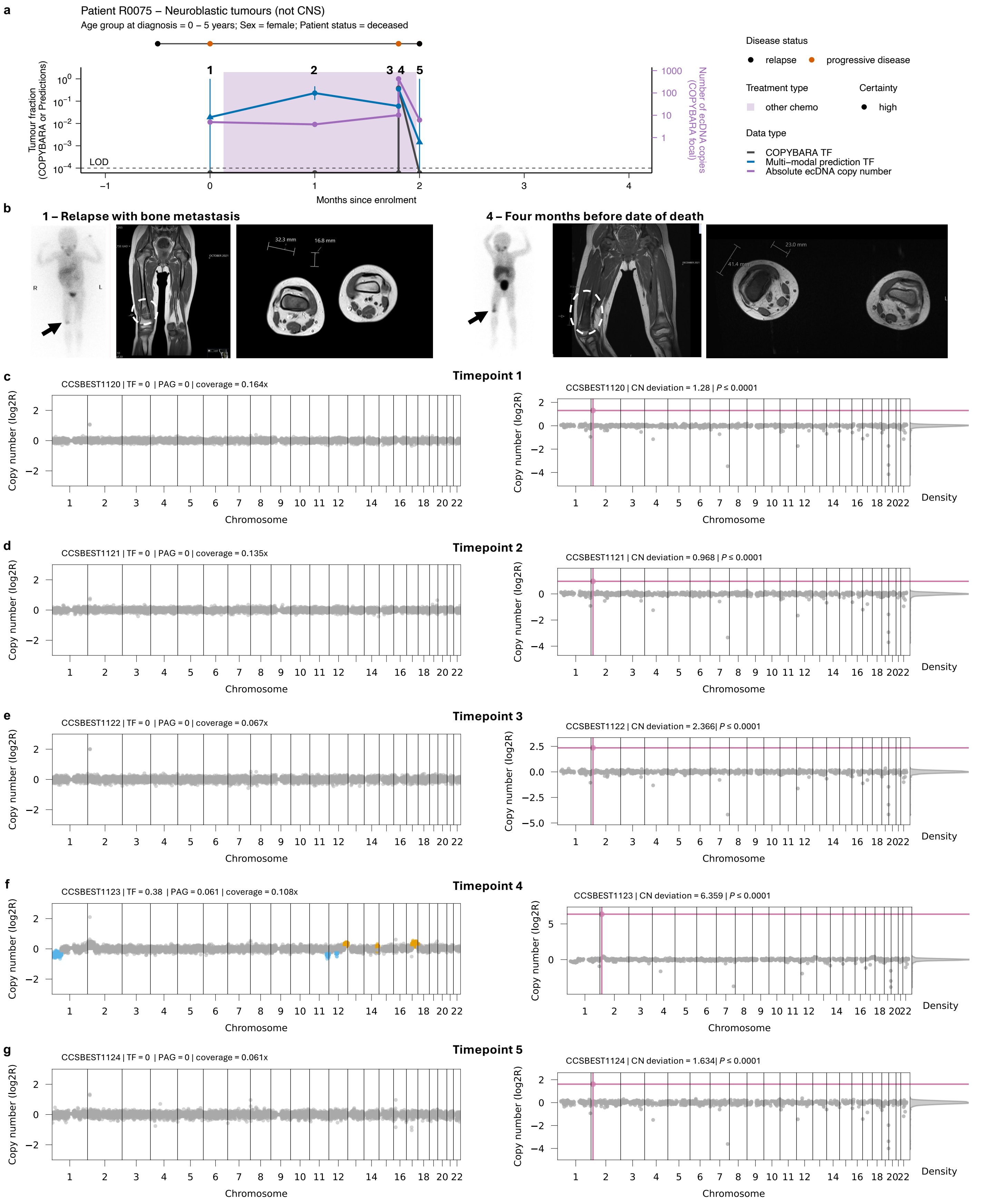


**Extended Data Figure 7. Longitudinal tracking of tumour burden using ecDNA and multimodal CP for neuroblastoma Patient R0075.**
**(a)** Longitudinal disease trajectory for a female neuroblastoma patient with bone metastases showing the TF predictions over time using either COPYBARA (genome-wide copy number analysis; dark grey) or predictions computed using multimodal CP (blue). 90% confidence intervals for TF predictions are shown in blue. Predictions for which the 90% confidence interval goes beyond the LOD (as indicated by the dashed line) are considered of low certainty and are depicted as triangles. The purple line shows the absolute ecDNA copy number detected using COPYBARA-focal informed by ecDNA-associated SV breakpoints. Treatment is indicated by coloured boxes. Disease status by RECIST 1.1 criteria are indicated for each cfDNA sample by coloured points. **(b)** Clinical images from metaiodobenzylguanidine (MIBG) scans taken at the timepoints marked with numbers. **(c-g)** Genome-wide COPYBARA copy number profile (left) and detection of the ecDNA (shown in pink) using COPYBARA-focal (right) for selected timepoints. Gains and amplifications are shown in yellow and orange, respectively. Losses and deletions are shown in light blue and dark blue, respectively. CN deviation: copy number deviation; ecDNA: extrachromosomal DNA; LOD: limit of detection; PAG: percentage abnormal genome; SCNA: somatic copy number aberration; SV: structural variant; TF: tumour fraction.

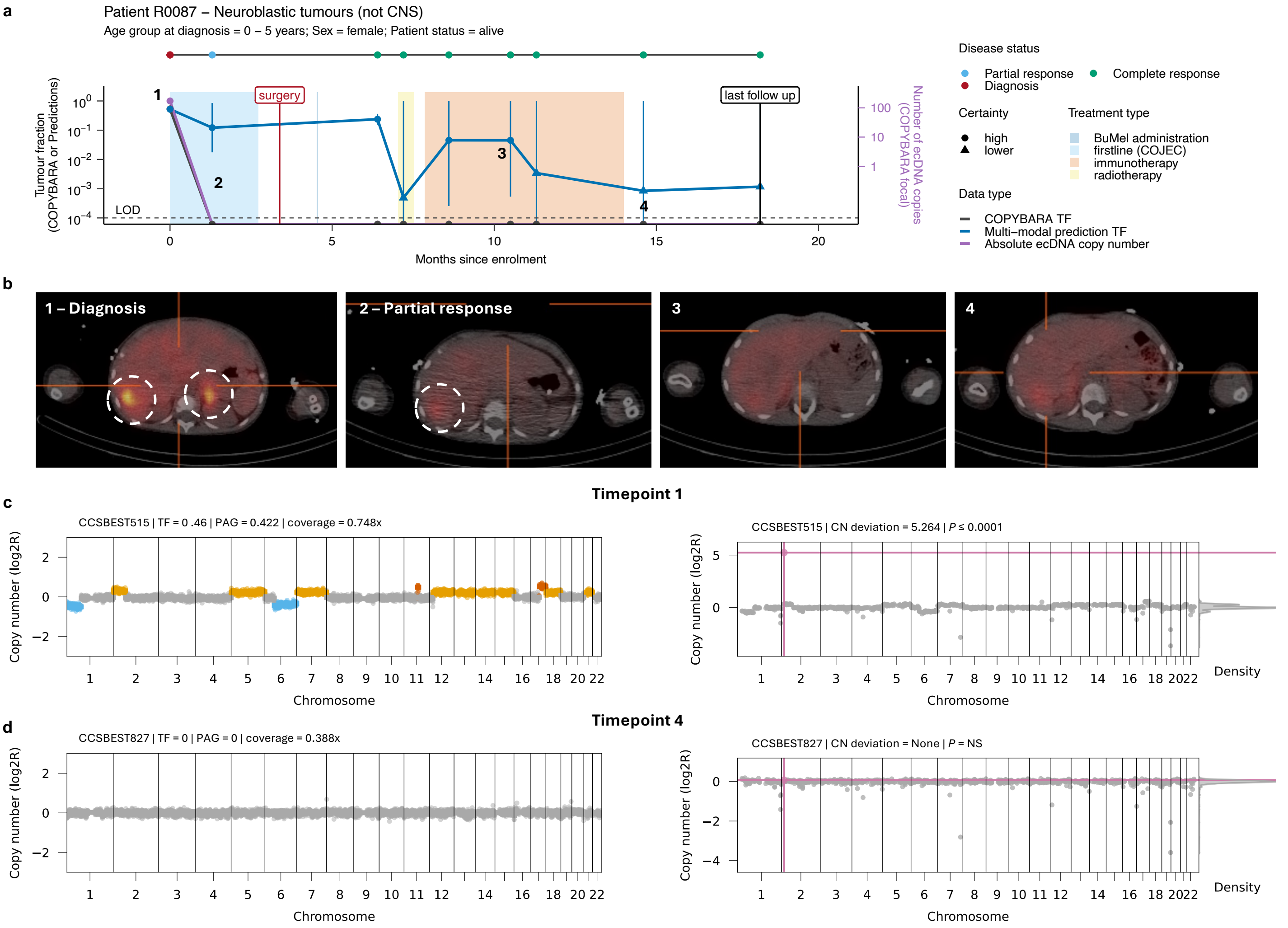


**Extended Data Figure 8. Longitudinal tracking of tumour burden using ecDNA and multimodal CP for neuroblastoma Patient R0087.**

**(a)** Longitudinal disease trajectory for a female neuroblastoma patient showing the TF predictions over time using either COPYBARA (genome-wide copy number analysis; dark grey) or predictions computed using multimodal CP (blue). 90% confidence intervals for TF predictions are shown in blue. Predictions for which the 90% confidence interval goes beyond the LOD (as indicated by the dashed line) are considered of low certainty and are depicted as triangles. The purple line shows the absolute ecDNA copy number detected using COPYBARA-focal informed by ecDNA-associated SV breakpoints. Treatment is indicated by coloured boxes. Disease status by RECIST 1.1 criteria are indicated for each cfDNA sample by coloured points. **(b)** Clinical images from metaiodobenzylguanidine (MIBG) scans taken at the timepoints marked with numbers. **(c-d)** Genome-wide COPYBARA copy number profile (left) and detection of the ecDNA (shown in pink) using COPYBARA-focal (right) for selected timepoints. Gains and amplifications are shown in yellow and orange, respectively. Losses and deletions are shown in light blue and dark blue, respectively.

CN deviation: copy number deviation; ecDNA: extrachromosomal DNA; LOD: limit of detection; PAG: percentage abnormal genome; SCNA: somatic copy number aberration; SV: structural variant; TF: tumour fraction.

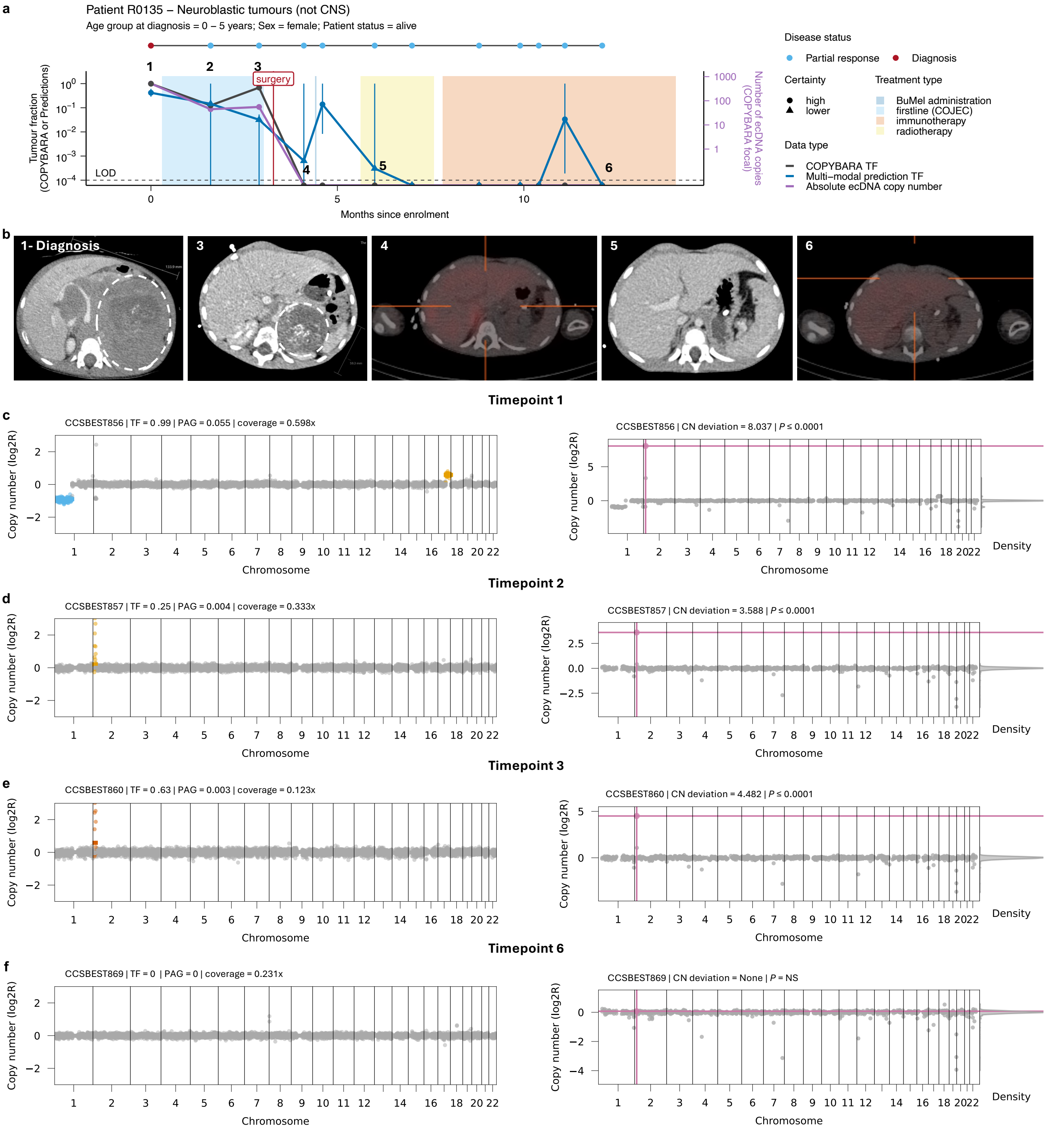


**Extended Data Figure 9. Longitudinal tracking of tumour burden using ecDNA and multimodal CP for neuroblastoma Patient R0135.**
**(a)** Longitudinal disease trajectory for a female neuroblastoma patient showing the TF predictions over time using either COPYBARA (genome-wide copy number analysis; dark grey) or predictions computed using multimodal CP (blue). 90% confidence intervals for TF predictions are shown in blue. Predictions for which the 90% confidence interval goes beyond the LOD (as indicated by the dashed line) are considered of low certainty and are depicted as triangles. The purple line shows the absolute ecDNA copy number detected using COPYBARA-focal informed by ecDNA-associated SV breakpoints. Treatment is indicated by coloured boxes. Disease status by RECIST 1.1 criteria are indicated for each cfDNA sample by coloured points. **(b)** Clinical images from metaiodobenzylguanidine (MIBG) scans taken at the timepoints marked with numbers. **(c-f)** Genome-wide COPYBARA copy number profile (left) and detection of the ecDNA (shown in pink) using COPYBARA-focal (right) for selected timepoints. Gains and amplifications are shown in yellow and orange, respectively. Losses and deletions are shown in light blue and dark blue, respectively.
CN deviation: copy number deviation; ecDNA: extrachromosomal DNA; LOD: limit of detection; PAG: percentage abnormal genome; SCNA: somatic copy number aberration; SV: structural variant; TF: tumour fraction.

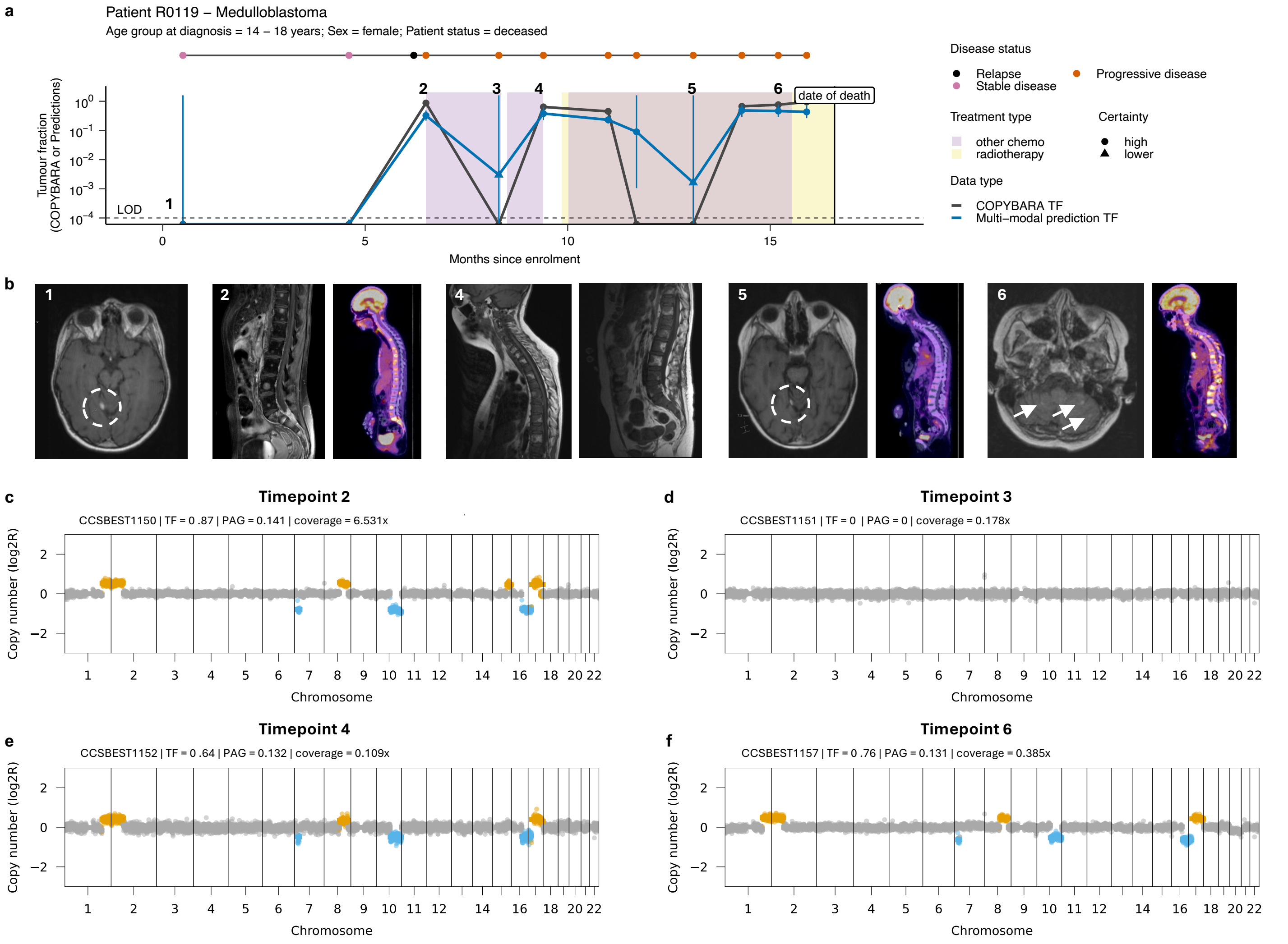


**Extended Data Figure 10. Longitudinal tracking of tumour burden using ecDNA and multimodal CP for medulloblastoma Patient R0119.**
**(a)** Longitudinal disease trajectory for a male medulloblastoma patient with metastatic disease showing the TF predictions over time using either COPYBARA (genome-wide copy number analysis; dark grey) or predictions computed using multimodal CP (blue). 90% confidence intervals for TF predictions are shown in blue. Predictions for which the 90% confidence interval goes beyond the LOD (as indicated by the dashed line) are considered of low certainty and are depicted as triangles. The purple line shows the absolute ecDNA copy number detected using COPYBARA-focal informed by ecDNA-associated SV breakpoints. Treatment is indicated by coloured boxes. Disease status by RECIST 1.1 criteria are indicated for each cfDNA sample by coloured points. **(b)** Clinical images from MRI and FDG-PET scans taken at the timepoints marked with numbers. **(c-f)** Genome-wide COPYBARA copy number profile (left) and detection of the ecDNA (shown in pink) using COPYBARA-focal (right) for selected timepoints. Gains and amplifications are shown in yellow and orange, respectively. Losses and deletions are shown in light blue and dark blue, respectively.
CN deviation: copy number deviation; ecDNA: extrachromosomal DNA; LOD: limit of detection; PAG: percentage abnormal genome; SCNA: somatic copy number aberration; SV: structural variant; TF: tumour fraction.

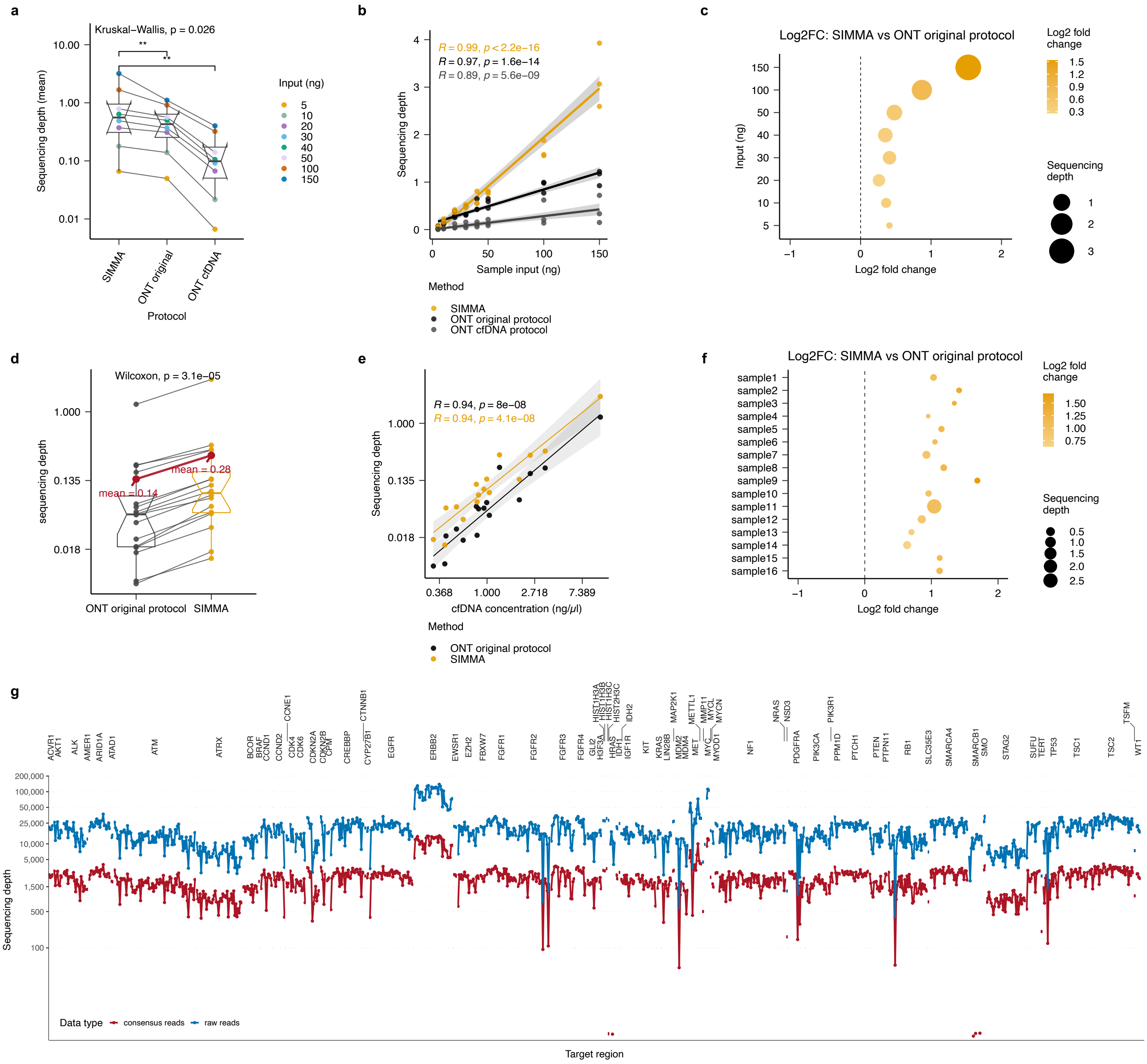


**Supplementary Figure 1**. **Assessment of the sequencing efficiency of SIMMA. (a)** Boxplot comparing sequencing outputs (depth) obtained from a 30-fold dilutions series of NA12878 DNA ranging from 5-150ng (see Fig. 1b) using the SIMMA, original ONT and cfDNA ONT protocols. **(b)** Correlation of total cfDNA input (ng) vs sequencing output (depth) comparing the SIMMA, original ONT and cfDNA ONT protocols. **(c)** Log2 fold enrichment of sequencing output (depth) of SIMMA sequencing compared to the original ONT protocol. Circle sizes indicated the sequencing depth achieved. Side-by-side comparison showing **(d)** boxplots, **(e)** correlation plot, and **(f)** log2 fold enrichment plot of 16 SMPaeds samples sequenced using either the SIMMA (orange) or original ONT protocols (black). **(g)** Sequencing depth for all targets regions included in the custom IDT panel across 76 genes, including amplifications in *ERBB2, MET* and *MYC*. Raw and consensus reads (post UMI-processing) are shown in blue and red, respectively. Box plots in **a,d** show the median, first and third quartiles (boxes) and the whiskers encompass observations within 1.5× the interquartile range from the first and third quartiles.

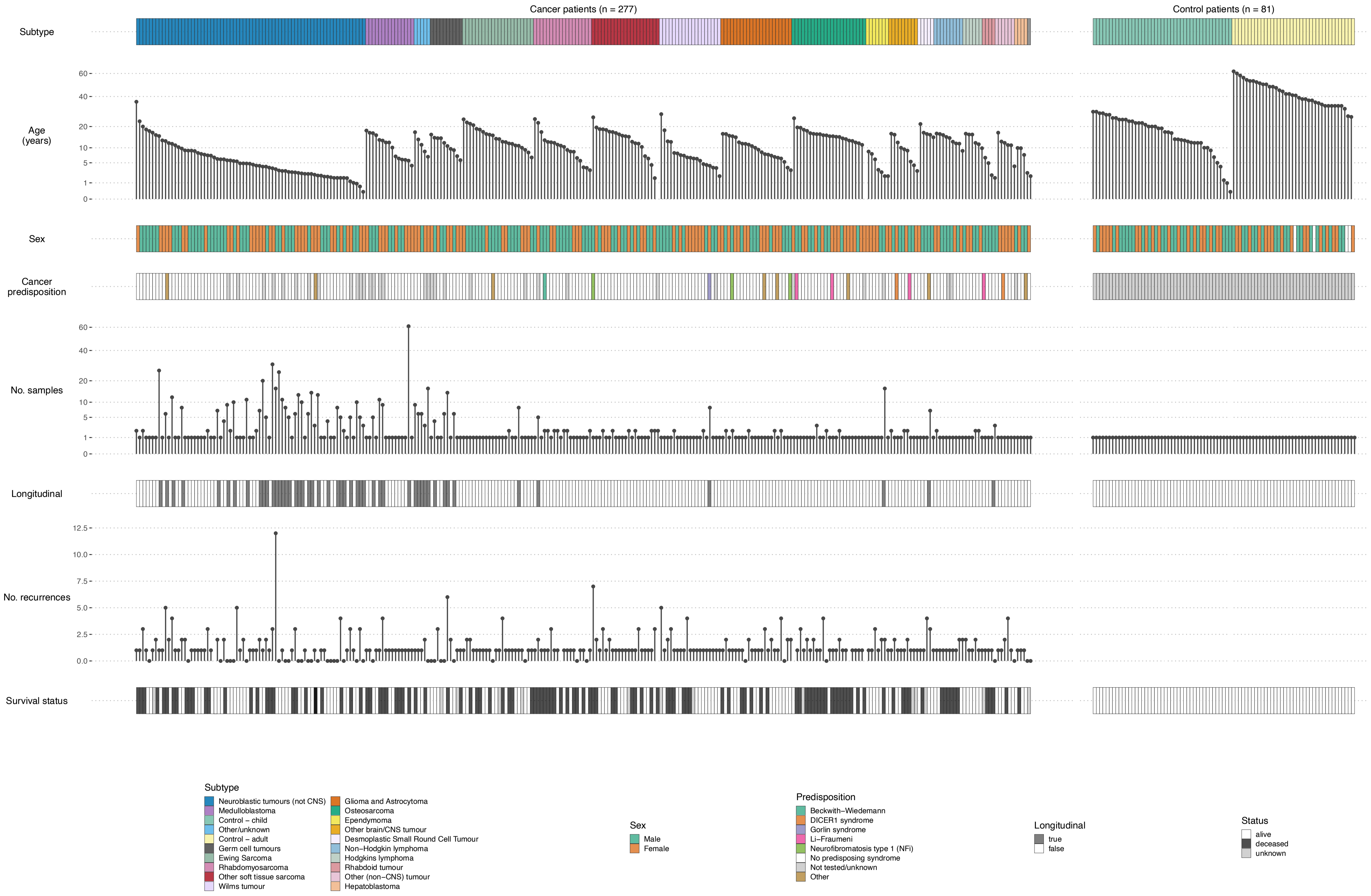


**Supplementary Figure 2. Cohort overview** of patients included in this study showing disease subtypes, age, sex, cancer predisposition syndromes, total number of samples collected for each patient, whether longitudinal samples were analysed, total number of disease recurrences/relapses and survival status.

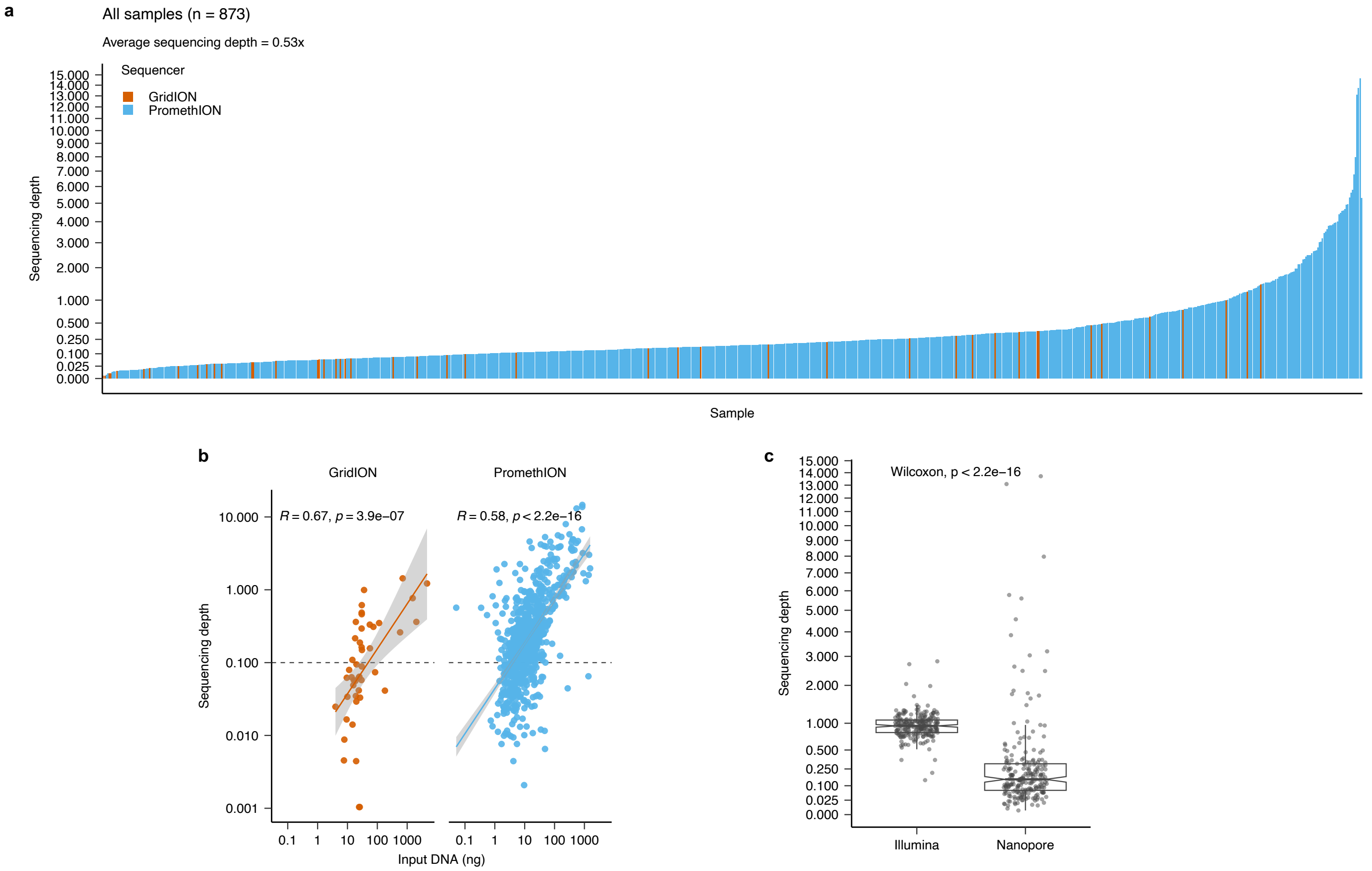


**Supplementary Figure 3. SIMMA sequencing performance across all cfDNA samples analysed in this study. (a)** Genomewide sequencing depth (i.e., genome equivalents) achieved using SIMMA for all cfDNA samples included in this study. Samples sequenced using GridION and PromethION sequencing machines (ONT) are shown in orange and blue, respectively. Samples are shaded by whether the target depth of 0.1x (dashed horizontal line) was achieved. Mean sequencing depth across the entire sample cohort was 0.5x. **(b)** Correlation of total cfDNA input (ng) vs genomewide sequencing depth for all samples. **(c)** Comparison of sequencing depth achieved using Illumina sequencing with PCR amplification and nanopore sequencing of non-amplified native cfDNA using SIMMA for a subset of 225 samples with aliquot-matched sequencing data. Box plots in **c** show the median, first and third quartiles (boxes) and the whiskers encompass observations within 1.5× the interquartile range from the first and third quartiles.

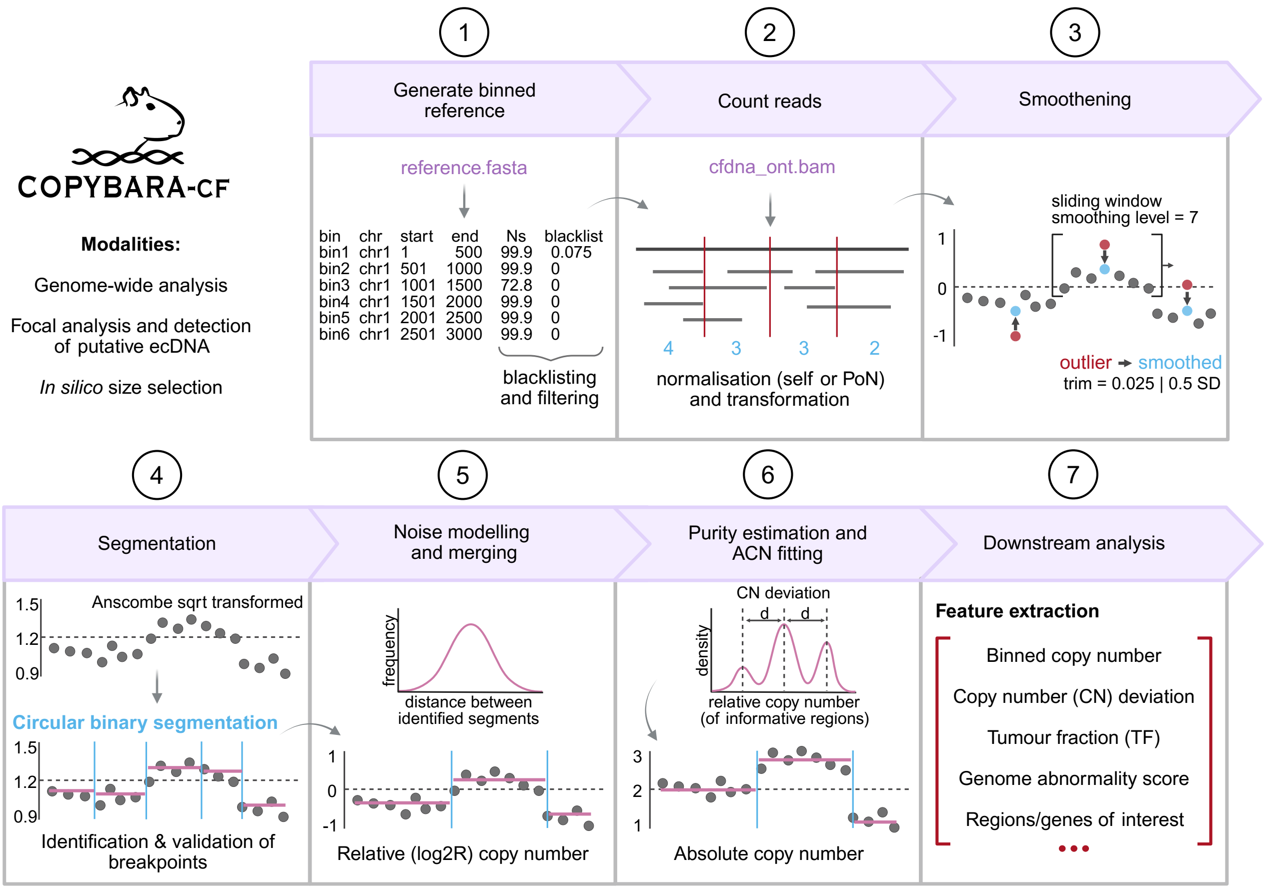


**Supplementary Figure 4. Overview of COPYBARA.** Overview of the methodology for the analysis of copy number aberrations from low coverage single-molecule cfDNA whole-genome sequencing data using COPYBARA.
CN deviation: copy number deviation; PoN: panel of normals; TF: tumour fraction

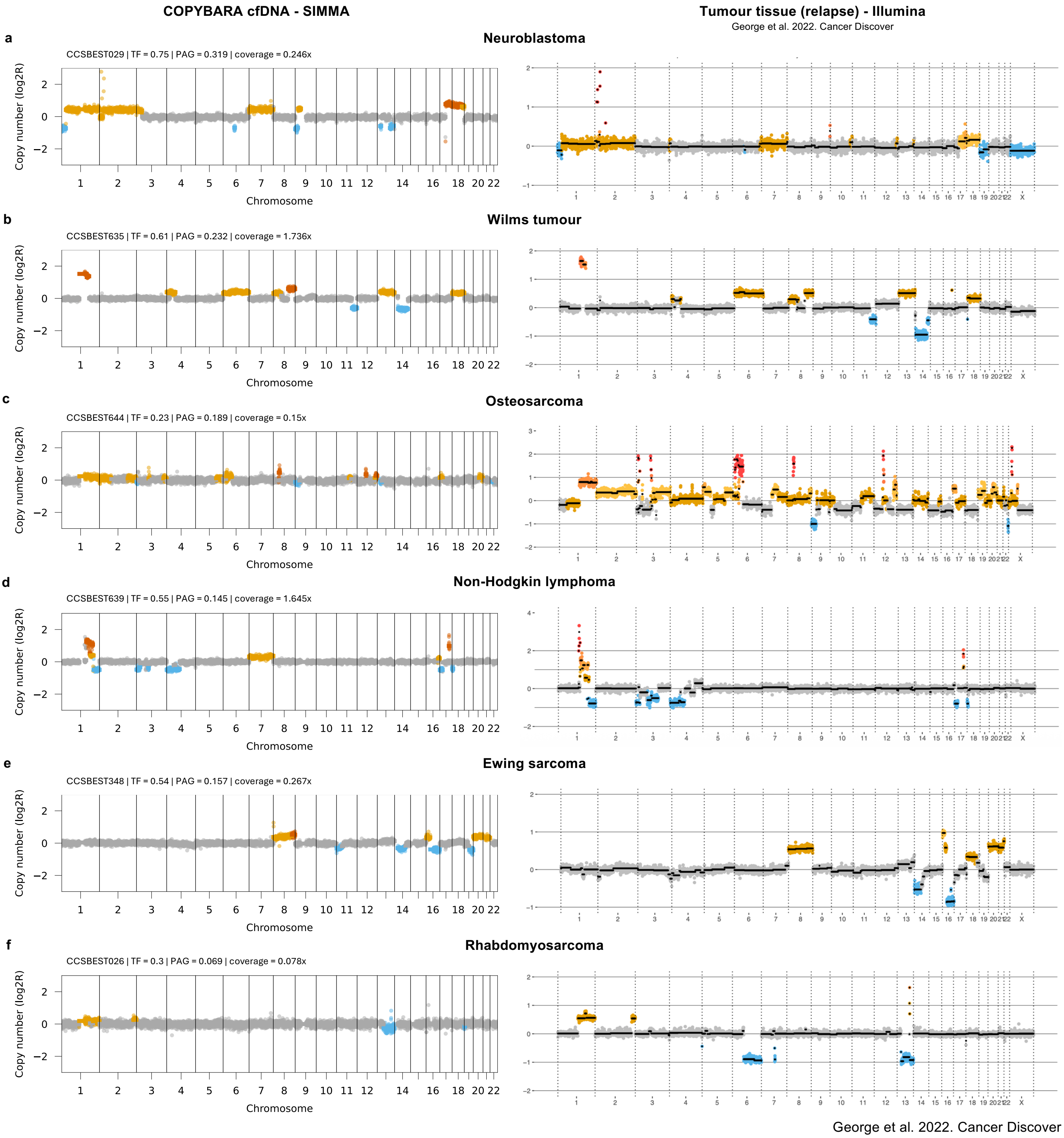


**Supplementary Figure 5. Comparison of SCNAs detected using SIMMA and matched Illumina tumour WGS data across different paediatric cancer types.** Comparison of genomewide SCNA profiles inferred using COPYBARA applied to cfDNA data generated using SIMMA against SCNA profiles inferred from standard-of-care Illumina WGS data for relapse tumour tissue from the same patient (George et al. 2022 Cancer Discovery).

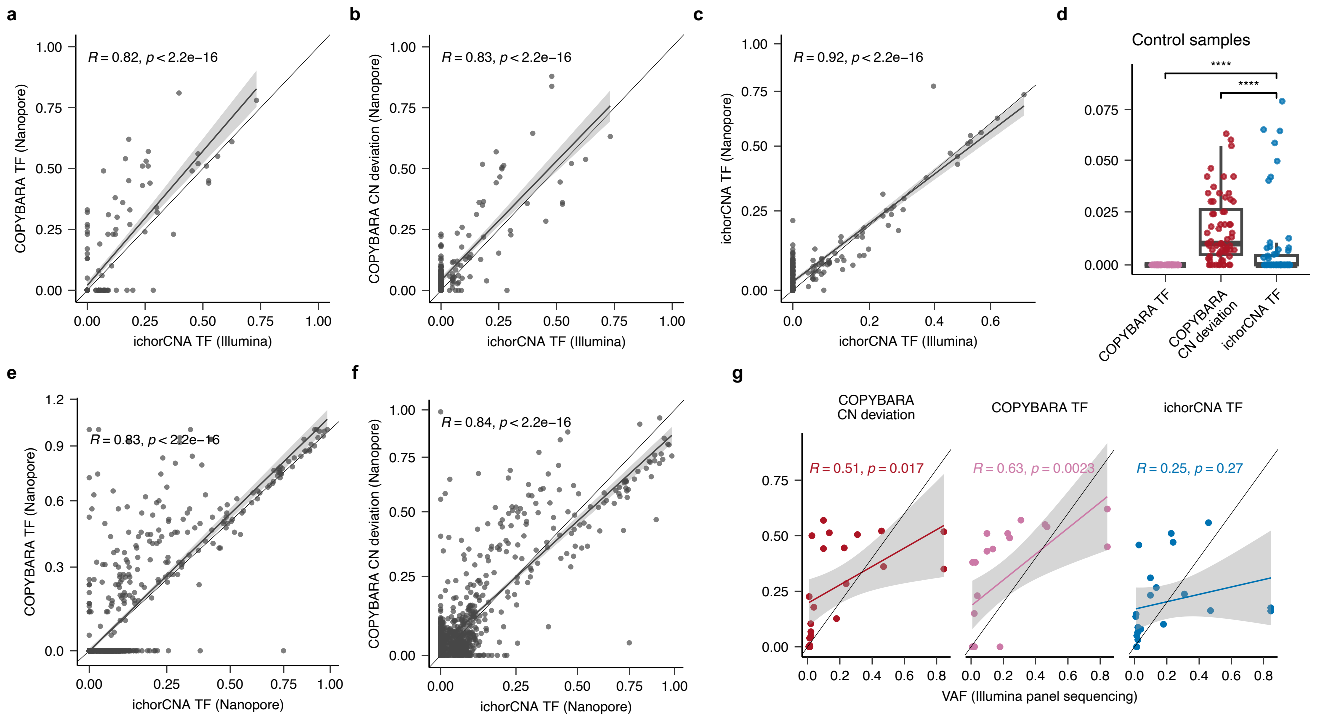


**Supplementary Figure 6. Validation of COPYBARA for SCNA analysis using single-molecule cfDNA data. (a)** Correlation of tumour fraction (TF) values estimated with COPYBARA (nanopore) and ichorCNA (Illumina) for matched samples (n=198). **(b)** Correlation of the copy number (CN) deviation values estimated with COPYBARA (nanopore) vs ichorCNA TF (Illumina) for matched samples. **(c)** Correlation of ichorCNA TF values estimated from nanopore vs Illumina cfDNA sequencing data for matched samples. **(d)** Comparison of COPYBARA TF, COPYBARA CN deviation and ichorCNA TF values across all cancer-free controls analysed in this study using SIMMA (n=46). **(e)** Correlation of COPYBARA TF and ichorCNA TF values estimated from nanopore data across all samples analysed in this study using SIMMA (n=838). **(f)** Correlation of CN deviation values estimated with COPYBARA and ichorCNA TF values estimated from Nanopore data across all samples included in this study. **(g)** Correlation of COPYBARA CN deviation values (left), COPYBARA TF values (middle) and ichorCNA TF values (right) vs variant allele frequencies (VAF) of somatic SNVs detected using targeted panel Illumina sequencing from matched cfDNA samples (George et al. *Cancer Discovery,* 2022). Box plots in **d** show the median, first and third quartiles (boxes) and the whiskers encompass observations within 1.5× the interquartile range from the first and third quartiles.

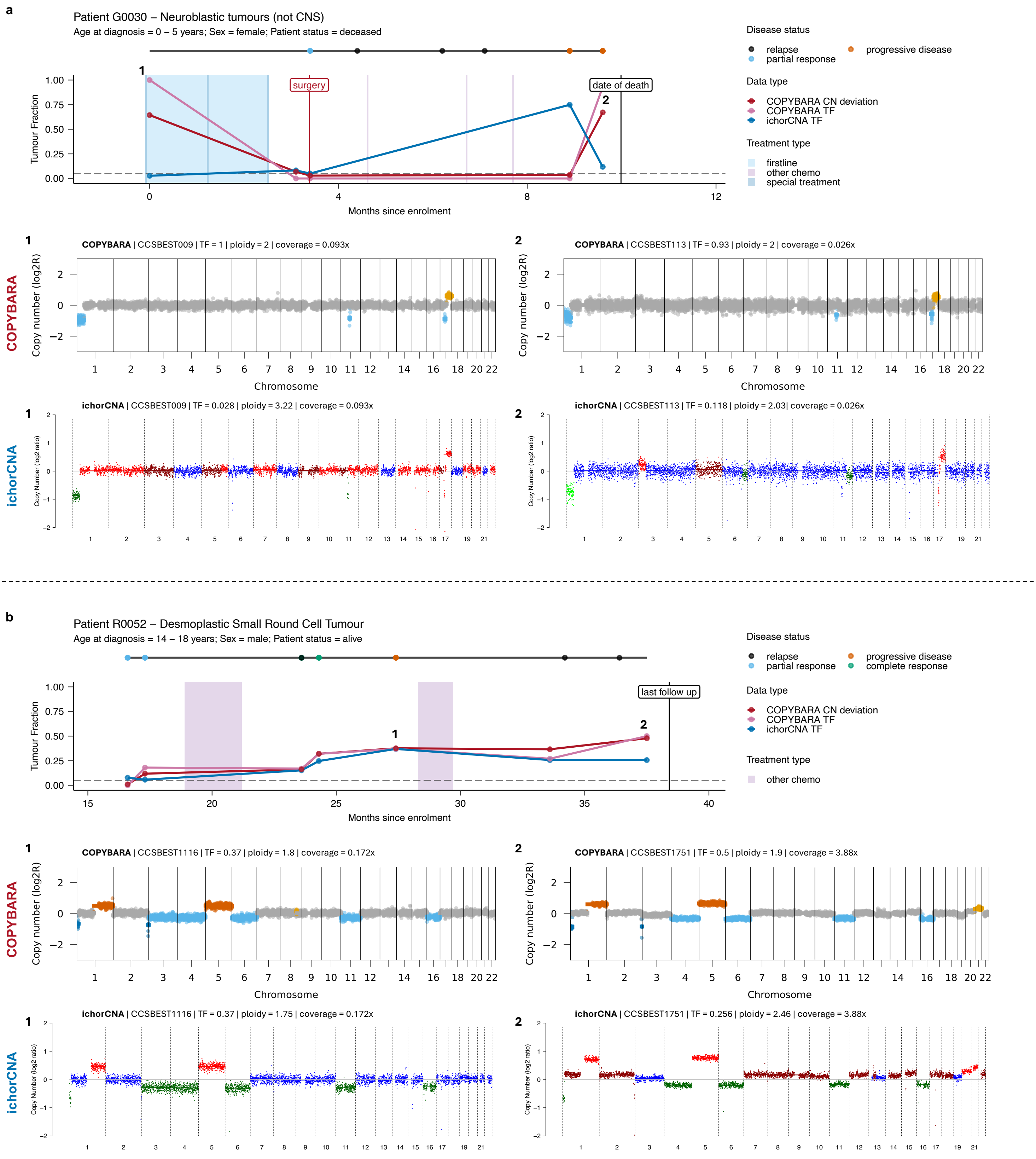


**Supplementary Figure 7. Validation of COPYBARA for SCNA analysis of single-molecule cfDNA data.** Comparison of tumour fraction (TF) values estimated using ichorCNA (blue) or COPYBARA (TF in pink and CN deviation shown in red) for patients **(a)** G0030 and **(b)** R0052. For each patient, the top panel depicts the longitudinal disease trajectory showing the TF and CN deviation values computed using COPYBARA and the TF values estimated using ichorCNA. Genomewide COPYBARA and ichorCNA CN profiles are shown for the selected timepoints marked with numbers for which COPYBARA and ichorCNA predicted discrepant TF estimates. Overall, these results show the the higher accuracy of COPYBARA for the analysis of SCNAs using single-molecule cfDNA data.

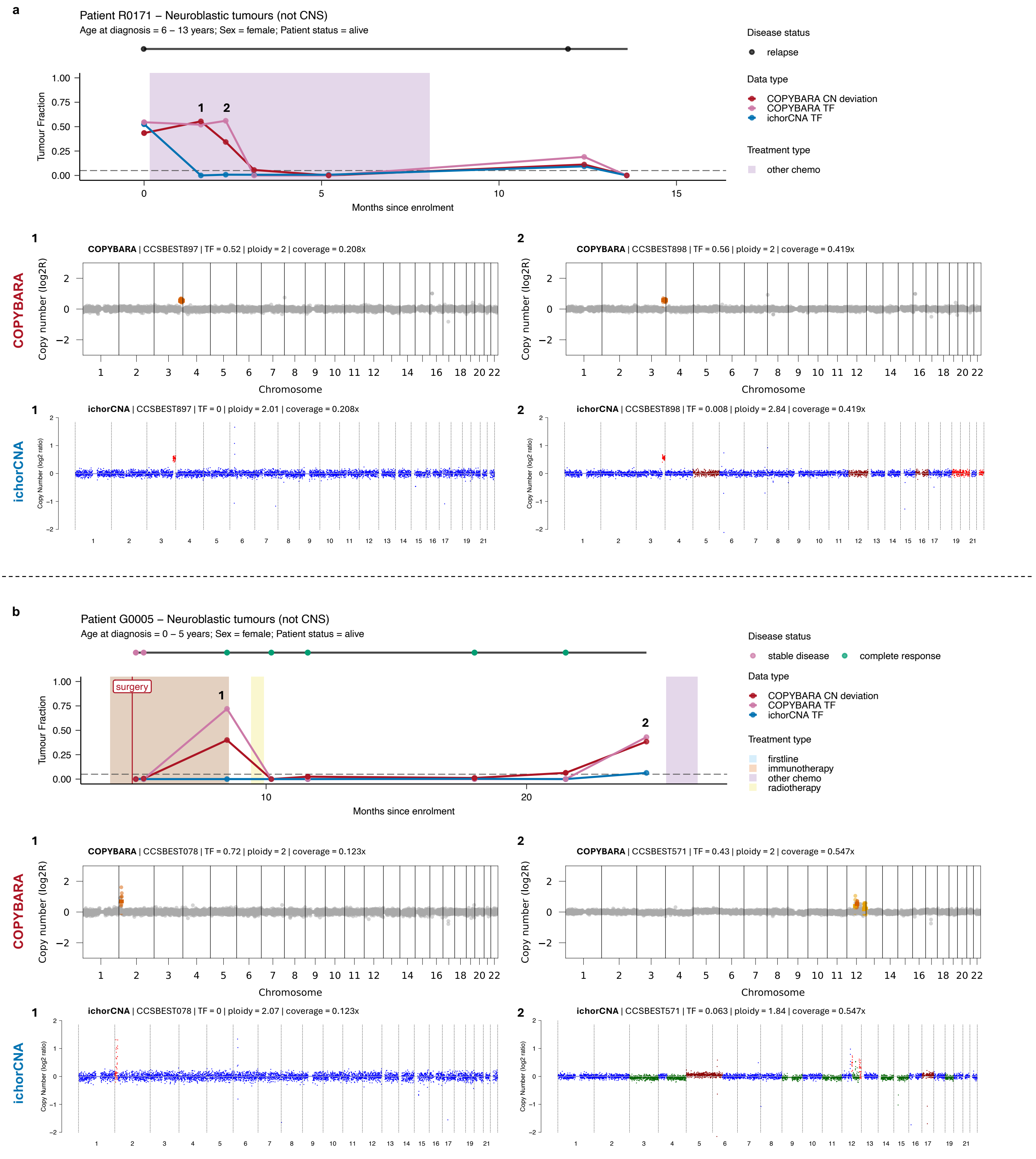


**Supplementary Figure 8. Validation of COPYBARA for copy number analysis of single-molecule cfDNA data. .** Comparison of tumour fraction (TF) values estimated using ichorCNA (blue) or COPYBARA (TF in pink and CN deviation shown in red) for patients **(a)** R0171 and **(b)** G0005. For each patient, the top panel depicts the longitudinal disease trajectory showing the TF and CN deviation values computed using COPYBARA and the TF values estimated using ichorCNA. Genomewide COPYBARA and ichorCNA CN profiles are shown for the selected timepoints marked with numbers for which COPYBARA and ichorCNA predicted discrepant TF estimates. Overall, these results show the the higher accuracy of COPYBARA for the analysis of SCNAs using single-molecule cfDNA data.

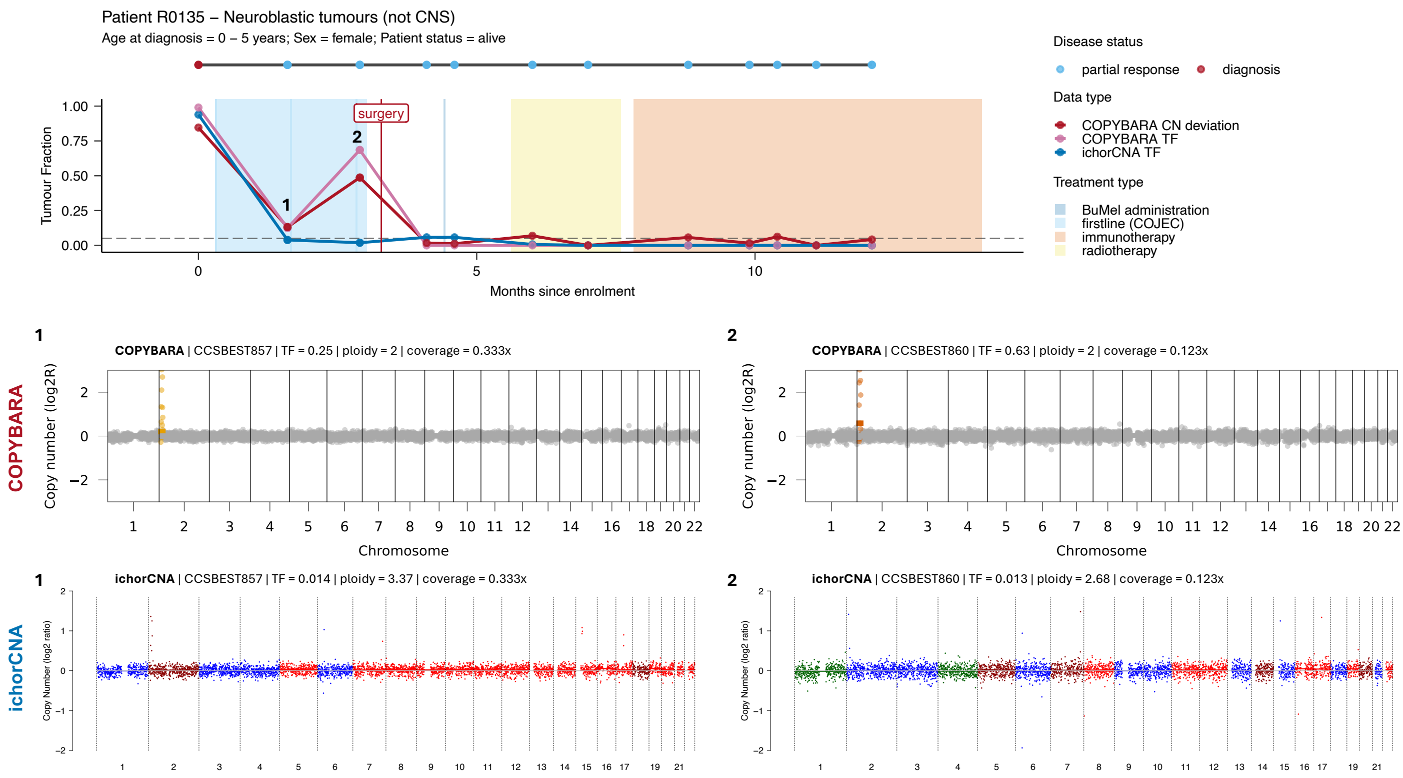


**Supplementary Figure 9. Validation of COPYBARA for copy number analysis of single-molecule cfDNA data. .** Comparison of tumour fraction (TF) values estimated using ichorCNA (blue) or COPYBARA (TF in pink and CN deviation shown in red) for patients R0135. For each patient, the top panel depicts the longitudinal disease trajectory showing the TF and CN deviation values computed using COPYBARA and the TF values estimated using ichorCNA. Genomewide COPYBARA and ichorCNA CN profiles are shown for the selected timepoints marked with numbers for which COPYBARA and ichorCNA predicted discrepant TF estimates. Overall, these results show the the higher accuracy of COPYBARA for the analysis of SCNAs using single-molecule cfDNA data when ctDNA signal is concentrated on focal amplifications.

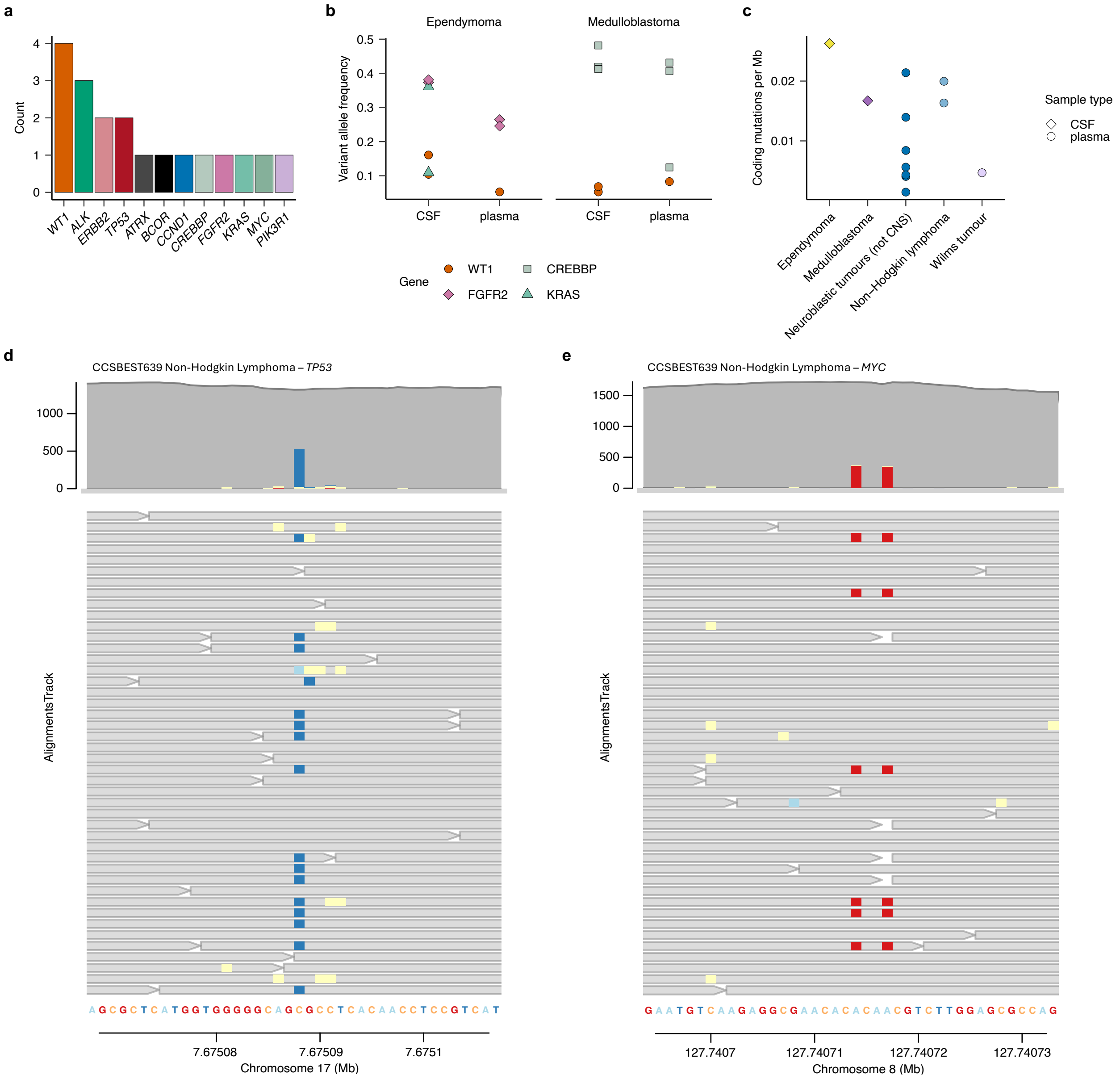


**Supplementary Figure 10. Detection of pathogenic SNVs in cfDNA using SIMMA targeted sequencing.**
**(a)** Barplot showing the total count of pathogenic SNVs detected across oncogenes and tumour suppressor genes. **(b)** Variant allele frequency of detected pathogenic SNVs for matched CSF and plasma samples from one ependymoma and one medulloblastoma patient. **(c)** Mutational burden estimated as the frequency of pathogenic SNVs per megabase (Mb) across different tumour subtypes. (**d-e**) Aligned cfDNA consensus sequencing reads at two genomic regions in which pathogenic SNVs were detected. Non-reference bases correspond to pathogenic SNVs in **(d)** *TP53* and **(e)** *MYC* from cfDNA sample CCSBEST639. Light yellow squares represent bases that were identified as ambiguous across reads from the same UMI family during consensus read calling and were masked (N) to remove potential sequencing errors.

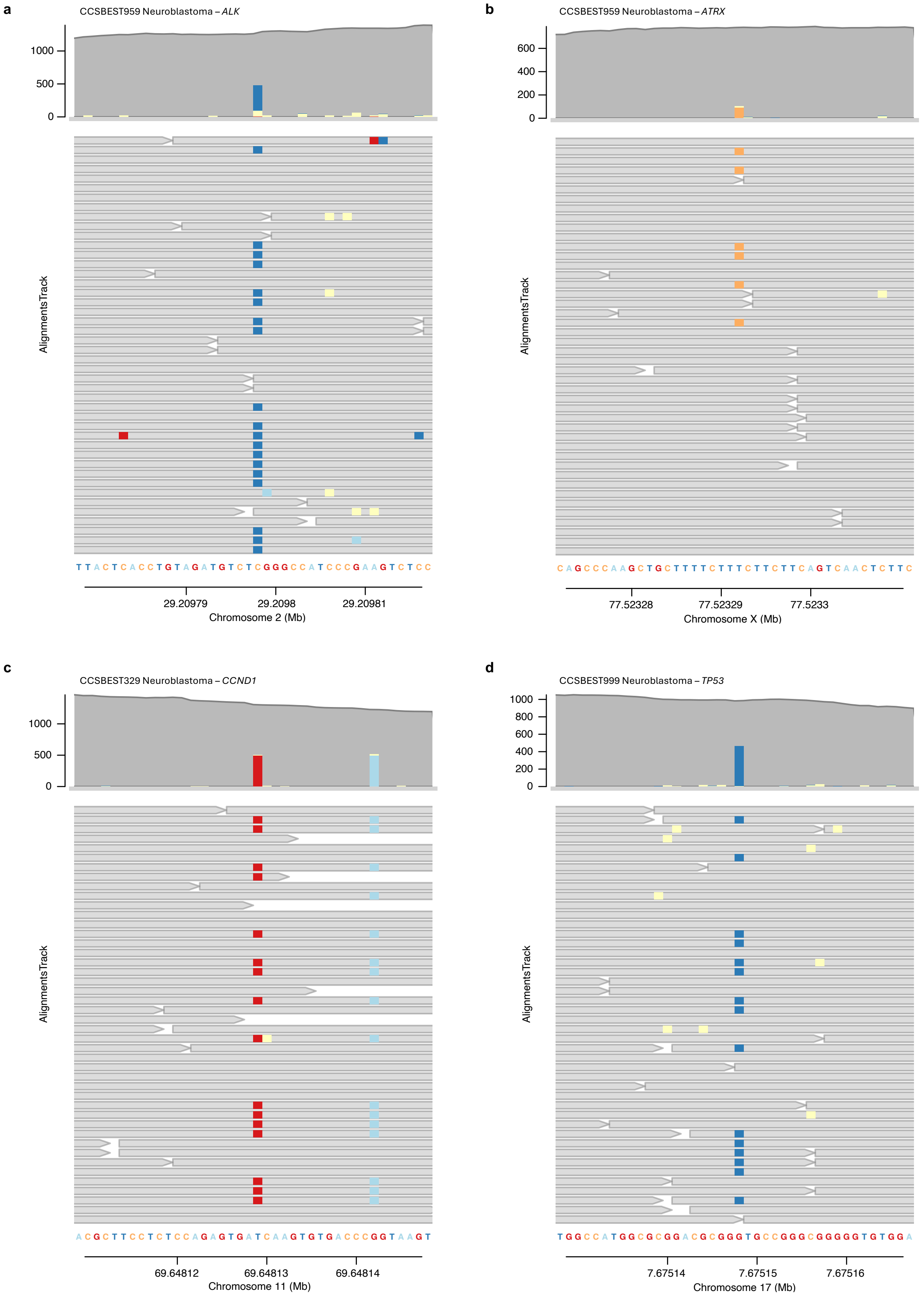


**Supplementary Figure 11. Detection of pathogenic SNVs in cfDNA using SIMMA targeted sequencing.**
Aligned cfDNA consensus sequencing reads at genomic regions in which pathogenic SNVs were detected. Non-reference bases correspond to pathogenic SNVs in in **(a)** *ALK* and **(b)** *ATRX* from sample CCSBEST959, **(c)** *CCND1* in sample CCSBEST329 and **(d)** *TP53* in cfDNA sample CCSBEST999. Light yellow squares represent bases that were identified as ambiguous across reads from the same UMI family during consensus read calling and were masked (N) to remove potential sequencing errors. The mutation in *CCND1* in (**c**) is phased with a nearby heterozygous SNP, showing that the mutation is only supported by reads of one parental allele, as expected for somatic mutations.

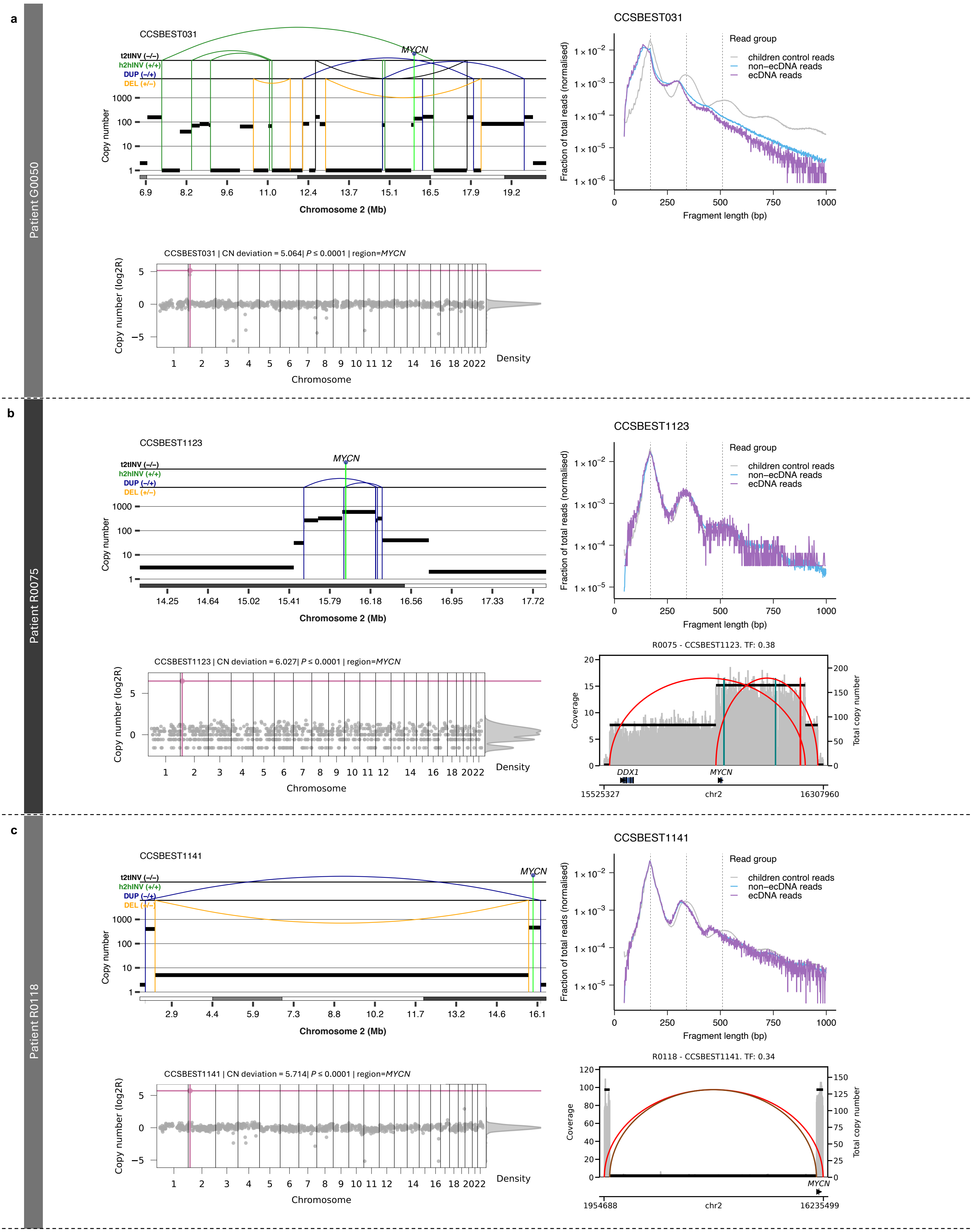

**Supplementary Figure 12. Detection and reconstruction of ecDNA in plasma cfDNA** from patients **(a)** G0050, **(b)** R0075 and **(c)** R0118.
For each patient, the **top left** panel depicts the genomic rearrangement profiles showing SCNAs and SVs detected using SAVANA showing the ecDNA amplifying *MYCN*. Absolute copy number data are represented by the black horizontal lines. SVs are represented by vertical lines. DEL: deletion-like rearrangement; DUP: duplication-like rearrangement; h2hINV: head-to-head inversion; t2tINV, tail-to-tail inversion. The **top right** panel shows the fragment size profile of cfDNA reads mapping to ecDNA (purple) and non-ecDNA (blue) regions. The fragmentation profile computed using cfDNA WGS data paediatric control samples is shown in grey for comparison. The **bottom left** panel shows the detection of ecDNA (pink dot) using COPYBARA-focal. Where available, the **bottom right** panel depicts the genomic copy number profile showing SCNAs and SVs detected using CoRAL.

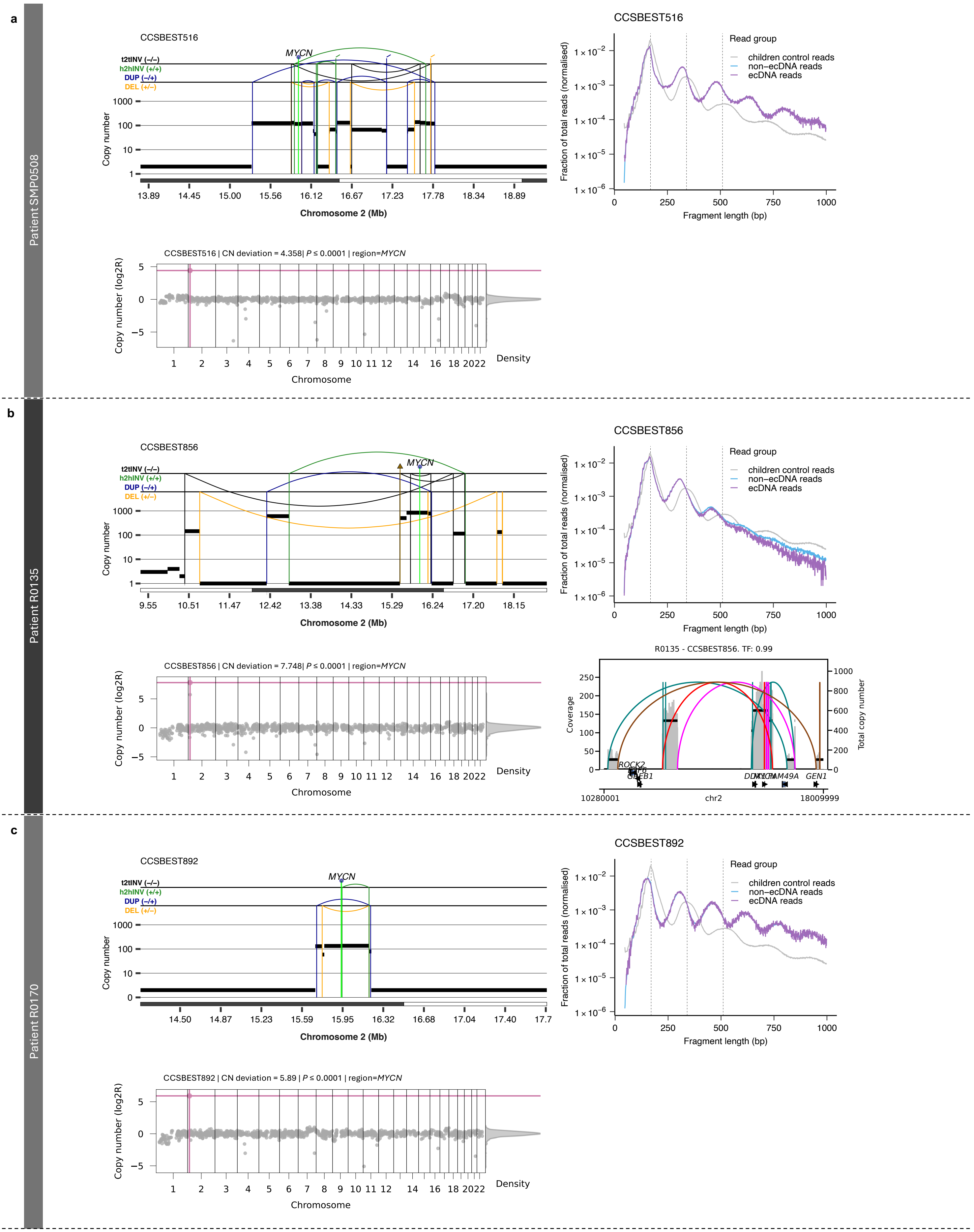


**Supplementary Figure 13. Detection and reconstruction of ecDNA in plasma cfDNA** from patients **(a)** SMP0508, **(b)** R0135 and **(c)** R0170. For each patient, the **top left** panel depicts the genomic rearrangement profiles showing SCNAs and SVs detected using SAVANA showing the ecDNA amplifying *MYCN*. Absolute copy number data are represented by the black horizontal lines. SVs are represented by vertical lines. DEL: deletion-like rearrangement; DUP: duplication-like rearrangement; h2hINV: head-to-head inversion; t2tINV, tail-to-tail inversion. The **top right** panel shows the fragment size profile of cfDNA reads mapping to ecDNA (purple) and non-ecDNA (blue) regions. The fragmentation profile computed using cfDNA WGS data paediatric control samples is shown in grey for comparison. The **bottom left** panel shows the detection of ecDNA (pink dot) using COPYBARA-focal. Where available, the **bottom right** panel depicts the genomic copy number profile showing SCNAs and SVs detected using CoRAL.

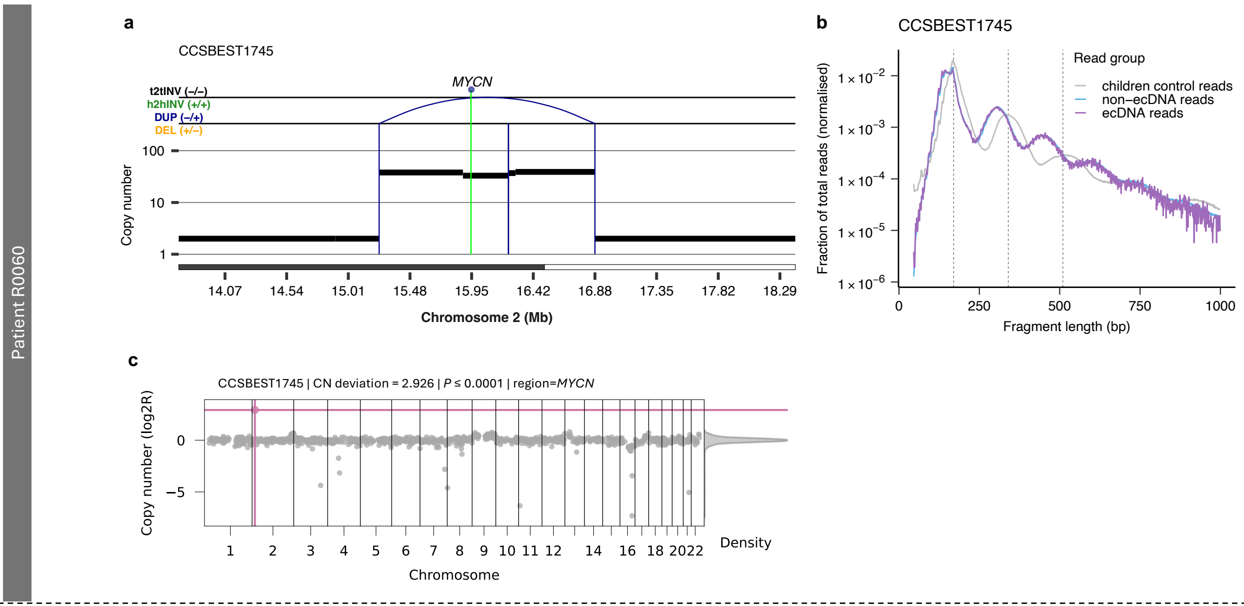


**Supplementary Figure 14. Detection and reconstruction of ecDNA in plasma cfDNA from patients R0060.**

**(a)** Genomic rearrangement profile showing SCNAs and SVs detected using SAVANA showing the ecDNA amplifying *MYCN*. Absolute copy number data are represented by the black horizontal lines. SVs are represented by vertical lines. DEL: deletion-like rearrangement; DUP: duplication-like rearrangement; h2hINV: head-to-head inversion; t2tINV, tail-to-tail inversion. **(b)** Fragment size profile of cfDNA reads mapping to ecDNA (purple) and non-ecDNA (blue) regions. The fragmentation profile computed using cfDNA WGS data paediatric control samples is shown in grey for comparison. **(c)** Detection of ecDNA (pink dot) using COPYBARA-focal.

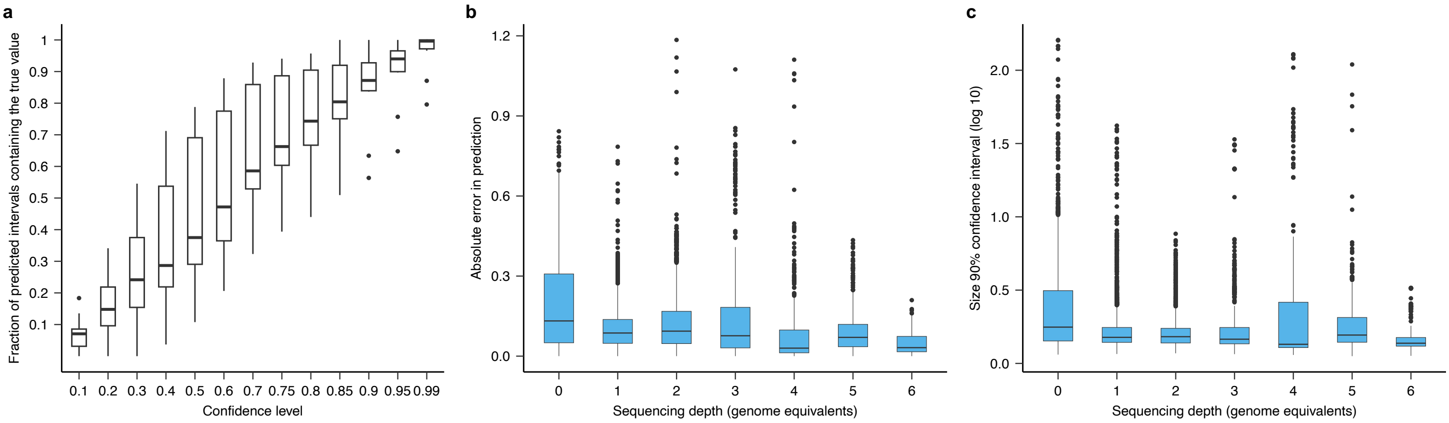


**Supplementary Figure 15. Multimodal Conformal Prediction (CP).**

**(a)** Analysis of the validity of the confidence intervals generated using multimodal CP. Correlation between the theoretical confidence level (1–error rate) and the fraction of predictions for which the true value lies outside the confidence interval predicted at the confidence level indicated in the x axis. **(b)** Absolute error in prediction for predictions across LOPO models stratified based on genome-wide sequencing depth. **(c)** Size of the predicted 90% confidence intervals across LOPO models stratified based on genome-wide sequencing depth. Box plots in **a-c** show the median, first and third quartiles (boxes) and the whiskers encompass observations within 1.5× the interquartile range from the first and third quartiles.

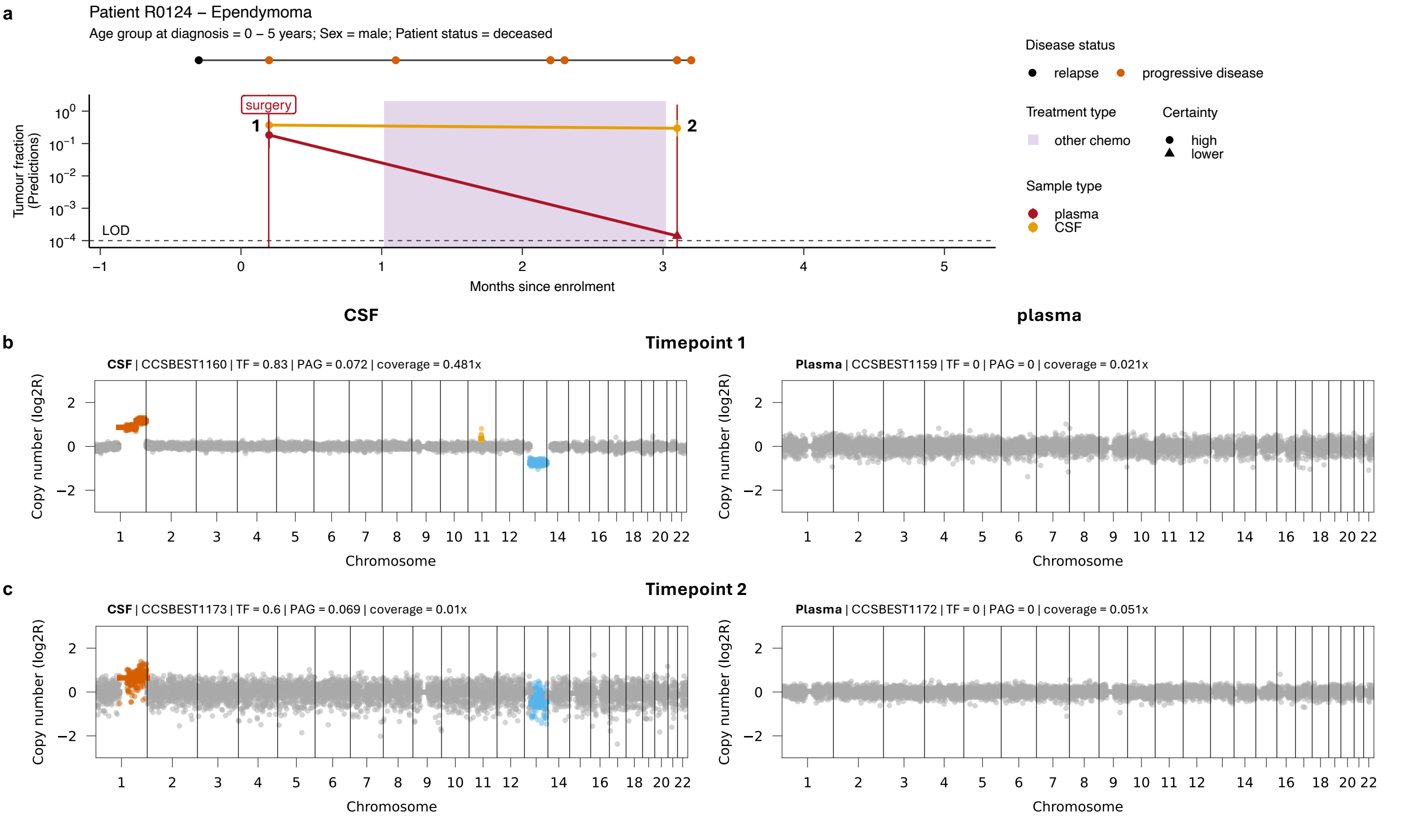


**Extended Data Figure 16. Longitudinal tracking of tumour burden using matched plasma and CSF cfDNA samples from ependymoma Patient R0124.**
**(a)** Longitudinal disease trajectory for a male ependymoma patient showing the TF predictions computed using multimodal CP obtained from time-matched plasma (red) or CSF (orange) samples. Predictions for which the 90% confidence interval goes beyond the LOD (as indicated by the dashed line) are considered of low certainty and are depicted as triangles. Treatment is indicated by coloured boxes. Disease status by RECIST 1.1 criteria are indicated for each cfDNA sample by coloured points. **(b-c)** Genome-wide COPYBARA copy number profile for matched CSF (left) and plasma (right) cfDNA samples for selected timepoints. Gains and amplifications are shown in yellow and orange, respectively. Losses and deletions are shown in light blue and dark blue, respectively.
CN deviation: copy number deviation; PAG: percentage abnormal genome; TF: tumour fraction.

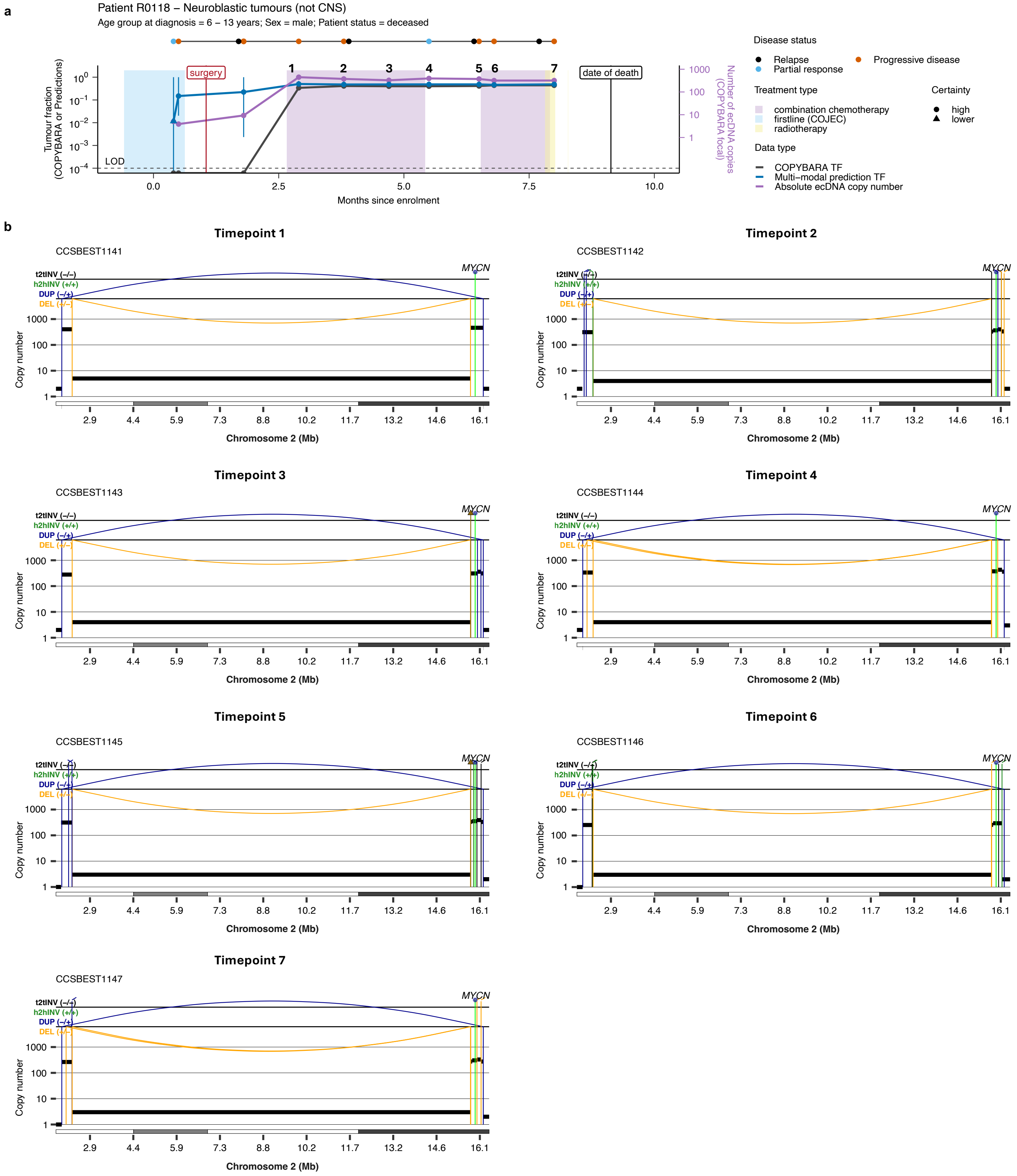


**Supplementary Figure 17. Longitudinal tracking of tumour burden using ecDNA and multimodal CP for neuroblastoma Patient R0018.**
**(a)** Longitudinal disease trajectory showing the TF predictions over time using either COPYBARA (genome-wide copy number analysis; dark grey) or predictions computed using multimodal CP (blue). 90% confidence intervals for TF predictions are shown in blue. Predictions for which the 90% confidence interval goes beyond the LOD (as indicated by the dashed line) are considered of low certainty and are depicted as triangles. The purple line shows the absolute ecDNA copy number detected using COPYBARA-focal informed by ecDNA-associated SV breakpoints. Treatment is indicated by coloured boxes. Disease status by RECIST 1.1 criteria are indicated for each timepoint by coloured points. **(b)** Rearrangement profiles showing the detection of the same ecDNA molecule at the indicated timepoints.

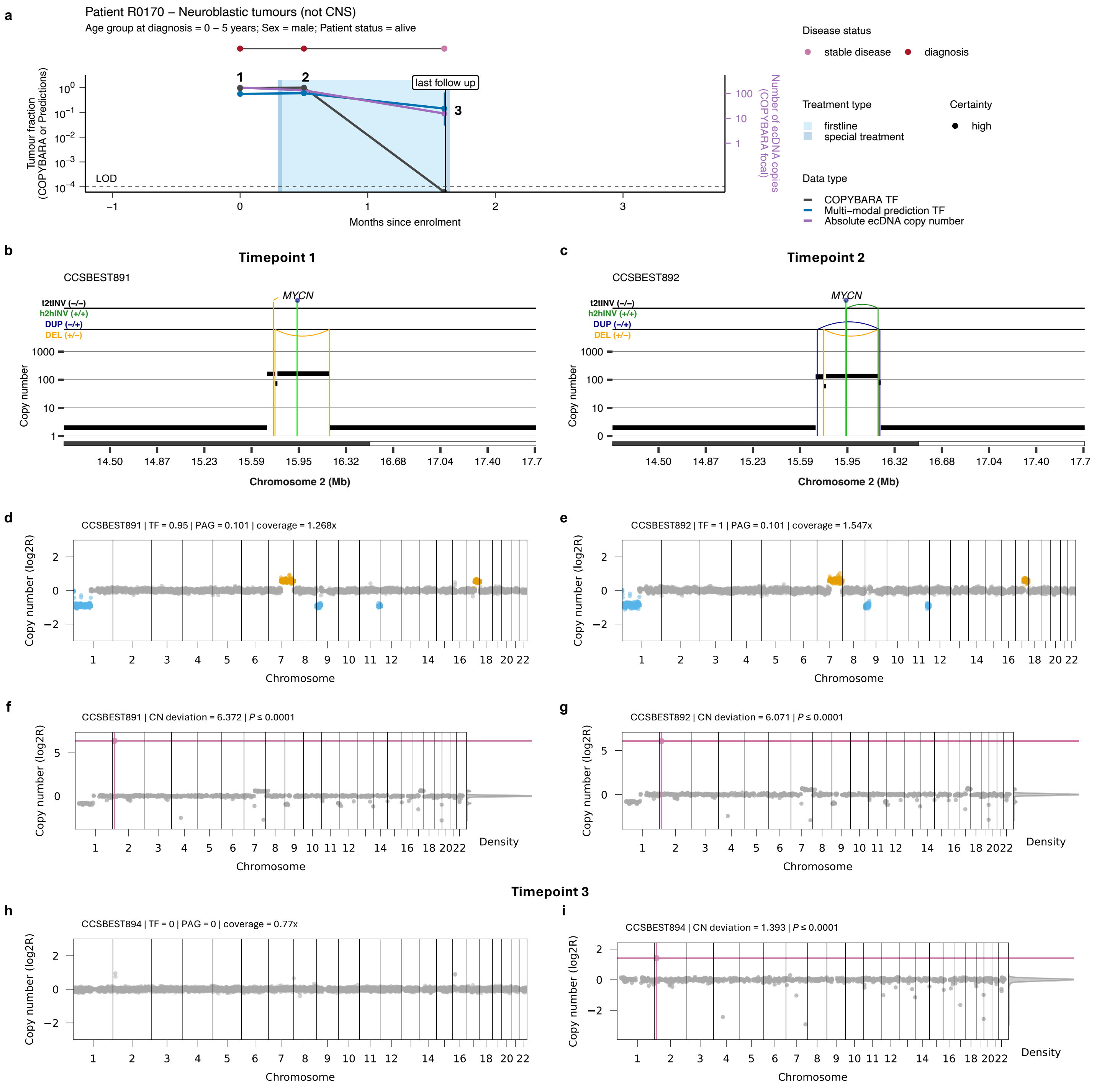


**Supplementary Figure 18. Longitudinal tracking of tumour burden using ecDNA and multimodal CP for neuroblastoma Patient R0170.**
**(a)** Longitudinal disease trajectory showing the TF predictions over time using ecDNA copy number (purple), COPYBARA (genome-wide copy number analysis; dark grey) and predictions computed using multimodal CP (blue). 90% confidence intervals for TF predictions are shown in blue. Predictions for which the 90% confidence interval goes beyond the LOD (as indicated by the dashed line) are considered of low certainty and are depicted as triangles. The purple line shows the absolute ecDNA copy number detected using COPYBARA-focal informed by ecDNA-associated SV breakpoints. Treatment is indicated by coloured boxes. Disease status by RECIST 1.1 criteria are indicated for each cfDNA sample by coloured points. **(b-c)** Rearrangement profiles showing the detection of the same ecDNA molecule at the indicated timepoints. Genome-wide COPYBARA copy number profiles (**d-e,h**) and detection of the ecDNA (pink dots; **f-g,i**) using COPYBARA-focal for selected timepoints. Gains and amplifications are shown in yellow and orange, respectively. Losses and deletions are shown in light blue and dark blue, respectively.
CN deviation: copy number deviation; PAG: percentage abnormal genome; SCNA: somatic copy number aberration; TF: tumour fraction.

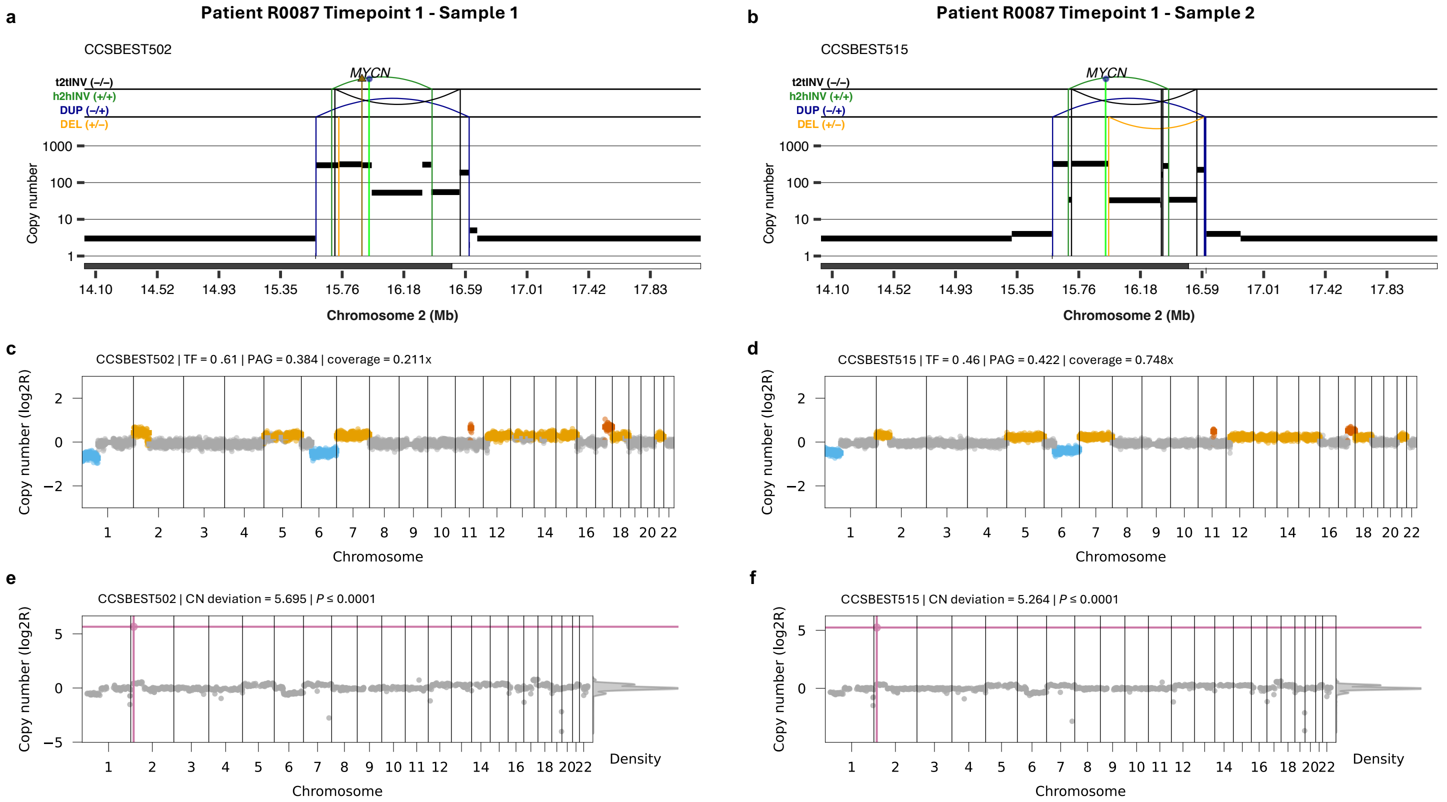


**Supplementary Figure 19. Detection of the same ecDNA molecule in two independent cfDNA samples from Patient R0087.**
**(a,b)** Rearrangement profiles showing the detection of the same ecDNA molecule in two cfDNA samples collected at the same timepoint. Genome-wide COPYBARA copy number profiles (**c,d**) and detection of the ecDNA (pink dots; **e,f**) using COPYBARA-focal for selected timepoints. Gains and amplifications are shown in yellow and orange, respectively. Losses and deletions are shown in light blue and dark blue, respectively.
CN deviation: copy number deviation; PAG: percentage abnormal genome; SCNA: somatic copy number aberration; TF: tumour fraction.
